# Effectiveness of Canadian travel restrictions in reducing burden of SARS-CoV-2 variants of concern

**DOI:** 10.1101/2023.09.12.23294140

**Authors:** Angela McLaughlin, Vincent Montoya, Rachel L. Miller, Canadian COVID-19 Genomics Network (CanCOGeN) Consortium, Michael Worobey, Jeffrey B. Joy

## Abstract

Evaluating travel restriction effectiveness in mitigating infectious disease burden is critical for informing public health policy. Here, we quantify where and when variants of SARS-CoV-2 were introduced into Canada to evaluate the extent to which travel restrictions averted viral introductions and COVID-19 case burden. Our results suggest that, across SARS-CoV-2 variants of concern subject to travel restrictions, at least 281 introductions were prevented, accounting for an averted burden of approximately 44,064 cases. This corresponds to approximately 441 averted hospitalizations, 24 averted deaths, and cost savings to Canadian health care systems of approximately 11.2 million Canadian dollars. Travel restrictions were found to be most effective when implemented rapidly during exponential case growth in the focal source and when global circulation was limited. Our analyses reveal that COVID-19 travel restrictions mitigated case burdens and highlight their value in future pandemic response.

**Summary:** COVID-19 travel restrictions against variants worked and were most effective when implemented rapidly and preceding new variants’ wider circulation.

## Introduction

Emergence and successive sweeps of SARS-CoV-2 variants of concern (VOCs) with elevated transmissibility, immune evasion, and/or virulence (*1*) have challenged effectiveness of COVID- 19 non-pharmaceutical interventions (NPI) and vaccines. Reconstructing the emergence and spread of VOCs can illuminate NPI effectiveness in mitigating viral burden, informing policy for ongoing and future pandemics. Phylogenetic analyses of SARS-CoV-2 genomes can be applied to infer viruses’ geographic and temporal origins (*2–4*), in addition to informing dynamic nomenclature systems (*5–7*), estimating key epidemiological metrics such as the effective reproduction number (*8*), and corroborating or refuting epidemiological linkage through contact tracing (*9*, *10*).

Travel restrictions are a class of NPI applied to mitigate pandemic burden; they include restricted entry of foreign nationals, flight bans, border entry requirements such as testing or vaccination, and quarantine requirements. Globally, evidence for their effectiveness has been mixed (*11–15*). Some evidence suggests countries where VOCs were first detected had diminishing contributions towards international dispersal over time, particularly those with high global connectivity, calling into question targeted travel restriction effectiveness on domestic case burden (*16*). Canadian COVID-19 travel restrictions were employed for travellers from regions where VOCs were first detected (**Fig. 1B** and **Supplemental Materials**), but their effectiveness in limiting introductions and case burden has not been characterized.

**Fig. 1.**
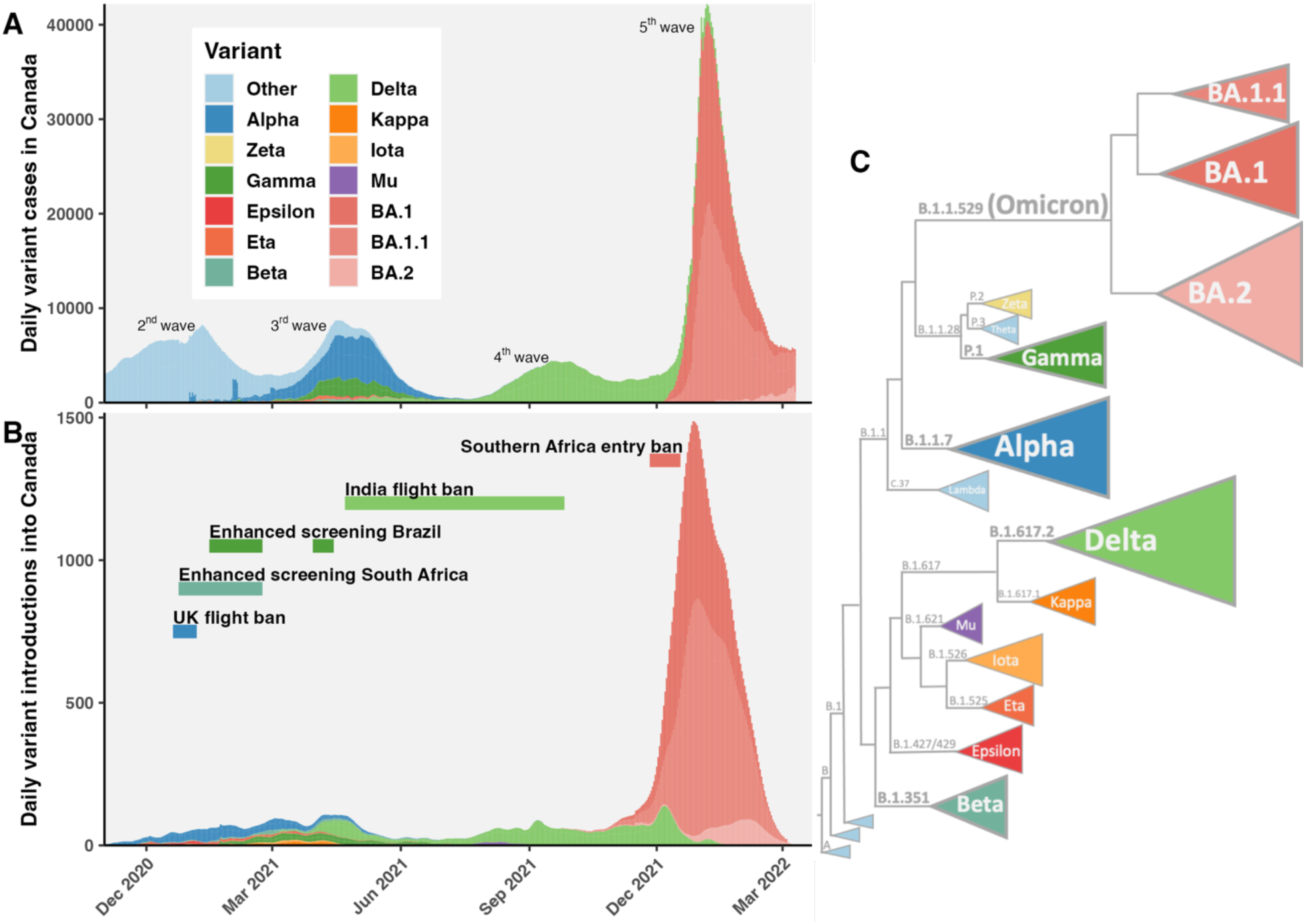
Epidemiological dynamics of SARS-CoV-2 variants in Canada. **(A)** Estimated daily cases of variants of concern (VOC) and (VOI) in Canada from November 1, 2020 until March 22, 2022. (**B)** Daily variant introductions into Canada, overlaid with timing of VOC-specific travel restrictions in Canada. Introductions include sublineages, international viral introductions into Canada with sampled descendants, and singletons, international viral introductions without sampled descendants. (**C)** Cartoon phylogeny of SARS-CoV-2 VOCs and VOIs scaled approximately by divergence from the root.

Previous evidence suggested the March 2020 entry ban for foreign nationals effectively reduced viral importations into Canada, but was insufficient to prevent new SARS-CoV-2 sublineages from seeding the second wave (*4*). We hypothesized that subsequent Canadian SARS-CoV-2 variant-specific travel restrictions were associated with reduced introduction rates from focal sources. Second, we hypothesized that these reduced importation rates would translate into reductions in Canadian case burden, and reduced economic impacts. To test these hypotheses, we inferred phylogeographic models of SARS-CoV-2 variant dispersal into and within Canada to evaluate the effectiveness of variant-specific travel restrictions in reducing importations and averting cases. We proportionally subsampled publicly available GISAID sequences available up to March 22, 2022, after which testing frequency dropped, to estimate variant-specific cases for each Canadian province or country. We then inferred maximum likelihood phylogenetic trees for each VOC and variant of interest (VOI) and reconstructed ancestral geographies to estimate the timing, origins, destination, and spread of variants into and within Canada. We estimated the effectiveness of flight bans for Alpha, Delta, and Omicron variants, or enhanced screening for Gamma and Beta variants, in averting viral importations and cases, using a counterfactual approach, whereby expected introductions in the absence of restrictions were predicted in relation to VOC cases in focal sources and travel volume. Our findings highlight that monitoring and statistically analyzing the emergence, introduction, and spread of novel sublineages is crucially important to guide public health responses to ongoing and future epidemics.

## Results

### Epidemiological synopsis of SARS-CoV-2 variants in Canada

This study spans from November 2020 (2^nd^ wave), when the first VOCs were identified in Canada, until March 2022 (5^th^ wave), dominated by early Omicron lineages BA.1 and BA.2 (**Fig. 1**). Daily variant cases were estimated for each Canadian province by multiplying confirmed diagnoses by the frequency of variants in the GISAID database. While the 2^nd^ wave (August 2020 – March 2021, Fig. 1) was primarily comprised of wild type SARS-CoV-2 lineages (‘Other’), VOCs and VOIs gained their foothold during this time. By the 3^rd^ wave (March 2021 – July 2021), wild type lineages were outcompeted primarily by Alpha (307,680 cases in Canada; **Table S3**), with notable contributions of Gamma (93,762 cases), Beta (18,326 cases), Eta (19,067 cases), Iota (8,325), Epsilon (5,591 cases), Zeta (2,906), Kappa (2,838), and Mu (405) across Canada, and even low-level detection of Delta lineages (first sampled in Canada on March 6, 2021). The maximum daily cases of the 2^nd^ and 3^rd^ waves were comparable, with the trough between the 3^rd^ and 4^th^ waves in summer 2021 low enough to justify relaxation of interventions. However, the 4^th^ wave spanning August 2021 – December 2021 driven by Delta brought significant burden on the Canadian health care system with 500,617 estimated Delta COVID-19 cases. Early Omicron lineages drove a 5^th^ wave with 630,408 BA.1 cases, 741,437 BA.1.1 cases, and 38,009 BA.2 cases cumulatively in Canada by March 22, 2022.

### Viral introduction rate reductions from focal sources following travel restrictions

VOC-specific travel restrictions were variably effective in lowering viral introduction rates from focal regions of first detection. The UK flight ban from December 20, 2020 – January 6, 2021 (duration: 17 d; delay from first sample: 44 d) in response to Alpha was associated with a non-significant 1.49 (95% confidence interval, two-sided t-distribution: 0.87-2.12)-fold reduction of the sublineage importation rate within two weeks and a 1.15 (0.91-1.39)-fold reduction of the proportion of sublineages from the UK (**Figs. 2A, 3**). However, the importation rate held steady amid exponentially rising Alpha cases in the UK (**Fig. S23A)** and then rebounded following ban repealment, rising to a maximum of 7.6 (6.3-8.8) sublineages from the UK per week on March 3, 2021. There were at least 672 (652-693) Alpha introductions into Canada, including 234 (228-240) sublineages and 438 (418-459) singletons. The UK contributed 50% (46-53%) of sublineages prior to the flight ban, 46% (39-53%) during the ban, and 37% (35-40%) after (**Fig. S30**). Altogether, 40% (38-42%) of sublineages and 45% (43-48%) of singletons originated in the UK (which represented 20% of international sequences), followed by Europe, the origin of 31% (29-32%) of sublineages and 23% (21-25%) of singletons, and the USA, contributing 23% (22-24%) of sublineages and 26% (25-28%) of singletons.

**Fig. 2.**
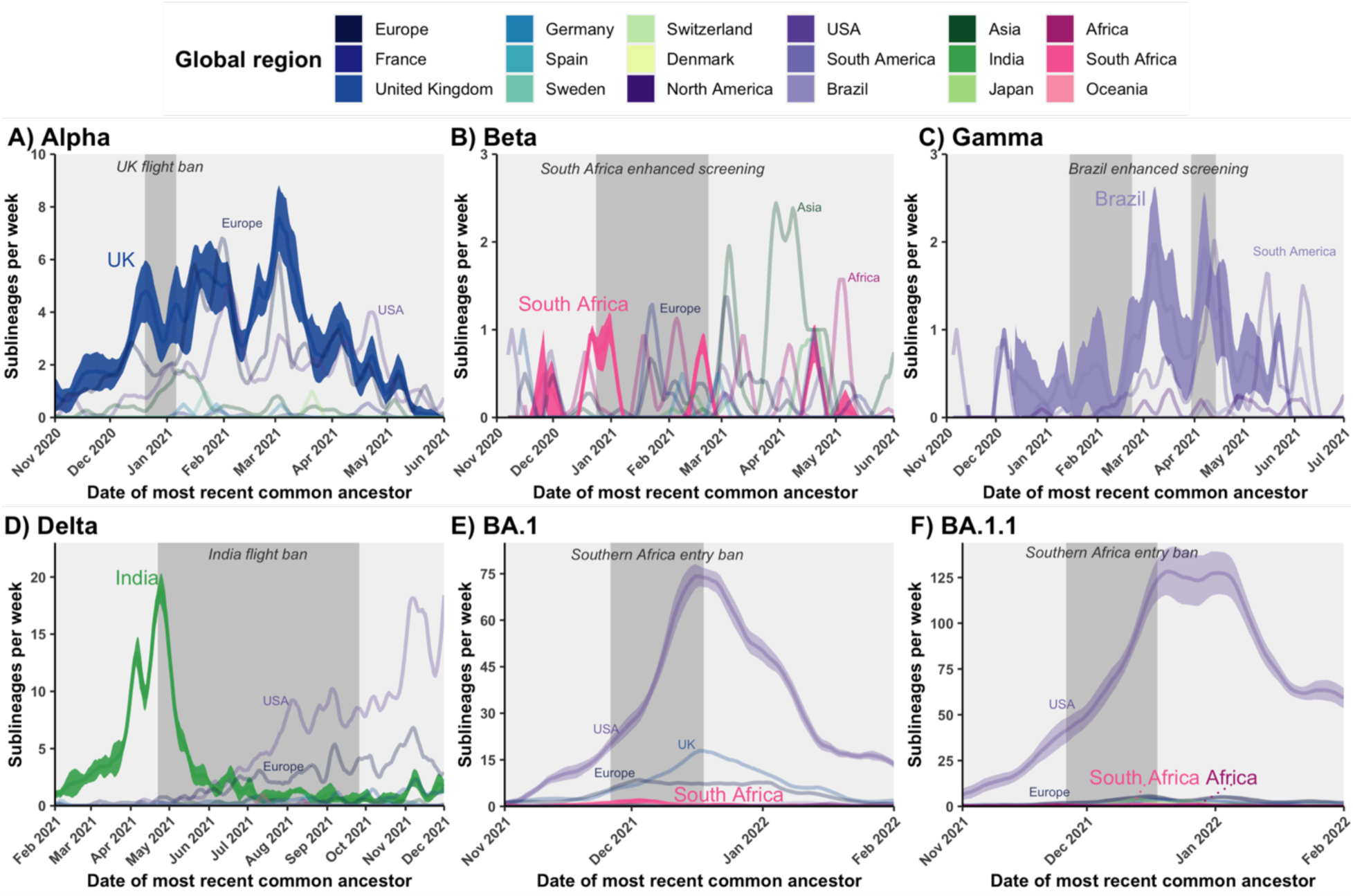
VOC sublineage introduction dynamics into Canada in the context of COVID-19 travel restrictions. The 7-day rolling average weekly sublineage introduction rates were inferred using maximum likelihood phylogeography for **(A)** Alpha, **(B)** Beta, (**C)** Gamma, (**D)** Delta, and (**E)** Omicron, BA.1 and **(F)** BA.1.1. Most introductions of Omicron BA.2 came after travel restrictions and thus BA.2 was excluded here. Global regions contributing most sublineages were directly annotated. 95% confidence intervals across 10 bootstraps for focal regions of first VOC detection are displayed and additionally for the USA for BA.1 and BA.1.1. Travel restriction durations for each variant shown with shaded grey rectangles.

**Fig. 3.**
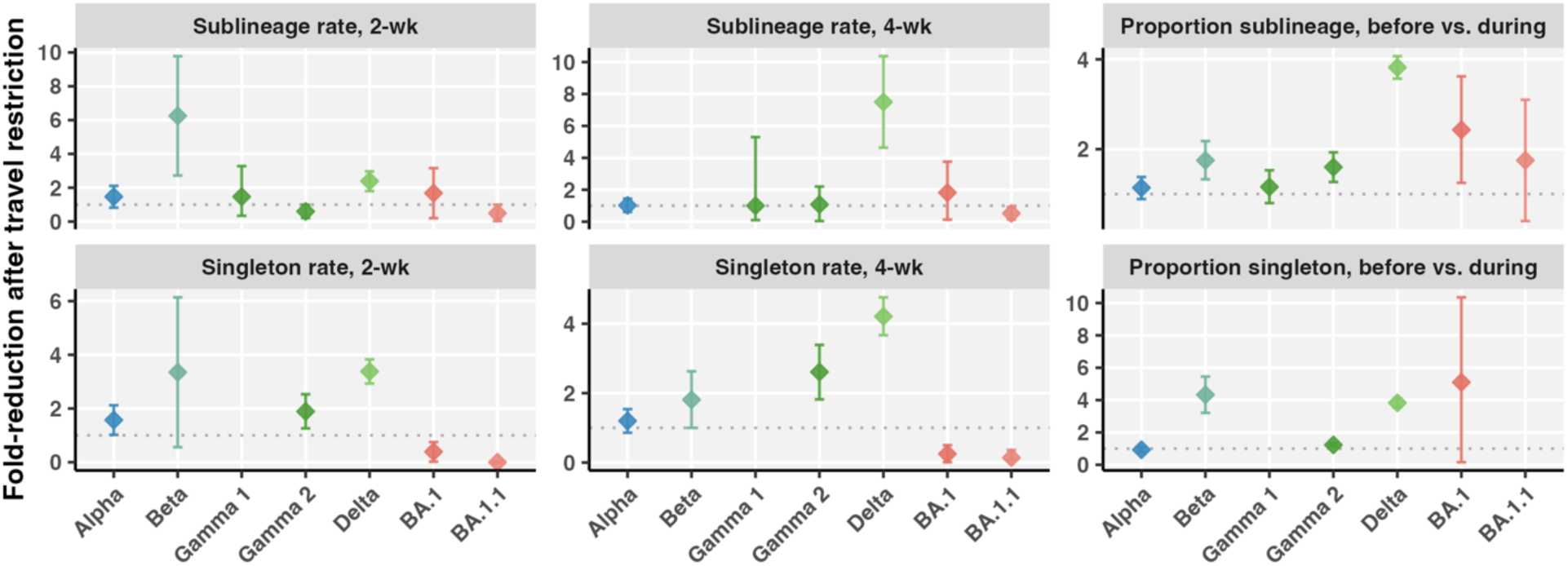
Comparison of fold-reduction in VOC importations from focal sources following travel restrictions. Metrics include fold-reduction of the 7-day rolling average sublineage importation rates or singleton importation rates from focal sources two and four weeks after restrictions began; and fold-reduction of the proportion of sublineages or singletons originating from focal sources before versus during travel restrictions. Diamonds show the mean and vertical lines depict 95% confidence interval across ten bootstraps. Estimates were not shown where comparisons included a zero. Dotted line at y=1 represents the null expectation of no change.

For Beta, enhanced screening and quarantine enacted for travellers who had been to South Africa, effective December 24, 2020 - February 22, 2021 (duration: 60 d; delay from first sample: 5 d), was associated with a significant 6.25 (2.72-9.78)-fold reduction of the Beta sublineage importation rate from South Africa, as well as a 1.75 (1.33-2.18)-fold reduction of the proportion of sublineages from South Africa (**Figs. 2B, 3**). During the restriction, there was a rise in Beta sublineages likely to have originated in Europe and other African nations; following restrictions, Beta importations from Asia increased. Overall, of 40 (40–41) total estimated sublineages, 14% (12-16%) were likely from South Africa, 31% (30-33%) from Asia, and 20% (18-22%) from other African nations (**Fig. S39**). Of 80 (77-83) total singletons, 8% (7-9%) were likely from South Africa, while 41% (39-42%) were from Asia, 22% (19- 24%) from Europe, and 18% (17- 20%) from other African nations.

Neither of the Gamma-related restriction periods with enhanced screening for travellers from Brazil was associated with significant reduction in sublineage importations from Brazil into Canada (**Figs. 2C, 3**). The second period of restrictions from March 30 to April 14 (duration: 15 d) was associated with a significant 1.6 (1.27-1.93)-fold reduction in the proportion of sublineages from Brazil; however, the number of Gamma sublineages observed during this period (n=3) was relatively low. Brazil was the likely origin for 52% (50–55%) of the 38 (37–40) Gamma sublineages in Canada, while the USA accounted for 29% (26-32%), other South American nations for 12% (10-14%), and other North American nations for 6% (5-8%) (**Fig. S33**). Of 159 (148-170) total Gamma singletons, 65% (63-67%) were likely from Brazil.

Suspension of flights from India to counter the Delta variant from April 22, 2021 to September 26, 2021 (duration: 157 d; delay from first sample: 47 d) was associated with a significant 2.4 (1.8- 3.0)-fold reduction of the sublineage importation rate from India within two weeks and 7.5 (4.6- 10.4)-fold within four weeks from a maximum of 19.0 (17.5-20.5) sublineages per week on April 24 (**Fig. 2D**). The singleton importation rate from India was also significantly reduced (**Figs. 3, S45**). By late June 2021, Delta sublineage importation from the USA and Europe superseded India, and for the USA in particular, continued to rise to a maximum of 20.7 (16.8-24.6) sublineages per week on December 4, 2021. There were at least 1822 (1794-1850) Delta introductions into Canada, including 537 (521-553) sublineages representing 90 (86-93) unique Pango lineages, and 1285 (1253-1317) singletons. The flight ban was additionally associated with a 3.8 (3.6-4.1)-fold reduction in the proportion of sublineages originating from India (Fig. 3). Prior to the restriction, 95% (93-97%) of 76 (72-80) Delta sublineages were from India, compared to 25% (23-26%) of 228 (218-237) sublineages introduced during the restriction (**Fig. S43**). After the flight ban, 233 (225-240) additional sublineages were introduced, primarily from the USA, Europe, and with a lesser contribution from India. Overall, the USA was the likely origin for 48% (47-49%) of Delta sublineages and 50% (49-52%) of singletons, followed by India for 26% (25-27%) of sublineages and 21% (20-22%) singletons, and then Europe for 19% (18-20%) of sublineages and 20% (19- 21%) of singletons (**Fig. S44**). India and the USA accounted for 15% and 42%, respectively, of global sequences sampled. The flight ban was also associated with a reduction of sister lineage Kappa (B.1.617.1) sublineage and singleton importation rates from India (**Fig. S42**).

The Omicron-related entry ban for foreign nationals arriving from southern African nations and enhanced screening for Canadians who had been to southern Africa (duration: 22 d) was largely ineffective towards reducing importations of BA.1 and BA.1.1. No BA.2 was known to have been introduced prior to the travel restrictions; therefore, we cannot comment upon the effect travel restrictions may have had on BA.2 introductions to Canada. The flight ban was not significantly associated with a reduction in the BA.1 or BA.1.1 sublineage importation rates from southern Africa two weeks following the restriction (1.7 (0.2-3.2)-fold reduction in BA.1 and 0.49 (-0.03 - 1)-fold reduction in BA.1.1; **Figs. 2E-F, 3**). There was a significant 2.4 (1.3-3.6)-fold reduction in the proportion of BA.1 sublineages from southern Africa following the ban, which changed from 3% (1-5%) of sublineages with southern African origins before restrictions to 1% (1-2%) during restrictions (**Fig. S50**); however, this coincided with increased importation rates from the USA, reaching a maximum of 74.2 (71.0-77.4) BA.1 sublineages per week on December 17, 2021 (**Fig. 2E**). There was no significant reduction (1.75-fold reduction, 0.4-3.1) in the proportion of BA.1.1 sublineages from southern Africa two weeks following restriction implementation. Before, during, and after the restrictions, BA.1.1 sublineages in Canada predominantly originated from the USA, which was the source of up to 128.2 (114.9-141.6) sublineages per week on December 21, 2021 (**Fig. 2F**). By contrast, the maximum flow of BA.1.1 from southern Africa was 0.93 (0.1-1.76) BA.1.1 sublineages per week on November 18, 2021 and 1.8 (0.9-2.7) BA.1 sublineages per week on December 3, 2021. The USA was the predominant source of Omicron BA.1 and BA.1.1 into Canada up to March 2022, contributing 72% (70-73%) of 615 (596-633) BA.1 sublineages (**Fig. S50**), 90% (89-91%) of 1148 (1103-1194) of BA.1.1 sublineages (**Fig. S51**), 74% (73-75%) of 3634 (3524 - 3744) BA.1 singletons, and 92% (91-93%) of 5572 (5337 - 5806) of BA.1.1 singletons. Africa and South Africa combined likely contributed 1% (1-2%) of BA.1 sublineages, <1% BA.1.1 sublineages, 2% (1-2%) of BA.1 singletons, and <1% BA.1.1 singletons. Predominant sources of early BA.2 introductions were India and the UK, and negligibly few BA.2 introductions originated from the USA or Africa (**Fig. S52**). Sequences from the USA represented 40% of international sequences subsampled for BA.1 and 66% for BA.1.1; i.e., USA introductions exceeded what would be expected by chance.

Comparing VOCs according to fold-reduction in sublineage importations (**Fig. 2, 3**), Delta and Beta restrictions were associated with significant reductions in sublineage importation rate from focal sources 2- and 4-weeks following implementation and the proportion of sublineages from the focal source. However, Beta restrictions were not associated with significant reductions in singleton rates from South Africa, and estimates have wide uncertainty due to rare events. The second Gamma intervention was associated with significant reductions in singleton importation rates from Brazil 2- and 4-weeks after implementation, but not sublineage importation rate. The Delta-related India flight ban was associated with significant reductions in both absolute and relative sublineage and singleton contributions from India to Canada within two and four weeks of implementation. However, these reductions in importations were eventually eclipsed by the influx of sublineages from the USA and other countries not specified in the respective travel restrictions.

### Introductions and cases averted via VOC travel restrictions

We applied counterfactual models relating introductions to variant cases in focal sources to quantify how many sublineages, singletons, and cases were averted due to travel restrictions. Averted travellers were estimated based on epochal and seasonal trends in international air arrivals (**Supplementary Methods**).

In the absence of the UK flight ban, we estimated there were likely to have been 724 additional travellers from the UK, 12 (9-16) additional Alpha sublineages that could have resulted in upwards of 5,682 (3,849-7,132) descendant cases, as well as 58 (44-73) singletons (**Fig. 3A**). The ban became more effective over time as daily Alpha cases in the UK rose during the latter part of the flight ban (**Fig. S23**). Significantly more sublineages, singletons, and cases were averted per day in the late period (December 28, 2020 – January 6, 2021) versus early period (December 20-27, 2020), with over three times as many sublineages averted per day in the late period (**Fig. 4**). In totality, these 5,740 (3,893-7,205) averted cases represent an additional 1.8% (1.2-2.2%) of all the 324,547 estimated Alpha cases detected in Canada. When normalized to the restriction duration of 17 days, an average of 338 (229-424) cases were averted per day during the UK flight ban; which for the late period was even higher at 482 (380-637) cases averted per day.

**Fig. 4.**
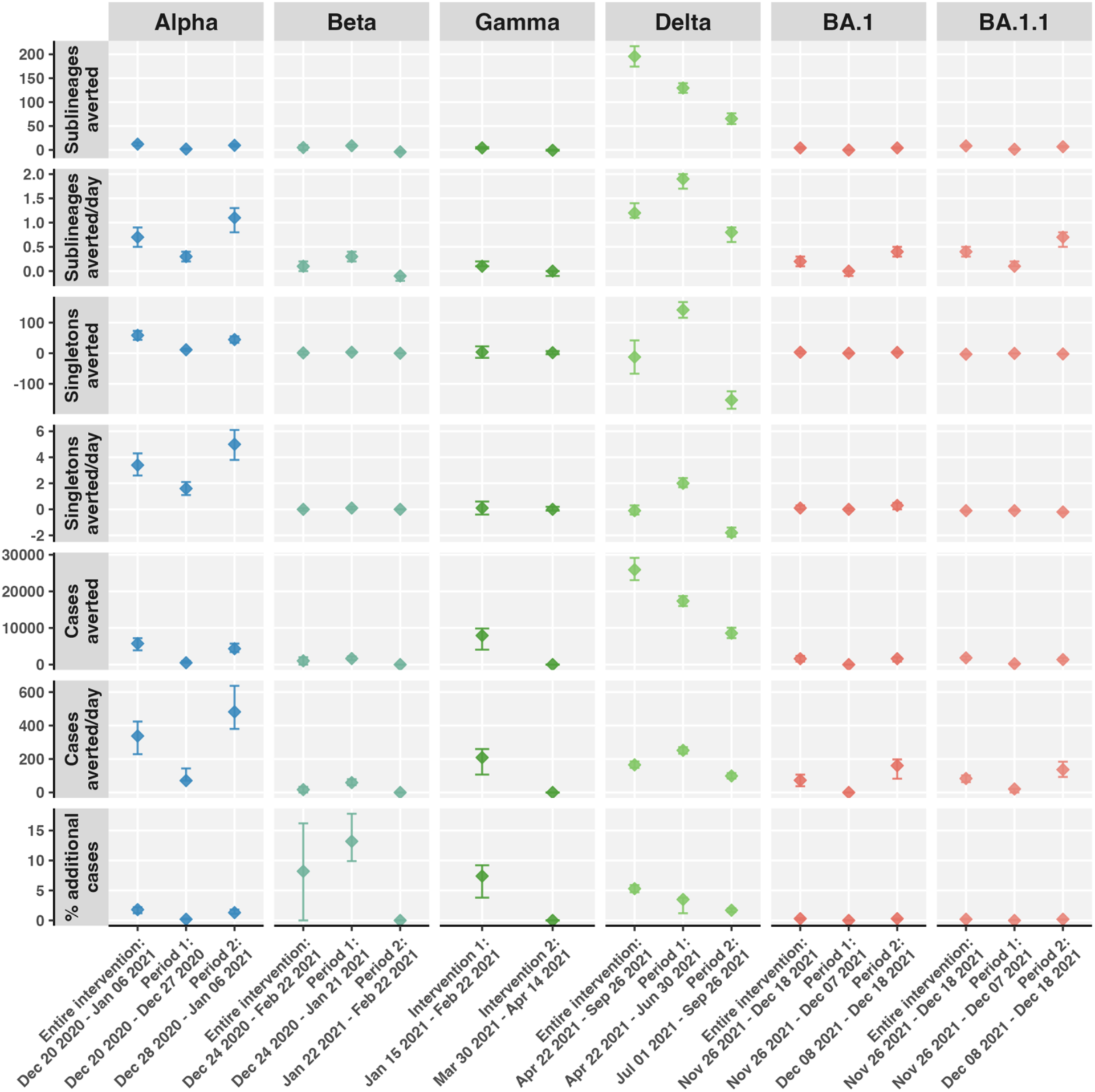
Averted introductions and cases attributable to VOC travel restrictions. Sublineages, singletons, and estimated cases averted overall and normalized to intervention duration; and percent additional cases averted relative to total variant cases in Canada. Metrics were estimated for entire intervention and for early and late periods of the intervention. Gamma interventions were considered only separately.

Enhanced screening for travellers from South Africa countering the Beta variant averted only 3 travellers, 5 (0-10) Beta sublineages, and 1 (-4-6) singletons (**Fig. 4**). Consistent with Alpha, the early (24 December 2020 – 21 January 2021) and late (22 January – 22 February 2021) periods of the ban displayed contrasting dynamics, although for Beta, effectiveness lessened. The early period was associated with 9 (6-11) averted sublineages that could have resulted in 1,645 (1,551-1,739) descendant cases, while the late period was associated with -4 (-6--2) averted sublineages (more observed than expected in the absence of the ban). Assuming their conservative net sum of 5 (0- 10) sublineages averted, there were 1,019 (0-2,111) descendant cases averted. These averted cases would have represented an additional 8.2% (0-16.2%) of the 12,462 total estimated Beta cases in Canada (**Fig. 3B; Fig. S24**). The average effectiveness for Beta was relatively low at 17 (0-34) cases averted per day, but the late period may have averted up to 59 (44-79) cases per day.

Initial enhanced screening for travellers from Brazil countering Gamma averted more travellers, introductions, and cases than the second intervention period (**Fig. 4**; **Fig. S25**). The first intervention averted 326 travellers, 4 (3-6) sublineages that may have given rise to 7,932 (4,081- 9,839) descendant cases, and 4 (-15-22) singletons. Averted sublineages during the first period had a relatively high probability of being large based on the right-skewed distribution of observed Gamma sublineage sizes until the end of restrictions. 7,935 (4,066-9,862) total averted cases during the first intervention represents an additional 7.4% (3.8-9.2%) of the estimated Gamma cases in Canada, and an average effectiveness of 209 (107-260) cases averted per day. The second intervention period was less effective, likely averting 58 travellers, -1 (-2-0) sublineages, and 2 (- 3-7) singletons.

The India flight ban was effective in averting many travellers, sublineages, singletons, and cases, however it diminished in effectiveness as Delta cases increased globally (**Fig. 4**). Over five months, the ban likely averted 5,148 travellers from India. The number of daily travellers averted was steadily high from late April to mid-June 2021, dropping in July 2021 before becoming erratic with rising travel volume via other countries, which was permitted with a negative PCR test (**Figures S19**). 63 Delta sublineages were observed during restrictions, whereas we predicted there would have been 259 (237-280) sublineages imported without restrictions, amounting to 196 (174- 217) Delta sublineages averted throughout the entire flight ban (**Fig. S26**). There were more sublineages averted in the first period of the ban up to the end of June (2 (2-2) sublineages averted per day) compared to July through September when the ban ended (1 (1-1) sublineages averted per day). Averted sublineages could have given rise to 25,935 (22,431 – 29,891) cases. An additional 525 (338-711) singletons were averted or 3 (2-5) singletons averted per day. More singletons were averted in the first period of the ban, 6 (5-7) per day, versus the second period, with 1 (0-2) per day. In totality, 26,460 (23,474-29,819) cases were likely averted. Relative to the 491,733 total estimated Delta cases in Canada, this would have represented 5.4% (4.8-6.1%) additional sampled Delta cases. Overall, 165 (147-186) cases were averted per day during the Delta ban; in the first period, 252 (232-271) cases were averted per day, which decreased to 99 (83-116) in the second period. We were unable to test the effectiveness of the two-month flight ban for Pakistan because the sequence representation was low enough to warrant its grouping with Asia; however, we estimated 116 travellers from Pakistan were averted (**Fig. S22**).

For Omicron, the entry ban for foreign nationals and enhanced screening for Canadians who had travelled to southern Africa nations was largely ineffective in reducing Omicron burden (**Fig. 4**). Although it may have averted 1,681 travellers from Africa, 4 (2-6) BA.1 and 8 (6-11) BA.1.1 sublineages, as well as 1,607 (823-2349) BA.1 cases and 1,838 (1364-2247) BA.1.1 cases, these were negligible compared to the massive burden of Omicron. The cases averted by southern Africa entry restriction represented only 0.3% (0.1-0.4%) of 634,334 BA.1 cases and 0.2% (0.2-0.3%) of 779,903 BA.1.1 cases confirmed by March 10, 2022. Overall intervention effectiveness was 73 (37-107) BA.1 and 84 (62-102) BA.1.1 cases averted per day. The second period of the intervention was more effective than the first period of the intervention at 161 (82-198) BA.1 cases and 137 (93-184) BA.1.1 averted per day. Omicron cases per day in South Africa and other African nations were higher in the second period of the intervention than the first (**Fig. S27, S28**). We did not find any evidence of BA.2 sublineages introduced from either South Africa or Africa, and only one singleton which likely was introduced in mid-February (**Fig. S53**).

Overall, variant-specific restrictions were variably effective, but conservatively may have averted at least 281 introductions and 44,064 additional cases in Canada. The majority of cases averted were Delta (25,922), followed by Gamma (7,937 v), Alpha (5,740), Omicron BA.1.1 and BA.1 (3,445), and Beta (1,020). The restrictions that averted the highest percentage of cases relative to the variant case burden observed in Canada were Beta (8.2%, with the caveat of wide uncertainty and relatively low burden), the first Gamma intervention (7.4%), Delta (5.3%), Alpha (1.8%), and lastly, Omicron BA.1 (0.3%) and BA.1.1 (0.2%). Comparing restrictions’ effectiveness in terms of cases averted per day, Alpha was the most effective (338 per day), followed by the first period of Delta (252 per day), the first Gamma intervention (209 per day), and Omicron BA.1 and BA.1.1 (157 per day).

Based on cases averted, average COVID-19 hospitalization and death rates, and estimated public healthcare cost of a COVID-19 patient in Canada, we conservatively estimated averted hospitalizations and deaths, and corresponding costs averted. Between May 2020 and April 2021, the average COVID-19 hospitalization rate in the USA was estimated as 1093.9 per 100,000 (1.1%) (*17*). Therefore, without considering differences in variant virulence or age demographics, we coarsely assume 1% of COVID-19 cases averted could have been hospitalized. The average cost per COVID-19 patient hospitalization in Canada was $15,000 for those not in ICU and $55,000 for those admitted to ICU, with financial information up to May 2023 (*18*). Across data up to March 2022, 21% of hospitalized COVID-19 patients were admitted to the ICU (*18*), of whom 56% received ventilation and 25% died in the facility. Correspondingly, approximately 441 COVID-19 hospitalizations were averted, of which 93 may have been admitted to ICU, and 24 may have died. The cumulative medical cost of these averted instances is approximately $11,214,364 Canadian dollars (CAD). We do not consider the additional costs (and suffering) incurred by families and friends, funeral and estate costs, or diminished economic activity due to employee absences.

## Discussion

Our findings suggest that Canadian COVID-19 travel restrictions were variably effective towards reducing SARS-CoV-2 VOC importations and cases, but cumulatively may have averted more than 280 introductions, 44,000 cases, 440 hospitalizations, and 24 deaths; conservatively corresponding to an estimated savings greater than $11.2 million CAD. We developed a novel methodology to estimate introductions and cases averted which applies a counterfactual model considering estimated variant cases in focal origins, travel volume, and their empirical relationships with introductions into Canada. This enabled us to both quantify the overall effectiveness of interventions on case burden, but also quantify effectiveness over time, providing retrospective insights into when restrictions could have ended.

The India flight ban against Delta introductions was the most effective restriction, corresponding to a large decline in the importation rate from India and averting over 30,000 additional cases. However, the ban’s effectiveness decreased past June 2021, after which Delta cases and importations from the USA and other countries rose and cases in India decreased. Regionally coordinated policy towards the Delta lineage by Canada, USA, and other high volume travel hubs may have led to further durability in travel restriction effectiveness. Other analyses of Delta transmission dynamics in Canada found early introductions of AY.25 and predecessor of AY.27 (which may have arisen in Canada) drove Delta burden in Canada, corroborating the futility of later stages of the flight ban (*19*). Repeated extension of the India flight ban was unnecessary past June 2021, although was likely guided by the precautionary principle due to concerns related to case underreporting and Delta’s elevated virulence and transmissibility (*20–22*). The ban should have been implemented earlier and relaxed earlier in light of evidence that Delta was globally and domestically widespread.

Our results suggested the UK flight ban targeting Alpha increased in effectiveness as cases climaxed in the UK, while the Beta enhanced screening for South Africa and Gamma enhanced screening for Brazil became less effective over time and were potentially leakier. Omicron-targeted southern Africa entry bans were largely unsuccessful in reducing importations or case burden, as BA.1 and BA.1.1, followed by BA.2, BA.4, and BA.5, were introduced via other global sources. In instances where the proportion of sublineages from an area was reduced following travel restrictions, but the sublineage importation rate was not significantly reduced, the interpretation is that there the travel restriction had a minor effect. When a new variant has emerged and global regions have similar case trajectories and stable relative travel volumes, then the proportion of introductions should remain constant in the absence of restrictions or if restrictions were entirely ineffective. Where the sublineage importation rate and proportion of sublineages decrease in association with travel restrictions, there is a major effect, particularly if cases were rising in the region targeted by the restriction. The counterfactual approach we have taken allowed us to take into consideration variant cases and travel in the targeted nations to predict sublineage importation rates without restrictions.

Globally, evidence relating to travel restrictions’ effectiveness in reducing COVID-19 burden is mixed. Several reviews on the topic highlight a lack of consensus in methodologies for empirically evaluating COVID-19 interventions’ effectiveness, including different definitions or criteria for effectiveness (*11*, *12*, *23*). In United Arab Emirates, a decline of transmissions from international sources followed implementation of international travel restrictions in 2020 (*14*). In Zimbabwe, initial public health interventions delayed onset of community transmission following international introduction during the first waves of COVID-19 (*24*). The implementation of international travel restrictions and stringent domestic interventions within a zero-COVID policy until the end of 2021 in New Zealand eliminated community transmission until the arrival of the Delta variant (*21*). Other studies found intermediate effectiveness, such as in South Korea, where international traveler quarantine in 2020 reduced introductions and spread of SARS-CoV-2, but control was incomplete (*25*). Mandatory hotel quarantines in response to the Delta variant reduced onward transmission of Delta importations in England, but subsequent NPI relaxation drove rapid transmission of early introductions (*26*). As for evidence of low effectiveness, restriction of travel from Brazil to the USA did not prevent establishment of the Gamma variant in New York City, as the majority of Gamma transmission lineages had already been established at least two weeks prior originating from other regions in the USA (*13*). In Hong Kong, border control measures were deemed insufficient to prevent outbreaks, especially during a period of lower stringency of social restrictions (*15*). While this by no means comprises a comprehensive review of evidence, these examples highlight the lack of consensus regarding how to measure effectiveness and consider the epidemiological circumstances in which a measure was enacted. What can be distilled from this body of work is that, as implemented, travel restrictions do not stop all introductions, but they can reduce the flow, and their impact depends upon the epidemiological, immunological, and stringency context in which they are invoked, as well as their coordination with neighboring countries.

Many factors can impact travel restriction effectiveness and should be weighed at the time of implementation. Of first consideration are the magnitude and rate of change in prevalence in the country where a variant is first detected, as well as in other geographies that could act as sources. Additionally, importations and cases were most effectively averted when restrictions were implemented rapidly. Part of the reason the Omicron ban may have been less effective is that it was discovered on November 19, 2021, over a month following its estimated origin around October 9, 2021 (*27*). By the time restrictions were implemented for travel from southern Africa, Omicron had already dispersed globally, and the USA was a dominant early exporter of BA.1 globally (*16*). The Delta-related flight bans commenced 47 days after the first Delta case was sampled in Canada, and Beta-related measures began 5 days after first detection. The extent of variant spread during this delay is affected by the focal geographies’ global travel volume and degree of connectedness (*16*), as well as a variant’s inherent transmissibility. Another factor is a variant’s antigenic distance and immune evasion potential, which in combination with immunological waning from natural infections and vaccines, determine the extent of population- level susceptibility and thus potential for introductions to become large outbreaks. Outbreak sizes can also be minimized in the presence of domestic NPIs, such as restrictions on social gathering or mask requirements that reduce contact rates or transmission probabilities, and higher compliance. Legal and logistical travel loopholes could compromise restriction effectiveness by increasing border porosity. For the Delta-related flight bans, there was a loophole whereby travellers from India and Pakistan could enter Canada indirectly through intermediary flights, but were required to present a negative COVID-19 test upon arrival (*28*). Travel volume from India expanded during later stages of the ban (**Fig. S14**). Variant virulence and rates of hospitalizations and deaths are also important to consider for overall health care burden.

There are multiple sources of uncertainty and limitations in this analysis. Publicly available genome sequences and confirmed diagnoses over time are affected by differences in case ascertainment across geographies, time, variants, case severity, and sociodemographic groups. Therefore, daily new diagnoses based on PCR confirmed positive cases do not equate to daily new infections. Within any given jurisdiction, there were changes in frequency of symptomatic infection and hospitalization, testing criteria and public health recommendations, testing capacity, and availability of rapid antigen tests (for which results were not always reported). One way to consider changes in age-based testing criteria is to associate fluctuations in case counts among the widely tested 70+ age group, for which testing recommendations did not change in late 2021, with cases in other age groups (*29*). However, this approach fails to consider differential changes in behaviour and social contact across age groups in response to the pandemic, as well as does not consider other sources of ascertainment bias affecting all age groups. In addition to case ascertainment bias, jurisdictions varied in extent and reason for sequencing, as well as in their proclivity to share this data publicly. When analyzing empirical sequence data sets, where reason for sequencing could be due to hospitalization, association with an outbreak, travel history, or variant suspicion (spike gene target failure requiring variant confirmation, for example), we must interpret frequencies cautiously. An idealized surveillance strategy would be to longitudinally surveil a randomly selected cohort for infection (regardless of symptoms) with all cases sequenced, which would yield accurate estimates of the percent positivity (prevalence), variant frequencies, and levels of re-infection in the wider community. Accurate case counts are important to quantify burden, as well as to inform sequence subsampling strategies to minimize bias.

Accuracy of phylogeographic inferences has been shown to be high when the migration rate is low, however sampling differences across jurisdictions can contribute to biases (*30*). We developed a sequence subsampling strategy to reduce sampling bias in the publicly available sequence data, while also rendering the analysis computationally feasible for maximum likelihood phylogenetic inference. With moderately large subsamples of 50,000 sequences, regions that contributed high sequences per case were less overrepresented, but regions with few sequences available relative to cases remain underrepresented. With smaller subsamples, the ratio of sequences to cases can be more normalized but this reduces the data informing the model, and thus it is a fine balance of maximizing signal while ensuring representativeness. Differences in reason for sequencing are also not alleviated via subsampling. Future developments in subsampling and phylogeographic methodologies could profit from structured consideration of reason for sequencing, as well as carrying forward sampling proportion or case ascertainment in the phylogeographic inference. Recent methodological developments to include unsampled nodes in phylogeographic inference are promising, but are at present, not scalable to tens of thousands of sequences (*31*).

Inclusion of 50% Canadian and 50% global sequences in each subsample was based upon findings from previous sensitivity analyses, whereby this relative representation identified the maximum number of sublineages and singletons (*4*). Additionally, the 50% split imposes a prior expectation that any given sequence from Canada is equally likely to have a close relative from either Canada or global. One might ask, if we are interested in quantifying primarily introduction dynamics from focal sources, why not include more sequences from the focal source relative to other global regions? Doing so would increase the chance of misattributing the origin of a sublineage to the focal source when it could have come from a different country. While the focus is on the dynamics of the focal source sublineage importation rate, contributions from other regions remain important. By treating the focal source equally to other global regions at the subsampling stage, we do not impose a bias towards the focal source in the ancestral reconstruction; thus, our estimates represent lower bounds of sublineages per week, but proportionally and temporally, should be accurate. By subsampling multiple bootstraps with replacement, we considered how the relative inclusion of sequences and geographies affect uncertainty in our estimates of importations and sublineage sizes.

In addition to uncertainty as to whether sequence sets were representative of the pandemic in Canada and globally, there was also uncertainty due to phylogenetic tree inference, time-scaling the tree under a relaxed molecular clock model, and ancestral character estimation of geographic origins. For 50,000 tips, it is impossible to search the entire tree space for the most likely tree, therefore the trees analyzed represent local maxima. Furthermore, this pipeline requires scaling trees from units of divergence to time, which presupposes a molecular clock rate can be generalized for the tree. Even though a relaxed molecular clock rate was assumed with no fixed mean, a prior variance of 0.2 across the tree was assumed, which could misrepresent the underlying evolutionary process. By partitioning tree inference by variant with a small subset of parental lineage sequences, we have partially accounted for differences in molecular clock rates across variants, including during their emergence. Tip dates inform rescaling the tree, but themselves can be speckled with errors, for instance due to delay between infection and sampling, data entry errors, and in some cases, incomplete metadata collection dates. Incomplete dates in the Canadian metadata were dealt with by randomly sampling from potential days. This added some uncertainty to the analysis, particularly in regards to the timing of sublineage introduction dates. Incomplete dates in metadata associated with Canadian samples from particular regions were an ongoing issue despite calls for public access to full data to enable genomic epidemiological surveillance (*32–34*). In previous analyses for the first two waves of SARS-CoV-2, incomplete tip dates were inferred using least squares dating (*35*), which slowed the time-scaling of trees and in some cases was intractable due to insufficient memory.

A comprehensive understanding of how variant dynamics were impacted by travel restrictions requires consideration of changes in the environment, host, and virus. Human behavioral changes relating to contact networks and transmission probability via other non-pharmaceutical interventions should be considered in relation to any detected changes in transmission. Further, heterogeneity in population-level immunity due to vaccination and infection confound our ability to identify changes in sublineage characteristics, such as transmissibility or lifespan. Disentangling the effects of public health interventions from viral genetic adaptations in regards to migration, transmission, and health care burden is a formidable challenge. VOCs vary in their selective advantages to outcompete previously circulating diversity (*36*), which we have not explicitly considered in these models. This was indirectly considered by stratifying variants in the analysis.

Methodological developments on estimating variant-specific reproduction numbers and fitness advantages offer some promise in this regard (*37*). Of course, as with all models, we have endeavoured to infer sufficiently complex models with as few assumptions as possible while maintaining generalizability. Further developments in bridging phylogeographic inference with mathematical modelling could be fruitful in this regard (*38*).

We applied travel volume data in our phylogeographic methods by using it to adjust our null expectation of travel volume from a given country if there had not been a travel restriction imposed on that country. By considering seasonal and epochal changes in travel volume into Canada, we estimated travel volume in the absence of restrictions and travellers averted. This approach considers broad changes in propensity or ability to travel, considering discovery of new variants or changes in vaccination and testing requirements. Although we attempted to adjust models relating sublineage flows to variant cases by travel volume, in practice, these models only improved the goodness-of-fit for the BA.1 model. Further explorations of travellers compared to importations averted are warranted. Another way to use flight data is to model exportation intensities from different regions as the product of the estimated number of infected people in a given geography at a given time by travel volume into a country of interest. This has been applied in place of inferring transmission lineages’ origin locations when introductions were identified using binary state ancestral reconstruction (*3*). Another way to use flight data is to investigate whether phylogeographically inferred importation rates are consistent with expected importation intensities. When available, individuals’ travel history can also be used as priors in in travel-aware phylogeographic models (*31*).

The questions facing the global community to respond to and learn from SARS-CoV-2 have evolved, as have the methodologies used to approach these questions (*39*). Combining phylogenetics, mathematical modelling, and machine learning could improve the accuracy of infectious disease forecasting (*38*). For instance, phylogenetics offer insight into taxonomic groupings for model structuring, estimates of migration rates, and evolutionary rates, which can be incorporated into compartmental models. Models would also benefit from improved variant-specific estimates of selection in the context of vaccine- and natural infection-induced immunity. Longitudinal prospective cohort studies of generalizable populations (i.e., not just those above the age of 70) would be helpful in estimating frequencies of circulating variants or sublineages, positivity rates, period of infectiousness, incidence, and the true sampling rate. Accurate estimations of case ascertainment and relatedly, sampling rates, in the broader population might be estimated compositely from knowledge for specific variants of the difference between expected percent positivity from the cohort vs observed in broader data, as well as knowledge of reason for sequencing in the publicly available data.

However, we have provided evidence that they reduce the flow of pathogens across borders, particularly when implemented rapidly following initial detection. In instances where viral establishment is nearly inevitable, travel restrictions can contribute to flattening the curve to mitigate the burden on the health care system with limited finite resources. If a government adopts a travel restriction policy to limit burden, we recommend simultaneously adopting other NPI domestically, like installing adequate ventilation and promoting mask use, to prevent introductions from becoming widespread sublineages.

Travel restrictions to curb importations from countries where variants are first detected should be considered alongside other public health interventions when sufficient evidence suggests there is a high risk to individual- and population-level human health; equally, they should be repealed if there is strong evidence that the variant is already established or poses a low risk to health. Formalizing coordination among neighboring jurisdictions would increase the odds of successful restrictions. Sustained investment in developing genomic surveillance systems, including in wastewater, cohort surveys, and in wildlife, will enable rapid detection and monitoring of circulating variants. Ongoing deposition of anonymized pathogen sequence data into the public repository, with time and date of sampling, is important to facilitate genomic epidemiology studies.

## Conclusion

We have presented robust and novel methods for quantifying effectiveness of public health interventions in reducing pathogen introductions and averting cases, which can contribute to the evidence base considered for future implementation and policy. Ongoing funding and prioritization of viral genomic surveillance programs in humans and the animals will help to mitigate future pandemic threats and curtail the ongoing burden of the COVID-19 pandemic. Without global surveillance, we do not know how widely a variant has spread or how quickly it is spreading, limiting our ability to confidently react to the emergence of new variants or pathogens.

We conclude by emphasizing that it is critical to consider that travel restrictions can and have had very negative socioeconomic impacts. As such, they provide a strong disincentive to countries that are first to detect new SARS-CoV-2 variants of concern to make those discoveries public. This issue is one that we expect to continue as new subvariants and perhaps entirely new variants of SARS-CoV-2 emerge through time. Even more concerningly, these disincentives can be predicted to contribute to hesitation (or complete reluctance) to report future emerging pathogens on the part of countries first to detect them, likely the countries where they originated. This may contribute to the loss of a window of opportunity to prevent a potential new pandemic from being actualized. The scientific evidence presented here that travel restrictions can have public health benefits needs to be viewed in that larger context.

## Methods

### Sequence data sets and curation

9,487,106 SARS-CoV-2 sequences and associated metadata (298,892 collected in Canada) were downloaded from Global Initiative on Sharing All Influenza Data (GISAID) on March 22, 2022 (*40*, *41*). GISAID clade partitions were maintained throughout data cleaning to reduce computational burden. Sequences were excluded if they were listed on the Nextstrain exclude list updated on day of data download (n=8,782) (*7*, *42*), duplicate IDs (n=4,284), from a non-human host (n=6,803), environmental samples (n=4,907), or had incomplete dates outside of samples from Canada (n=218,233). For Canadian sequences with incomplete dates, days were randomly sampled within known months. If only year was provided, the sequence was discarded.

Sequences were aligned to Wuhan-Hu-1 (GenBank ID: MN908947.3) using the viralMSA python wrapper of minimap2 (*43*, *44*). Sequences were excluded if ambiguous sites or gaps exceeded 10% (n=7,968 and n=657,063, respectively), resulting in 8,585,792 total clean sequences (274,568 from Canada). Pango lineages in the metadata were called using pangolin v3.1.20 and pangoLEARN data release March 22, 2022 (*5*, *6*).

Total clean sequences available were grouped by variant (**Table S3**): Alpha (B.1.1.7, Q.*), Beta (B.1.351), Gamma (P.1), Delta (B.1.617.2, AY.*), Epsilon (B.1.429 and B.1.427), Zeta (P.2), Eta (B.1.525), Iota (B.1.526), Kappa (B.1.617.1), Mu (B.1.621), and Omicron (separately: BA.1, BA.1.1, and BA.2). Any sequence not identified as a variant was grouped into ‘Other’. Variants with fewer than 100 sequences from Canada were excluded (Lambda, Theta, GH/490R, and BA.3). The primary manuscript focuses on the five VOCs, Alpha, Beta, Gamma, Delta, and Omicron BA.1, BA.1.1, and BA.2, for which there were targeted travel restrictions. Omicron was split into its comprising Pango lineages as their spatiotemporal origins into Canada differed. The earliest global sample collection dates for each variant were obtained from cov-lineages.org (*45*) and outbreak.info (*46*) and earliest Canadian dates were pulled from clean GISAID data (**Table S1**). Canadian sequences with collection dates preceding the earliest global date for each variant were removed.

### Estimating variant-specific cases by Canadian province and global region

Canadian daily new COVID-19 diagnoses by province were obtained from the Public Health Agency of Canada (*47*). Global daily new diagnoses by country were obtained from the R package coronavirus (*48*), which pulls data from the Johns Hopkins University Center for Systems Science and Engineering Coronavirus repository. Rolling 7-day averages of daily new diagnoses were calculated to smooth the data. For global regions, countries were grouped into continents unless their sequence contributions were within the top 95^th^ percentile, which included Brazil, Denmark, France, Germany, India, Japan, South Africa, Spain, Sweden, Switzerland, UK, and USA.

Average daily variant-specific cases in Canadian provinces and global regions were calculated as the product of average daily new diagnoses and average daily variant frequency based on clean GISAID sequences (**Figs. S4, S6**). Total monthly contributions to variant-specific cases were calculated for each province and global regions, then tallies were grouped by variant to calculate geographies’ proportional contributions to cases of each variant for each calendar month (**Fig. S5, S7**). Monthly proportional contributions of each geography to variant-specific cases were used to inform the probability of subsampling a sequence from a given geography and month.

### Subsampling data to reduce spatiotemporal bias

For each of ten bootstraps, 50,000 sequences were subsampled where possible, including 50% sampled in Canada and 50% sampled globally, which was identified as an optimal split of sequences in previous subsampling sensitivity analyses (*4*). The temporal distribution of sampled sequences reflected the distribution of monthly estimated variant-specific cases for Canada and global regions, respectively (**Fig. S8**). If there were fewer sequences available than the target of 25,000 for a given variant in Canada or global, then all sequences were retained. The proportion of variant cases occurring in each month was multiplied by the total number of target sequences to obtain monthly targets. If fewer sequences were available in any given month than the target, then all sequences from that month were taken. If the target sequences for a given month were available, but sparse (fewer than 500) during the first half of the months where sequences are available, then the number of target sequences was specified as 500 (**Fig. S8**). Remaining sequences were redistributed among months where there were not sparse sequences, and the loop repeated until there were no sparse months and the target number of sequences sampled was achieved. Relative frequencies of variant cases, sequences available (**Table S3**), and sequences sampled (**Figs. S10, S11**) were summarized.

A small number of sequences were additionally included in each sample set from all parental lineages basal to variants, i.e. for B.1.1.7, sequences were included from lineages B, B.1, and B.1.1. For B, Wuhan-hu-1 was sampled; and for other parental lineages, ten sequences with a collection date preceding that of first global sample for the variant were sampled at random from the global dataset for each bootstrap independently. This was done to provide phylogenetic structure for early branching events and inform the estimation of the timing of variant emergence.

Sequences were subsampled according to the temporal distributions described above, with probabilities equal to monthly contribution of global regions or Canadian provinces to variant- specific cases, up to the total number of target sequences per month. Subsampling was repeated for ten bootstraps with replacement. Similar subsampling approaches based on countries’ cases or deaths have been applied elsewhere (*16*, *49*).

All 1,752,808 unique subsampled genome sequences from 200 countries and territories were collated into GISAID EPI_SET_230510yr, accessible at doi:10.55876/gis8.230510yr and via the attached Supplemental Table provided by GISAID. This serves as the data acknowledgement for all contributing and submitting laboratories. Those without GISAID Access Credentials may retrieve information about all data contributors by either clicking on the DOI or pasting the EPI_SET ID in the “Data Acknowledgement Locator” on the GISAID homepage.

### Maximum likelihood phylogenetic inference and ancestral state reconstruction

Problematic sites were censured from sequence alignments prior to phylogenetic inference according to de Maio et al. VCFv4.3 (*50*). Approximate maximum likelihood trees were generated using FastTree v2.1.11 under a generalized time reversible substitution model and the ‘-fastest’ algorithm (*51*). Trees were outgroup rooted on Wuhan-hu-1 in R package ape (*52*). A linear regression of root-to-tip distance versus time was generated for each tree, excluding parental lineage sequences. Tips with absolute value residuals greater than 0.001 or terminal branch lengths longer than 20 mutations were excluded as temporal outliers. Trees were time-scaled using IQ- TREE 2.1.2 with least squares dating (LSD2) (*35*, *53*), a relaxed molecular clock, 0.2 relative variance, and 50 bootstrap resamples to estimate internal branch lengths. Polytomies were randomly resolved.

Phylogeographic reconstruction of internal nodes’ geographic state as Canadian province or global region was conducted using maximum likelihood discrete ancestral character estimation in R package ape with symmetrical rates (*52*). The highest likelihood state was pulled for each internal node. Canadian sublineages were designated where Canadian internal nodes were preceded by a non-Canadian internal node, signifying an international introduction resulting in onward sampled transmission. Singletons were defined as Canadian sequences with non-Canadian parental origin and no sampled descendants. Sublineage and singleton importation rates were summarized as 7- day rolling mean of importations per week by global region, province of introduction, and Pango lineage. The sum of sublineages and singletons represents a lower limit for the total number of introductions due to subsampling. Rate estimates were reported as the mean across bootstraps, with 95% confidence intervals calculated using the t-distribution, *μ* ± *t* × *σ*/√*n*.

Summary metrics included the fold-reduction of sublineage and singleton importation rates from focal sources two weeks after implementation of restrictions versus before, and fold reduction in proportion of sublineages from focal sources compared to other regions before versus during restrictions. For either metric, the null hypothesis was that restrictions had no effect on either importation rates or proportion of importations from the focal source; therefore, if the 95% confidence intervals do not overlap one, we reject the null hypothesis and have evidence to support restrictions’ effectiveness.

### Estimating cases averted by restrictions

We estimated cases averted due to travel restrictions for each VOC by comparing observed introduction rates to expectations in the absence of travel restrictions, as predicted from a mechanistic counterfactual model of the relationship between introduction rates and variant cases in the focal source, trained on data preceding the intervention. Linear models consistently had the best goodness-of-fit in comparison with Poisson and negative binomial models. Training data were restricted to the week preceding the restriction or where variant cases in the source region exceeded a threshold chosen to maximize the correlation between cases and importations (described for each VOC in supplementary; **Figs. S23-28**). Observed daily variant cases in the focal source during the intervention and expected travel volume were used to predict sublineages per week in the absence of restrictions. Likelihood ratio tests were applied to evaluate inclusion of travel volume as a covariate; it was included for BA.1.1.

Statistics Canada data on international arrivals (*54*) was used to quantify observed and estimate expected travel volume from the focal sources in the absence of restrictions based on seasonal and epochal travel changes (**Supplementary Material**). Areas under curves of observed and predicted sublineages per week (converted to per day) were calculated to obtain numbers of sublineages observed and predicted during restrictions, the difference of which equates to sublineages averted. Sublineages averted was calculated for entire intervention durations and for early and late periods (**Fig. 4**). For each averted sublineage, sublineage size (number of sampled descendants) was drawn from VOC-specific gamma distributions fitted to sublineage sizes observed until the end of restrictions. Cases averted from averted sublineages were summed. This was repeated 1000 times to estimate a mean number of total descendant cases averted for the mean and confidence limits of sublineages averted. Confidence intervals were estimated by carrying forward upper and lower limits of importation rates estimated across bootstraps.

The model fitting process was repeated for singletons by fitting models of singletons per week to variant cases, predicting singletons per week in the absence of restrictions, and taking the difference in observed and expected singletons to estimate the number of singletons averted. Total cases averted is the sum of singletons averted and descendant cases from sublineages averted for each period. The percentage additional cases equals the ratio of total cases averted to total variant cases in Canada overall. Sublineages, singletons, and cases averted were normalized to days of restriction duration. Based on the cases averted and variant-specific hospitalization and death rates in Canada for each variant, we roughly estimated averted hospitalizations, healthcare costs, and deaths.

### Economic estimations

To approximate health care cost savings attributable to travel restrictions, we translated cases averted to COVID-19 hospitalizations, intensive care unit (ICU) visits, and deaths. Based on a literature search for case hospitalization rates, we assumed an average 1% hospitalization rate throughout the study period, as estimated for the USA between May 2020 and April 2021 of 1093.9 hospitalizations per 100,000 cases (1.1%) (*17*). The average costs of $15,000 for a COVID-19 hospitalization in Canada not in ICU and $55,000 for those admitted to ICU were obtained from the Canadian Institutes for Health Information (CIHI) (*18*). We also assume CIHI estimates that 21% of COVID-19 hospitalizations resulted in ICU admission, and that 56% of ICU patients received ventilation and 25% died in the facility (*18*).

## Supporting information

GISAID Epi Set

CanCOGeN Consortium Authorship

## Acknowledgments

We gratefully acknowledge all originating and submitting laboratories of SARS-CoV-2 genome sequences and metadata on GISAID. We also thank members of the Canadian COVID-19 Genomics Network (CanCOGeN) Consortium (**Supplementary file of contributions**) and the Canadian Public Health Laboratory Network (CPHLN) for their contributions towards publicly available data.

## Author contributions

Conceptualization: AM, JBJ.

Methodology: AM, VM, JBJ

Investigation: AM Visualization: AM

Funding acquisition: AM, JBJ

Project administration: JBJ

Supervision: JBJ

Writing – original draft: AM, JBJ

Writing – review & editing: AM, VM, RLM, MW, JBJ

## Data Availability

Viral genome sequences analyzed in this study were sourced from the Global initiative on sharing all influenza data (GISAID) coronavirus (CoV) database and are subject to the GISAID EpiFlu Database Access Agreement. Within this agreement, we cannot distribute data to any third part other than Authorized Users. A full set of all subsampled genome sequence accession IDs is available at GISAID EPI_SET_230510yr (doi:10.55876/gis8.230510yr), which serves as the data acknowledgement for all originating and submitting laboratories. Those without GISAID access credentials may retrieve information about all data contributors by either clicking on the DOI or pasting the EPI_SET ID in the "Data Acknowledgement Locator" on the GISAID homepage. Full and subsampled alignments can be shared to Authorized Users upon request. Scripts used for data curation, inferences, analyses, and visualization are available at github.com/AngMcL/sars-cov-2_variants_canada.

https://doi.org/10.55876/gis8.230510yr

