## Supplementary material for "Effectiveness of Canadian travel restrictions in reducing burden of SARS-CoV-2 variants of concern": GISAID Epi Set

### SUPPLEMENTAL TABLE

#### Data Availability

| | |
|---|---|
| GISAID Identifier: | EPI_SET_230510yr |
| doi: | 10.55876/gis8.230510yr |

All genome sequences and associated metadata in this dataset are published in GISAID's EpiCoV database. To view the contributors of each individual sequence with details such as accession number, Virus name, Collection date, Originating Lab and Submitting Lab and the list of Authors, visit 10.55876/gis8.230510yr

#### Data Snapshot

- EPI_SET_230510yr is composed of 1,752,808 individual genome sequences.
- The collection dates range from 2019-12-31 to 2022-03-17;
- Data were collected in 200 countries and territories;
- All sequences in this dataset are compared relative to hCoV-19/Wuhan/WIV04/2019 (WIV04), the official reference sequence employed by GISAID (EPI_ISL_402124). Learn more at https://gisaid.org/WIV04.
