## Supplementary material for "Effectiveness of Canadian travel restrictions in reducing burden of SARS-CoV-2 variants of concern": CanCOGeN Consortium Authorship

GenomeCanada

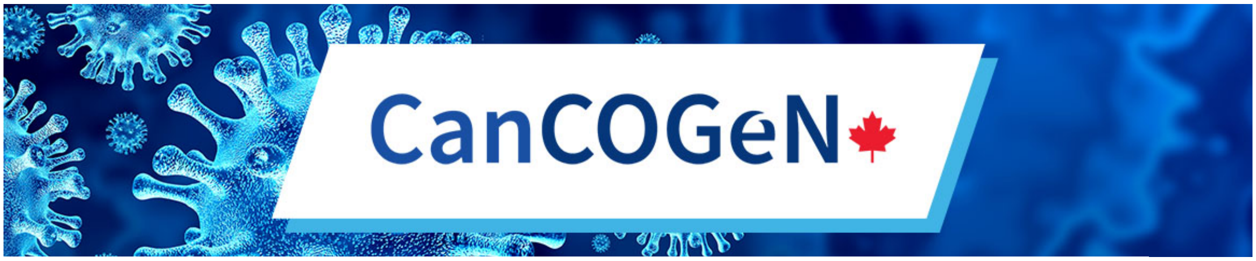


### The Canadian COVID-19 Genomics Network (CanCOGeN) consortium authorship

#### Steering Committee

Lorne Hepworth, Kenneth Baillie, Bettina Hamelin, Rob Annan, Ewan Harrison, Duncan MacCannell, Chris McMaster, Allison McGeer, Eric Meslin, Howard Njoo, Joris Veltman

#### Coordinating Committee

Catalina Lopez-Correa, Naveed Aziz, Fiona Brinkman, William Hsiao, Yann Joly, Steven Jones, Sandrine Moreira, Samira Mubareka, Natalie Prystajecky, Terrance Snutch, Lisa Strug, Gary Van Domselaar, Ma'an Zawati, David Alexander, Erin Gill, Jordan Lerner-Ellis, Calvin Sjaarda, Guillaume Butler-Laporte, Paul McLaren, Paul Vancaeseele, Diana Iglesias, Agnes Baross, Koko Agborsangaya, Megan Smallwood, Yoo Jin Park, Marc-André Langlois, Gerald Pfeffer

#### Data Sharing Committee

Art Poon, Fiona Brinkman, Catalina Lopez-Correa, Emma Griffiths, Eric Sutherland, , Gary van Domselaar, Gijs van Rooijen, Guillaume Bourque, Kieran O'Doherty, Kimberlyn McGrail, Jason Leblanc, Lisa Strug, Ma'n Zawati, Marc Fiume, Matthew Croxen, Michael Szego, Natalie Prystajecky, Sandrine Moreira, Steven Jones, Terrance Snutch, Will Hsiao, Yann Joly, Erin Gill Megan Smallwood, Eddy Nason, Koko Agborsangaya, Daryl Waggott.

#### Canadian Public Health Laboratory Network members and staffs having contributed data

##### *Cadham Provincial Laboratory (Manitoba); National Microbiology Laboratory*

Anna Majer, Shari Tyson, Grace Seo, Philip Mabon, Elsie Grudeski, Rhiannon Huzarewich, Russell Mandes, Anneliese Landgraff, Jennifer Tanner, Natalie Knox, Morag Graham, Gary Van Domselaar, Paul Van Caeseele, Jared Bullard, David Alexander, Kerry Dust, Nathalie Bastien, Yan Li, Timothy Booth, Darian Hole, Madison Chapel, Kirsten Biggar

***Hôpital Georges L. Dumont (New Brunswick); National Microbiology Laboratory***

Anna Majer, Shari Tyson, Grace Seo, Philip Mabon, Elsie Grudeski, Rhiannon Huzarewich, Russell Mandes, Anneliese Landgraff, Jennifer Tanner, Natalie Knox, Morag Graham, Gary Van Domselaar, Richard Garceau, Guillaume Desnoyers, Nathalie Bastien, Yan Li, Timothy Booth, Darian Hole, Madison Chapel, Kirsten Biggar

***Alberta Precision Labs; University of Alberta; University of Calgary***

Berenger B, Bernier F, Buss, E, Chui L, Croxen M, Deo A, Dieu P, Gill K, Ferrato C, Gordon P, Kellner J, Khan F, Koleva P, Lam LG, Li V, Lloyd C, Lynch T, Ma R, Melin A, Murphy S, Pabbaraju K, Shokoples S, Tipples G, Thayer J, Whitehouse M, Wong A, Yu C, Zelyas N

***Laboratoire de Santé Publique du Québec, McGill Génome Sciences Centre; CoVSeq Consortium***

Sandrine Moreira, Jiannis Ragoussis, Guillaume Bourque, Jesse Shapiro, Éric Fournier, Réjean Dion, Hugues Charest, Aurélie Guilbault, Benjamin Delisle, Sarah Reiling, Anne-Marie Roy, Shu-Huang Chen, Corinne Darmond, Sally Lee, Brent Brookes, Pierre Lepage, Jannick St-Cyr, Patrick Willet, Mathieu Bourgey, David Bujold, Hector Galvez, Paul Stretenowich, Pierre-Olivier Quirion, Romain Grégoire, Carmen Lia Murall, Julie Hussin, Raphaël Poujol, Jean-Christophe Grenier, Fatima Mostefai, Sylvie Laboissières, Alexandre Montpetit, Mark Lathrop, Michel Roger

***McMaster University***

Hooman Derakhshani, Sheridan J.C. Baker, Emily M. Panousis, Ahmed N. Draia, Jalees A. Nasir, Michael G. Surette, Andrew G. McArthur

***Toronto Invasive Bacterial Diseases Network; Sunnybrook Health Sciences***

Allison McGeer, Patryk Aftanas, Angel Li, Kuganya Nirmalarajah, Emily Panousis, Ahmed Draia, Jalees Nasir, David Richardson, Michael Surette, Samira Mubareka, Andrew G. McArthur

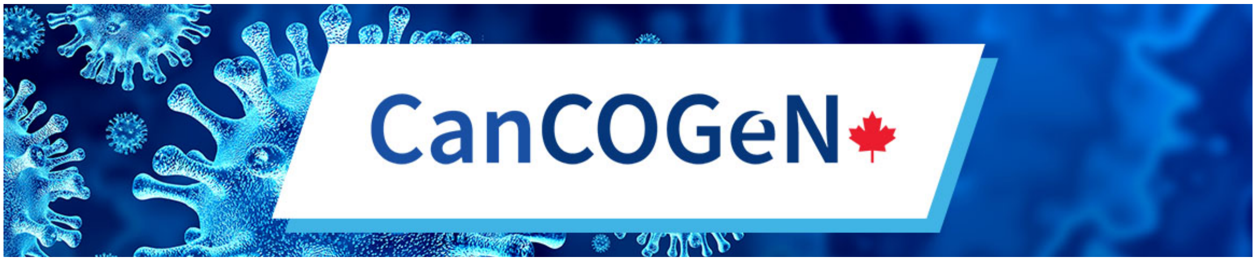


***Eastern Ontario Regional Laboratory Association***

Leanne Mortimer, Hooman Derakhshani, Emily Panousis, Ahmed Draia, Jalees Nasir, Robert Slinger, Andrew G. McArthur

***Public Health Ontario Laboratory***

Vanessa G Allen, Philip Banh, Yao Chen, Richard de Borja, Alireza Eshaghi, Nahuel Fittipaldi, Christine Frantz, Jonathan B Gubbay, Jennifer L Guthrie, Lawrence Heisler, Esha Joshi, Michael Laszloffy, Aimin Li, Michael C.Y. Li, Dean Maxwell, Sandeep Nagra, Samir N. Patel, Karthikeyan Sivaraman, Ashleigh Sullivan, Yogi Sundaravadanam, Sarah Teatero, Andre Villegas, Matthew Watson, Sandra Zittermann

***Ontario Institute of Cancer Research***

Jared Simpson

***Mount Sinai Hospital***

Jeff Wrana

***BC-CDC Public Health Laboratory***

Prystajecky Natalie, Linda Hoang, John R. Tyson, Dan Fornika, Shannon Russell, Kim Macdonald, Kimia Kamelian, Ana Pacagnella, Corrinne Ng, Loretta Janz, Richard Harrigan, Robert Azana, Mel Krajden, Jessica Caleta, Tara Newman.

***University of British Columbia***

John R. Tyson, Terrance P. Snutch

***Public Health Laboratory (Newfoundland) - Dr. Leonard A. Miller Centre for Health Services; National Microbiology Laboratory***

Anna Majer, Shari Tyson, Grace Seo, Philip Mabon, Elsie Grudeski, Rhiannon Huzarewich, Russell Mandes, Anneliese Landgraff, Jennifer Tanner, Natalie Knox, Morag Graham, Gary Van Domselaar, Robert Needle, Yang Yu, Adel Malek, Laura Gilbert, George Zahariadis, Nathalie Bastien, Yan Li, Timothy Booth, Darian Hole, Madison Chapel, Kirsten Biggar, Kerri Smith, Matthew Gilmour

***QEII Health Sciences Centre (Nova Scotia); National Microbiology Laboratory***

Anna Majer, Shari Tyson, Grace Seo, Philip Mabon, Elsie Grudeski, Rhiannon Huzarewich, Russell Mandes, Anneliese Landgraff, Jennifer Tanner, Natalie Knox, Morag Graham, Gary Van Domselaar, Todd Hatchette, Jason LeBlanc, Janice Pettipas, Dan Gaston, Nathalie Bastien, Yan Li, Timothy Booth, Darian Hole, Madison Chapel, Kirsten Biggar

**CanCOGeN VirusSeq committees and working groups' members:**

***CanCOGeN VirusSeq Implementation Committee***

Terrance Snutch, Fiona Brinkman, Marceline Côté, William Hsiao, Yann Joly, Sharmistha Mishra, Sandrine Moreira, Samira Mubareka, Jared Simpson, Gary Van Domselaar

***CanCOGeN Capacity Building Working Group***

Gary Van Domselaar, Matthew Croxen, Natalie Knox, Celine Nadon, Jennifer Tanner

***CanCOGeN Data Analytics Working Group***

Gary Van Domselaar, Fiona Brinkman, Zohaib Anwar, Robert Beiko, Matieu Bourgey, Guillaume Bourque, Ahmed Draia, Jun Duan, Marc Fiume, Dan Fornika, Eric Fournier, Erin Gill, Paul Gordon, Emma Griffiths, Jose Hector Galvez Lopez, Darian Hole, Will Hsiao, Jeffrey Joy, Kimia Kamelian, Natalie Knox, Philip Mabon, Finlay Maguire, Tom Matthews, Andrew McArthur, Samir Mechai, Sandrine Moreira, Art Poon, Amos Raphenya, Claire Sevenhuysen, Jared Simpson, Jennifer Tanner, Lauren Tindale, John Tyson, Geoff Winsor, Nolan Woods

***CanCOGeN Ethics and Governance Working Group***

Yann Joly, Fiona Brinkman, Erin Gill, Will Hsiao, Hanshi Liu, Sandrine Moreira, Gary Van Domselaar, Ma'n Zawati, Sarah Savić-Kallesøe

***CanCOGeN Metadata Working Group***

William Hsiao, David Alexander, Zohaib Anwar, Nathalie Bastien, Tim Booth, Guillaume Bourque, Fiona Brinkman, Hughes Charest, Caroline Colijn, Matthew Croxen, Guillaume Desnoyers, Rejean Dion, Damion Dooley, Ana Duggan, Leah Dupasquier, Kerry Dust, Nahuel Fittipaldi, Eleni Galanis, Emma Garlock, Greg German, Erin Gill, Gurinder Gopal, Tom Graefenhan, Morag Graham, Emma Griffiths, Linda Hoang, Naveed Janjua, Jeffrey Joy, Kimia Kamelian, Lev Kearney, Natalie Knox, Theodore Kuschak, Jason LeBlanc, Yan Li, Anna Majer, Adel Malek, Dionne Marcino, Ryan McDonald, David Moore, Celine Nadon, Samir Patel, Natalie Prystajecky, Anoosha Sehar, Claire Sevenhuysen, Garrett Sorensen, Laura Steven, Lori Strudwick, Marsha Taylor, Shane Thiessen, Gary Van Domselaar, Adrian Zetner

***CanCOGeN Research Collaborations Working Group***

Fiona Brinkman, Zohaib Anwar, Marceline Côté, Marc Fiume, Laura Gilbert, Erin Gill, Paul Gordon, Yann Joly, Sandrine Moreira, Samira Mubareka, Natalie Prystajecky, Jennifer Tanner, Gary Van Domselaar, Phot Zahariadis,

***CanCOGeN Sequencing Working Group***

Ioannis Ragoussis, Terrance Snutch, Patryk Aftanas, Matthew Croxen, Hooman Derakhshani, Nahuel Fittipaldi, Morag Graham, Andrew McArthur, Sandrine Moreira, Samira Mubareka, Natalie Prystajecky, Ioannis Ragoussis, Jared Simpson, Michael Surette, John Tyson

***Canadian VirusSeq Data Portal (CVDP) Team***

Guillaume Bourque, Lincoln Stein, Christina Yung, Hanshi Liu, Yann Joly, Adrielle Houweling, William Hsiao, Marc Fiume, David Bujold, Erin Gill, Fiona Brinkman

GenomeCanada

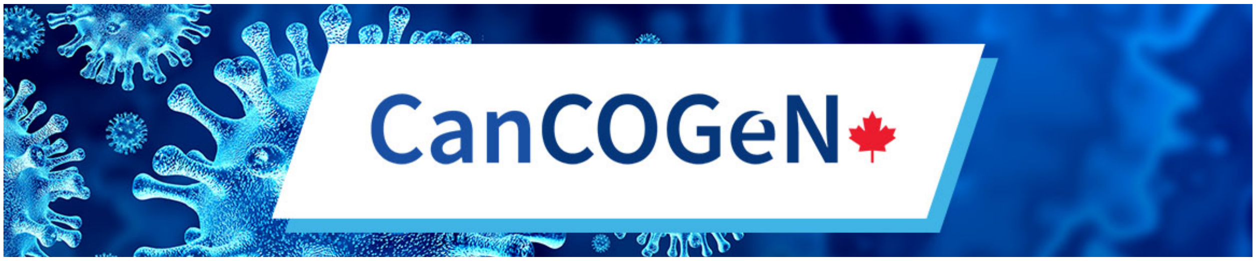


*CanCOGeN Modeling Working Group*

Samira Mubareka, Catalina Lopez-Correa, Erin Gill, Natalie Prystajecky, Beate Sander, Sarah P. Otto, Gary Van Domselaar, Guillaume Polliquin, Sandrine Moreira, Sharmistha Mishra, Caroline Colijn, Koko Agborsangaya, Troy Day, Sandrine Moreira, Micheal Li, Art F.Y. Poon, Jeffrey B. Joy, Marc Brisson, David Earn, Mathieu Maheu-Giroux, Julien Arino, Fiona Brinkman
