## Supplementary Materials for "Effectiveness of Canadian travel restrictions in reducing burden of SARS-CoV-2 variants of concern"

**The PDF file includes:**

Materials and Methods
Supplementary Text
Figs. S1 to S53
Tables S1 to S3
References 1-13

### Materials and Methods

<u>COVID-19 incidence, mortality, and case fatality rates across Canadian provinces and territories</u>
Incidence, mortality, and average case fatality rate (CFR) were compared across provinces and territories to investigate differences in case ascertainment (**Figs. S1, S2**). Although the predominance of Omicron from January 2022 onwards was associated with lower CFR and testing, affecting the final months of the study period, our analyses assume comparable case ascertainment across provinces and over time.

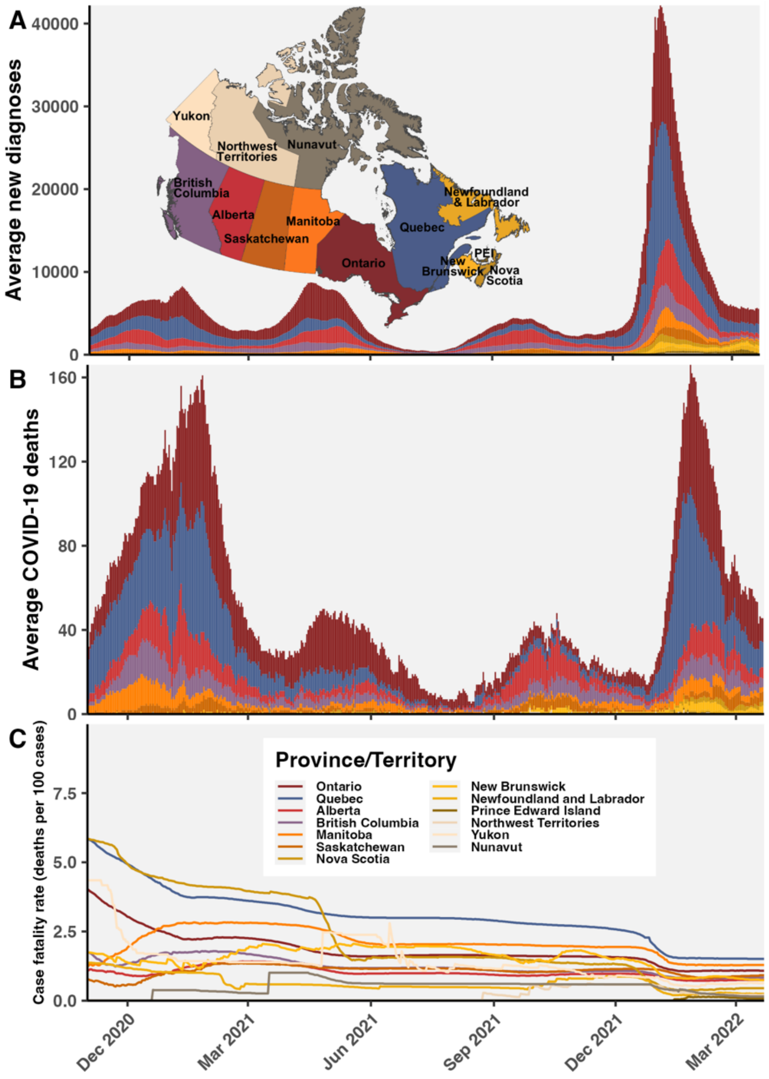


**Fig. S1**. **Comparison of COVID-19 cases, deaths, and case fatality rate across Canadian provinces and territories.** Average daily **(A)** new diagnoses, **(B)** deaths due to COVID-19, and **(C)** case fatality rate over time from November 1, 2020 to March 22, 2022. Averages calculated as right-aligned 7-day rolling averages. Data from Public Health Agency of Canada (*1*).

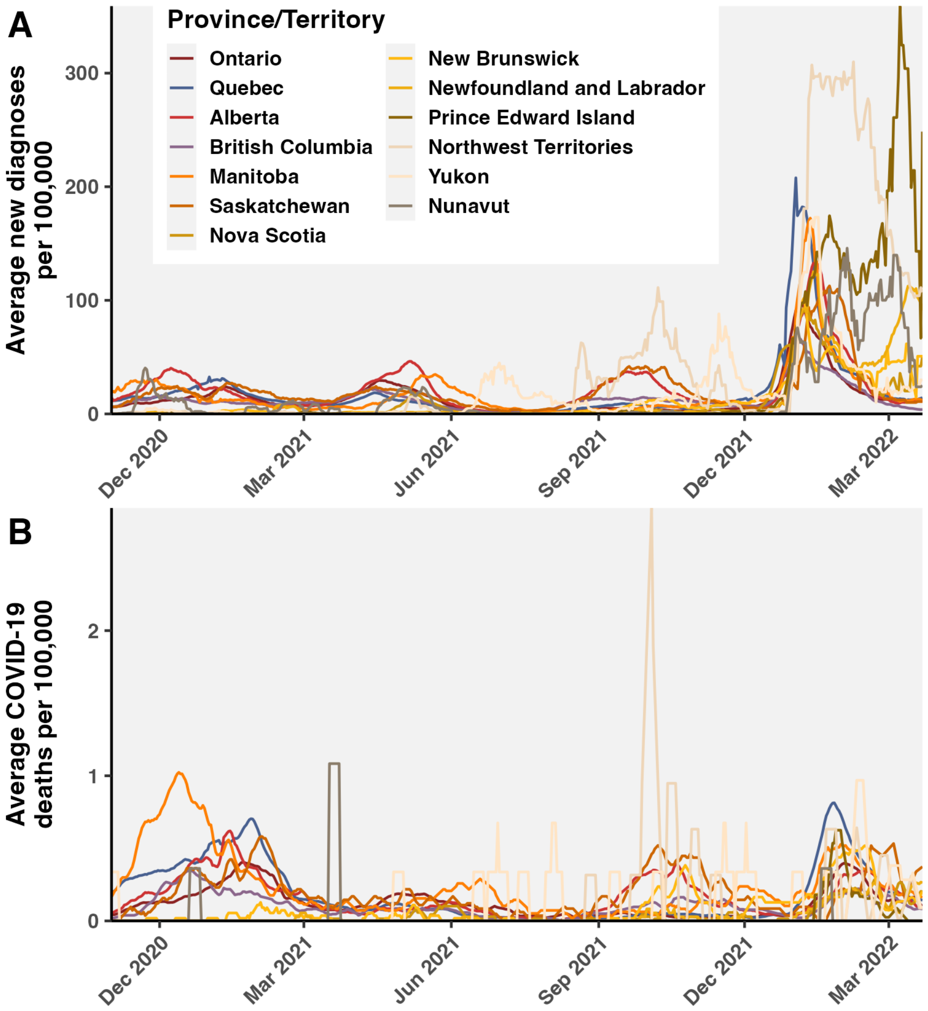


**Fig. S2**. **Population-normalized daily case incidence and death rates. (A)** Average daily new diagnoses per 100,000. **(B)** Average daily deaths from COVID-19 per 100,000. Population sizes for provinces and territories from the Statistics Canada 2021 Census (*2*).

Summary of variants of concern and interest studied

**Table S1. SARS-CoV-2 variants of concern (VOCs) and interest (VOIs) prior to and including early Omicron lineages, ordered by first Canadian sample date.** Lambda, Theta, GH/490R, and BA.3 were not included because they were sampled fewer than 100 times in Canada. First global sample dates were obtained from cov-lineage.org for VOCs and outbreak.info for VOIs; discrepancies between the two were noted. The first Canadian sample date was pulled from clean GISAID sequences, excluding dates preceding first global sample date or incomplete dates.

| Variant type | WHO variant | Pango lineage | GISAID clade | First global detection | First global sample date | First Can. sample date | Notes |
|---|---|---|---|---|---|---|---|
| VOC | Alpha | B.1.1.7; Q.* | GRY | UK | 2020-09-01 | 2020-11-06 | outbreak.info earliest date: 2020-02-07 |
| VOI | Zeta | P.2 | GR/484K.V2 | Brazil | 2020-04-13 | 2020-11-24 | n/a |
| VOC | Gamma | P.1 | GR/501Y.V3 | Brazil | 2020-09-22 | 2020-12-04 | outbreak.info earliest date: 2020-03-15 |
| VOI | Epsilon | B.1.427; B.1.429 | GH/452R.V1 | California, USA | 2020-04-11 | 2020-12-05 | Dates reflect B.1.427. For B.1.429, outbreak.info earliest date: 2020-01-26 |
| VOI | Eta | B.1.525 | G/484K.V3 | Nigeria; UK | 2020-03-25 | 2020-12-14 | Earliest sample from Senegal |
| VOC | Beta | B.1.351 | GH/501Y.V2 | South Africa | 2020-09-01 | 2020-12-19 | outbreak.info earliest date: 2020-02-15. First sample from Zimbabwe |
| VOI | Kappa | B.1.617.1 | G/452R.V3 | India | 2020-03-03 | 2021-02-26 | n/a |
| VOC | Delta | B.1.617.2; AY.* | GK | India | 2021-03-01 | 2021-03-06 | outbreak.info earliest date: 2020-03-27 |
| VOI | Iota | B.1.526 | GH/253G.V1 | New York, USA | 2020-01-28 | 2021-03-17 | n/a |
| VOI | Mu | B.1.621 | GH | Colombia | 2020-12-15 | 2021-06-08 | n/a |
| VOC | Omicron | BA.1; BA.2 | GRA | South Africa; Botswana | 2021-11-08 | 2021-11-04 | outbreak.info earliest date: 2020-05-24. First GISAID sample, BA.1: 2021-11-02 from UK; for BA.1.1: 2021-11-04 from Austria |

**Table S2**. **Summary of inferred number of VOC and VOI introductions by March 22, 2022.** Mean and 95% confidence interval calculated using t-distribution were reported.

| WHO variant | Pango lineage | N introduction | N sublineages | N singletons | % singletons |
|---|---|---|---|---|---|
| Alpha | B.1.1.7; Q.* | 672 (652-693) | 234 (228-240) | 438 (418-459) | 65.2 (64.1-66.2) |
| Zeta | P.2 | 24 (24-24) | 9 (9-9) | 15 (15-15) | 62.5 (62.5-62.5) |
| Gamma | P.1 | 197 (185-210) | 38 (37-40) | 159 (148-170) | 80.7 (80.0-81.0) |
| Epsilon | B.1.427; B.1.429 | 76 (74-78) | 27 (26-27) | 49 (48-51) | 65 (63.8-66.1) |
| Eta | B.1.525 | 62 (59-65) | 24 (23-24) | 38 (36-41) | 61.6 (60.4-62.7) |
| Beta | B.1.351 | 120 (117-124) | 40 (40-41) | 80 (77-83) | 66.7 (65.8-66.9) |
| Kappa | B.1.617.1 | 102 (101-103) | 21 (20-21) | 81 (81-82) | 79.8 (79.3-80.3) |
| Delta | B.1.617.2; AY.* | 1822 (1794-1850) | 537 (521-553) | 1285 (1253-1317) | 70.5 (69.8-71.2) |
| Iota | B.1.526 | 50 (49-52) | 14 (13-14) | 37 (36-38) | 72.6 (71.3-73.9) |
| Mu | B.1.621 | 62 (61-62) | 7 (7-7) | 55 (54-55) | 88.5 (88.1-89.0) |
| Omicron | BA.1 | 3634 (3524-3744) | 615 (596-633) | 3019 (2920-3119) | 83.1 (82.6-83.5) |
| Omicron | BA.1.1 | 5572 (5337-5806) | 1148 (1103-1194) | 4423 (4223-4624) | 79.4 (78.8-79.9) |
| Omicron | BA.2 | 549 (537-565) | 87 (83-92) | 462 (451-473) | 84 (83-85) |

Estimating variant-specific cases

We deconvoluted cases, similarly to (*3*), by multiplying variant frequencies from GISAID by confirmed cases over time to obtain variant-specific cases over time for all of Canada and globally (**Fig. S3**), Canadian provinces (**Fig. S4**), and global regions (**Fig. S6**). Monthly proportional contributions of geographic areas to variant cases were used to inform the subsampling probabilities of sequences from either Canadian provinces (**Fig. S5**) or global regions (**Fig. S7**).

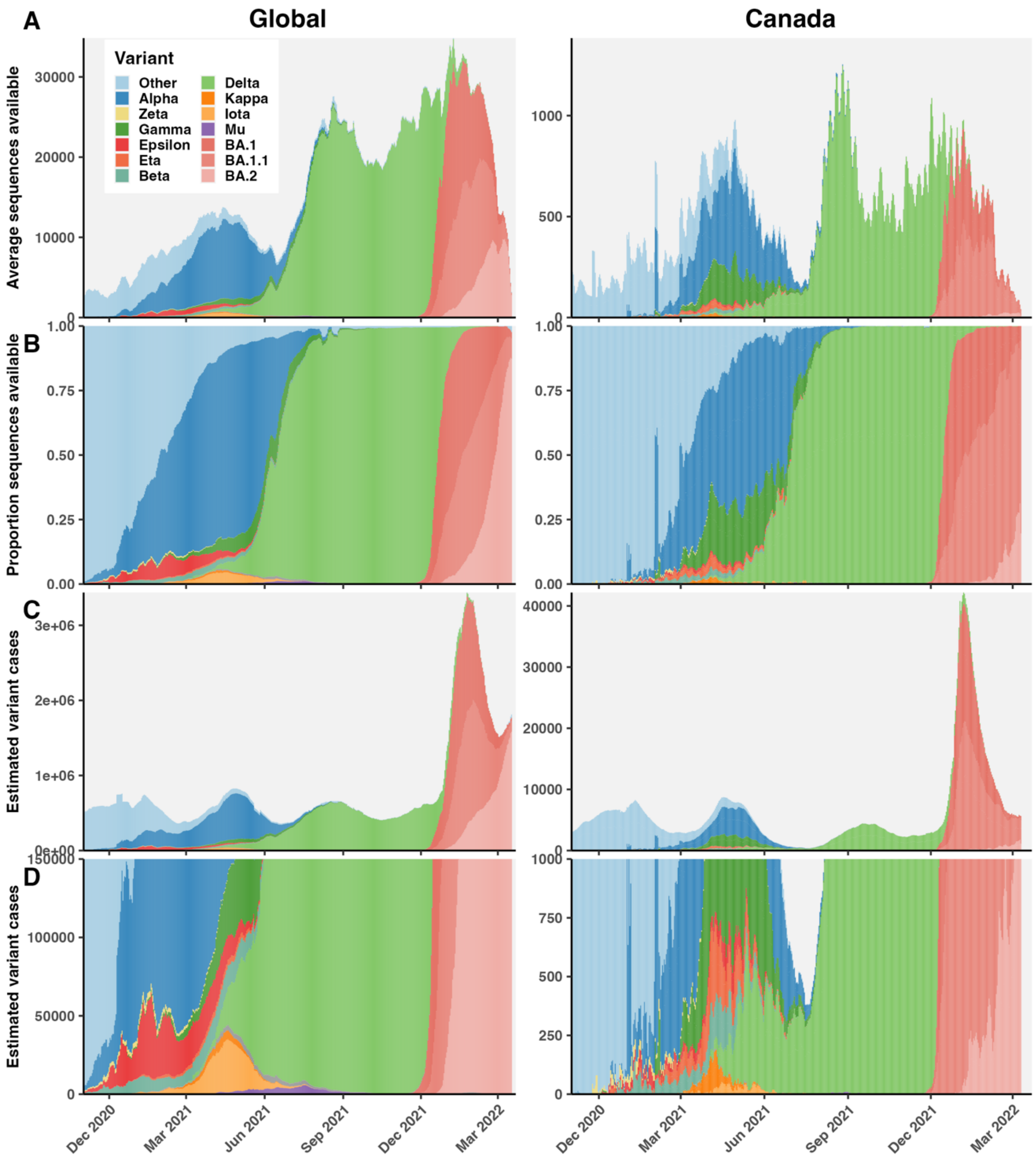


**Fig. S3. Estimation of Canadian and global variant cases as the product of diagnoses and variant frequencies. (A)** Average daily SARS-CoV-2 genome sequences available on GISAID after cleaning, by variant. **(B)** Proportional frequencies of variants among sequences available. **(C)** Estimated variant cases, and **(D)** zoomed in below 1000 cases/day and 150,000 cases/day.

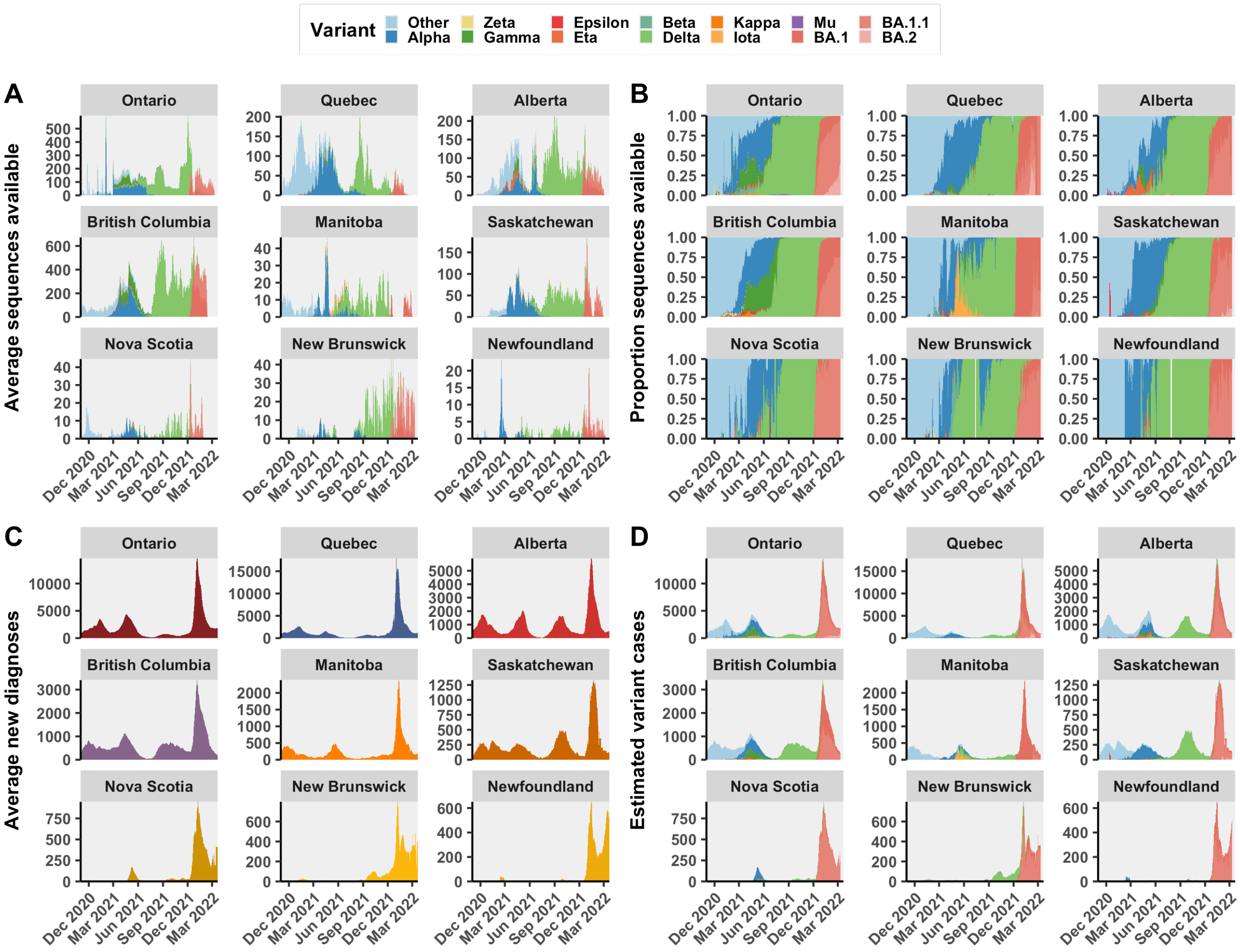


**Fig. S4. Estimation of daily variant cases by Canadian province. (A)** 7-day rolling average number of clean GISAID sequences available per day by province, colored by variant. **(B)** Daily average proportion of variant sequences within each province. If data was incomplete in the past 7 days, the raw count was used. For days with no sequences but with non-zero cases, variant proportions from the previous day with sequences available were carried forward. **(C)** Daily average new diagnoses by province**. (D)** Daily estimated variant-specific cases by province. Only provinces with sequences available on GISAID were shown.

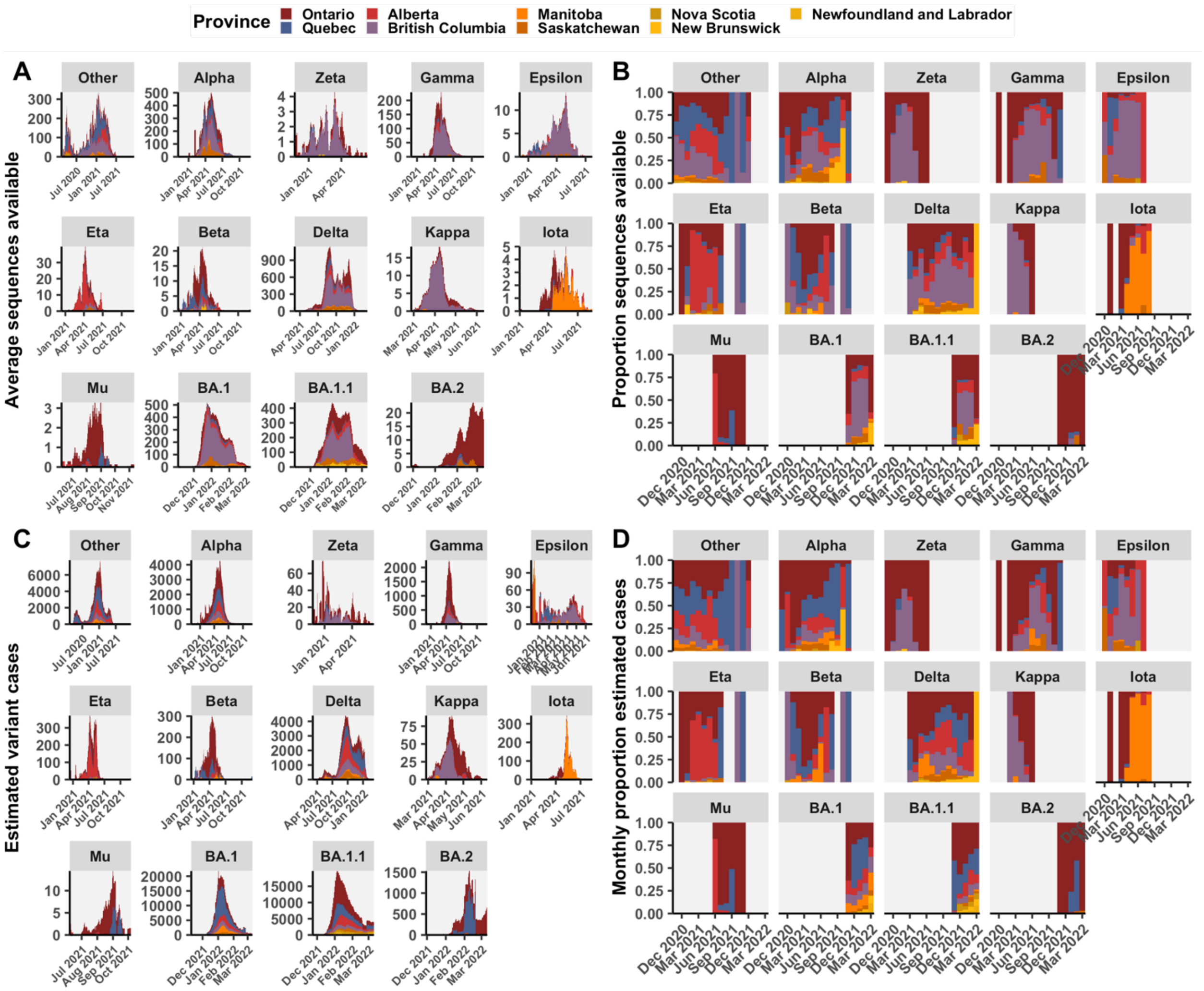


**Fig. S5. Distribution of available sequences and estimated variant cases by Canadian province. (A)** The rolling 7-day average daily and **(B)** monthly proportional sequences available by variant colored by province. **(C)** The estimated daily variant-specific cases and **(D)** monthly proportional contribution of each province to variant-specific cases; the latter informs the sampling probability of sequences in the analysis.

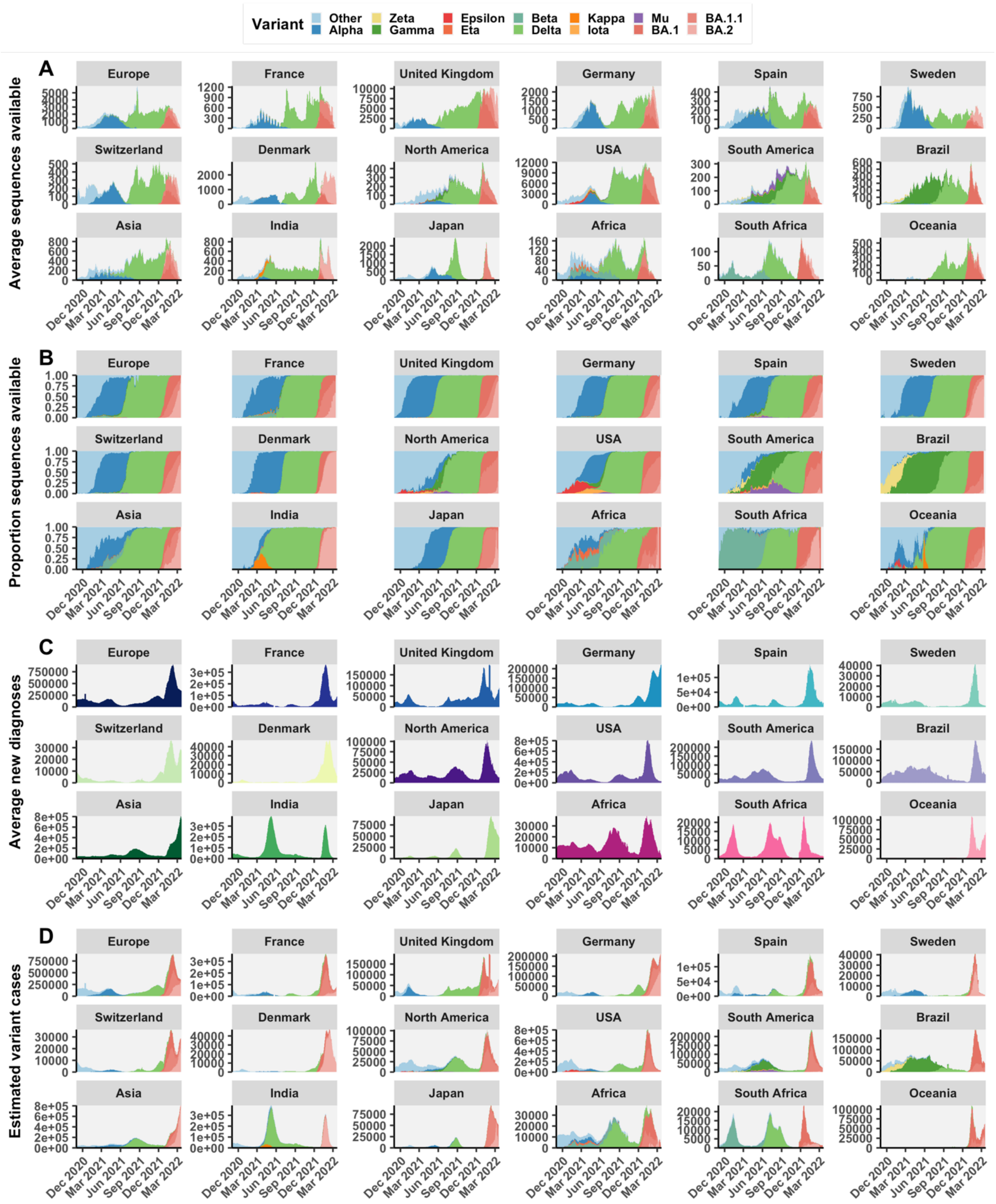


**Fig. S6. Estimation of daily variant cases by global region. (A)** Daily average clean GISAID sequences available by region, colored by variant. **(B)** Daily average proportion of sequences contributed by each variant to regional totals. If data was incomplete in the past 7 days, the raw count was used. For days with zero sequences and non-zero cases, variant proportions from the previous day with sequences available were carried forward. **(C)** Daily average new diagnoses by global region. **(D)** Daily estimated variant-specific cases by region.

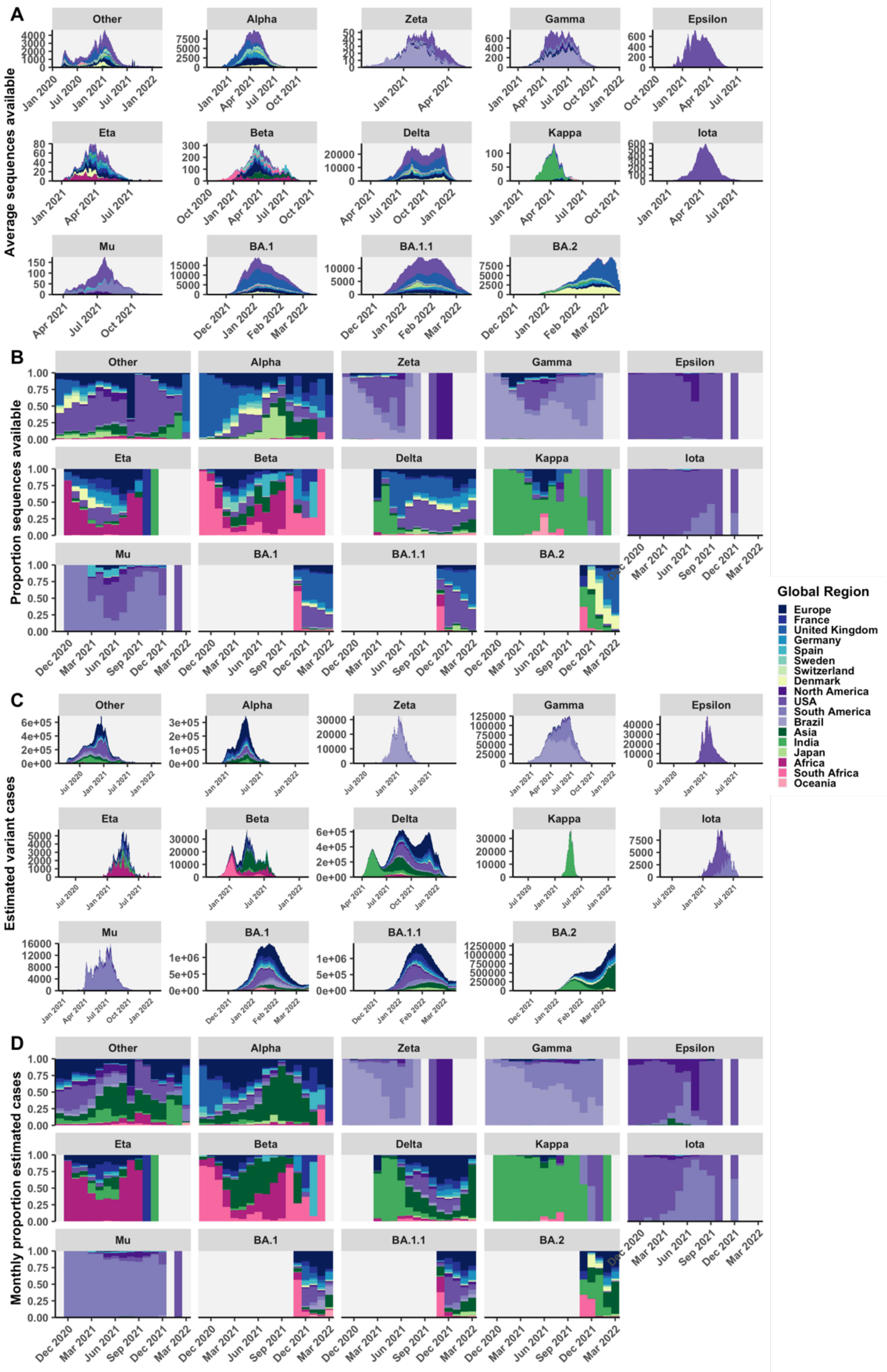


**Fig. S7. Distribution of available sequences and estimated variant cases, by global region. (A)** The rolling 7-day average daily and **(B)** monthly proportional sequences available by variant colored by region. **(C)** The estimated daily variant-specific cases and **(D)** monthly proportional contribution of each region to variant-specific cases; the latter informs the sampling probability of sequences in the analysis.

**Table S3. The cumulative number of estimated variant cases globally and in Canada up to the day of data download March 22, 2022, relative to clean sequences available on GISAID.** Cumulative cases by variant were summed across global regions or Canadian provinces. Excludes sequences with unidentified lineage, Theta, Lambda, and GH/490R.

| | Global | | | Canada | | |
|---|---|---|---|---|---|---|
| **Variant** | **Cumulative cases** | **Clean sequences** | **Sequence per case** | **Cumulative cases** | **Clean sequences** | **Sequence per case** |
| Other | 109,632,102 | 942,487 | 0.009 | 930,569 | 58,001 | 0.062 |
| Alpha | 31,471,326 | 1,065,890 | 0.034 | 307,680 | 42,596 | 0.138 |
| Zeta | 2,520,842 | 4,721 | 0.002 | 2,906 | 244 | 0.084 |
| Gamma | 15,750,416 | 98,939 | 0.006 | 93,762 | 13,488 | 0.144 |
| Epsilon | 3,223,780 | 63,369 | 0.02 | 5,591 | 752 | 0.135 |
| Eta | 445,556 | 6,733 | 0.015 | 19,067 | 1,800 | 0.094 |
| Beta | 4,290,486 | 35,606 | 0.008 | 18,326 | 1,414 | 0.077 |
| Delta | 111,501,622 | 3,921,060 | 0.035 | 500,617 | 111,192 | 0.222 |
| Kappa | 1,638,294 | 6,621 | 0.004 | 2,838 | 481 | 0.169 |
| Iota | 917,808 | 38,216 | 0.042 | 8,325 | 302 | 0.036 |
| Mu | 1,369,309 | 13,140 | 0.01 | 405 | 126 | 0.311 |
| BA.1 | 65,751,679 | 903,272 | 0.014 | 630,408 | 21,242 | 0.034 |
| BA.1.1 | 69,901,000 | 791,147 | 0.011 | 741,437 | 21,920 | 0.03 |
| BA.2 | 42,513,492 | 404,349 | 0.01 | 38,009 | 817 | 0.021 |

<u>Subsampling strategy</u>

In our previous study of the first two waves of SARS-CoV-2 in Canada in 2020 and early 2021, we subsampled sequences from provinces or countries with probabilities reflecting geographies' monthly proportional contributions to cases (*4*). The more data were subsampled, the more bias was reduced for previously overrepresented geographies; however, when less than 50% of the data were from Canada, our ability to identify introductions was lessened. An even split of 50% global and 50% Canadian sequences per subsample identified the most introductions. The current analysis of variant dynamics differs in a few fundamental ways. First, the available number of sequences for this period of the pandemic is a magnitude larger than what was available previously. Previously, we were sampling from ~60,000 Canadian sequences and ~500,000 global sequences; the current analysis draws from ~300,000 Canadian sequences and >5,000,000 global sequences. Another difference is the emergence of variants that are not distinguished in the case data, which necessitated that we estimate the variant-specific case distribution over time (described above). Geographies' monthly contributions to variant-specific cases informed sequences' sampling probabilities, downweighing geographies with relatively more sequences per cases. Further, in this analysis we applied temporally distributed sampling of sequences per month that mimicked the distribution of variant cases (**Fig. S5, S7**). A simple algorithm was written in R to augment sampling of sparse months where relatively few cases were identified (**Fig. S8**).

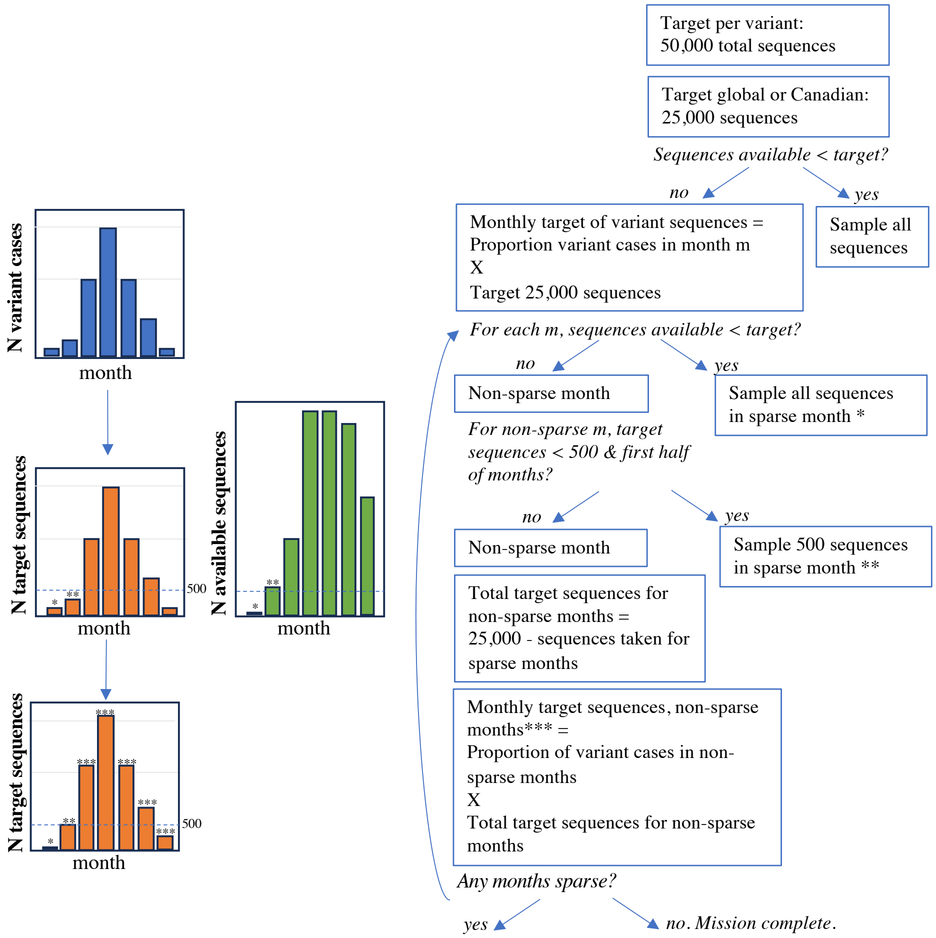


**Fig. S8. Graphical depiction of algorithm for temporally distributed sequence subsampling.** Sequences subsampled mimic the distribution of variant cases over time with augmented representation of sparse early months.

Subsampling improved the correlations of monthly cases to sequences among Canadian provinces and global regions (**Fig. S12, S13**). For variants like Delta, where sequences were abundant but differentially collected and submitted (widely different sequence per case ratio), subsampling improved the correlation for Canadian provinces (Pearson's correlation coefficients: from 0.58 to 0.69) and global regions (from 0.47 to 0.76). Similarly, Alpha correlation improved for global (0.60 to 0.90) and Canadian (0.65 to 0.77) following subsampling. The correlation between monthly sequences sampled and monthly cases was improved more with temporally distributed sampling than uniform sampling for Alpha and Delta variants.

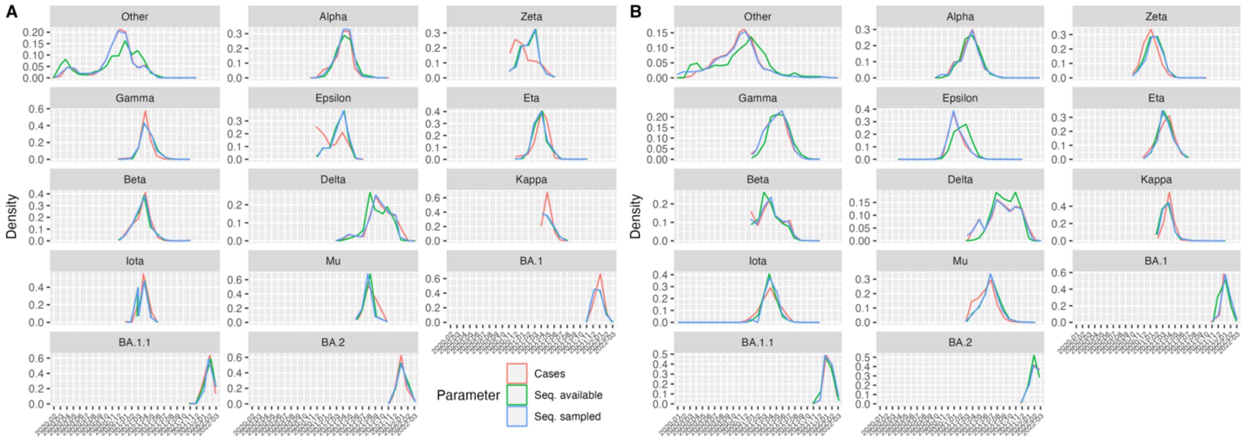


**Fig. S9. The relative monthly distributions of cases and sequences**. Proportions of cases, available sequences (seq. available), and sequences taken (seq. sampled) using the temporally distributed sampling strategy for each variant in a representative subsample for **(A)** Canadian and **(B)** global datasets.

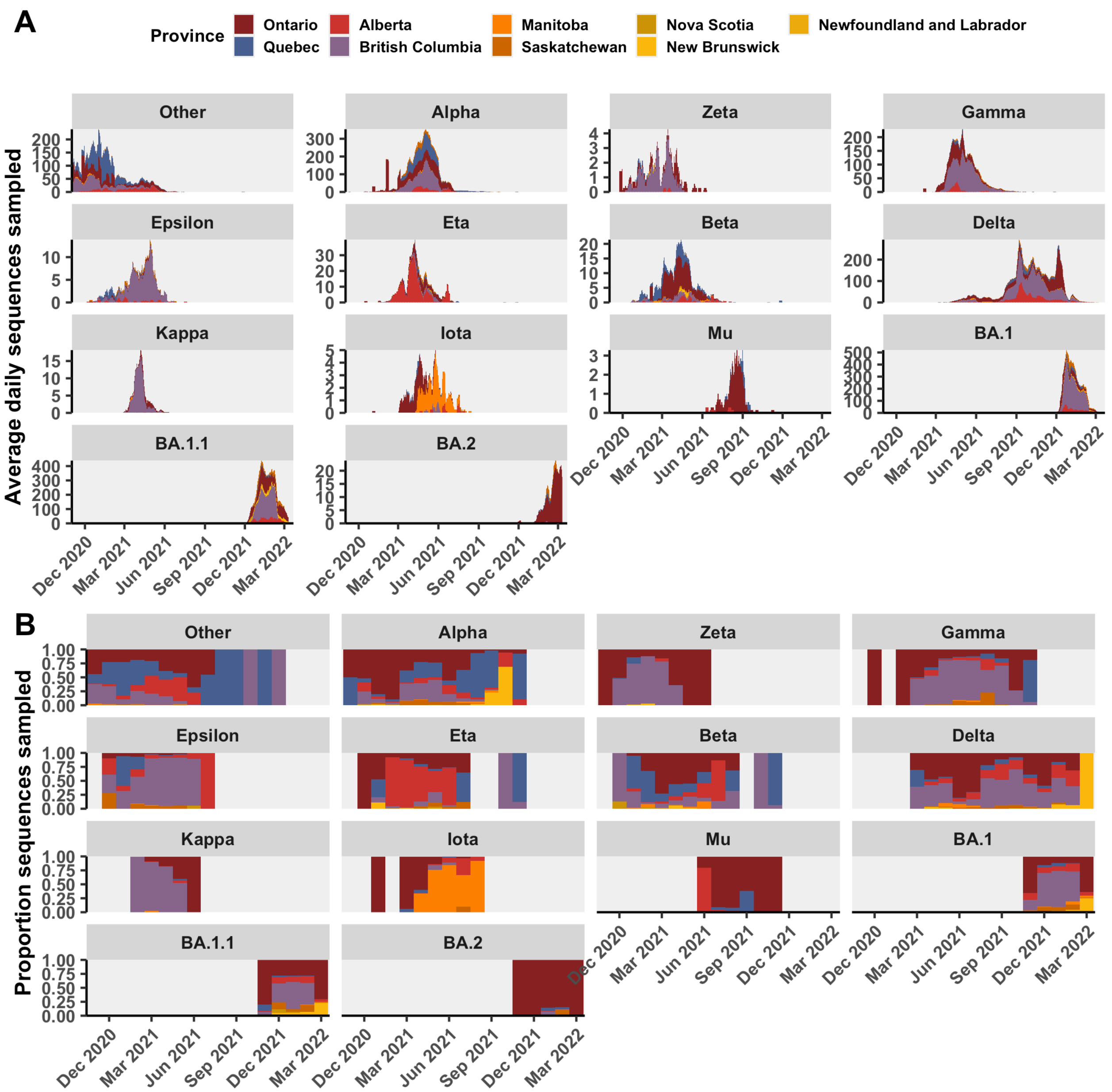


**Fig. S10**. **Average daily and monthly proportion of sequences sampled for each Canadian province for a representative subsample, grouped by variant.**

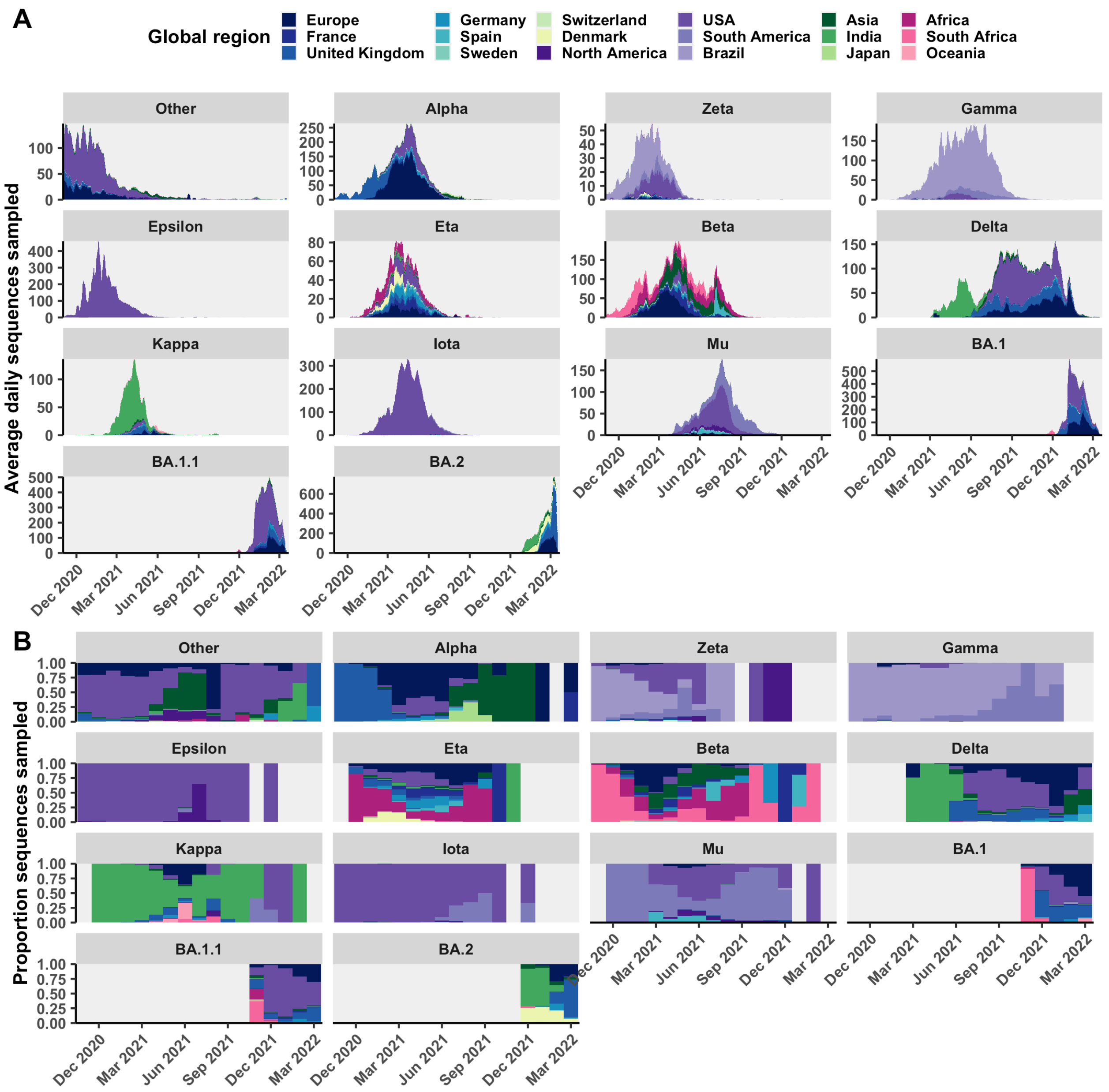


**Fig. S11. Average daily and monthly propotion of sequences sampled for each global region for a representative subsample, grouped by variant.**

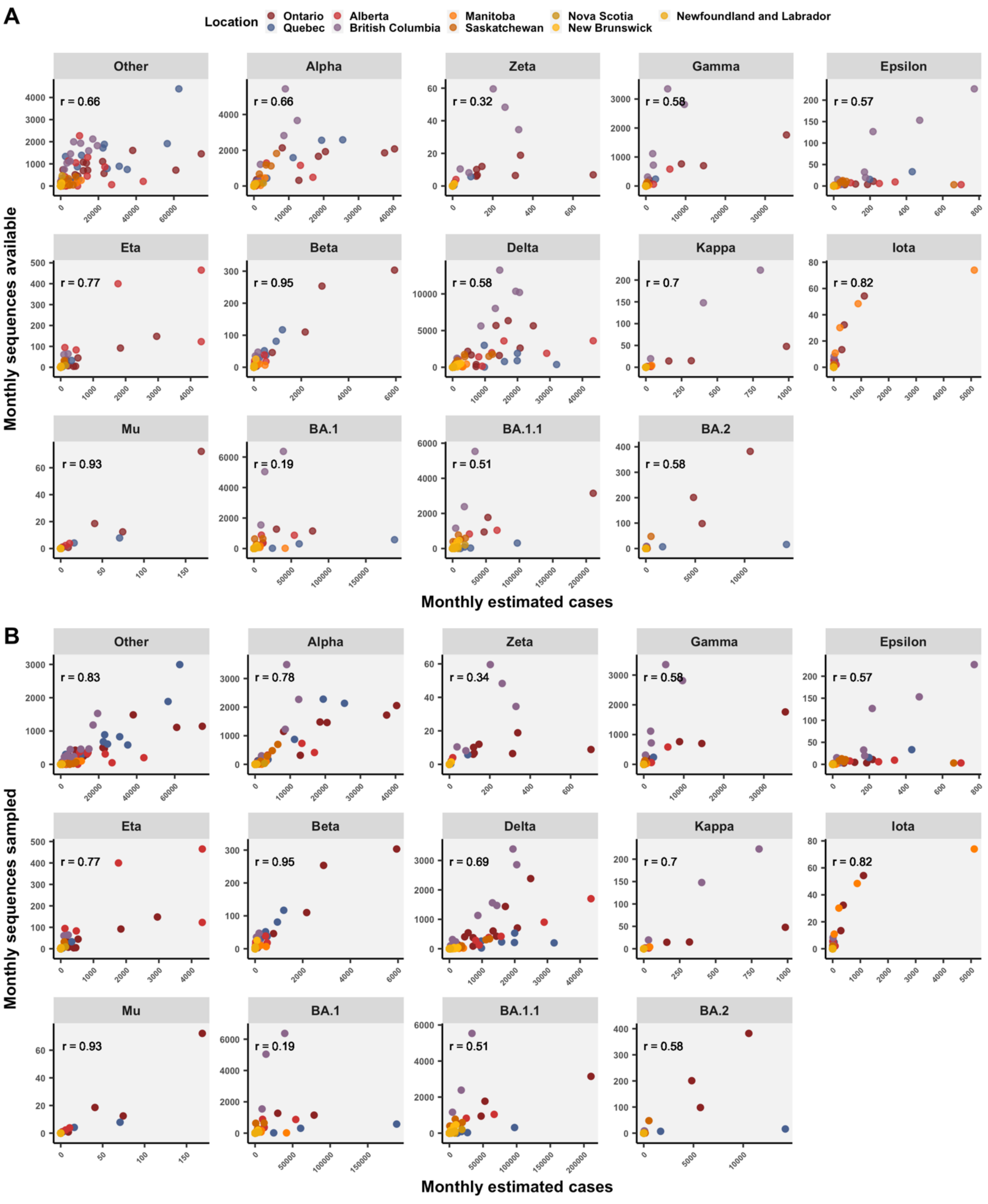


**Fig. S12. Canadian sequence representation**. The relationship between Canadian monthly estimated variant cases and **(A)** monthly sequences available (pre-subsample) for provinces, or **(B)** monthly sequences sampled. Relationships summarized by Pearson's correlation coefficients.

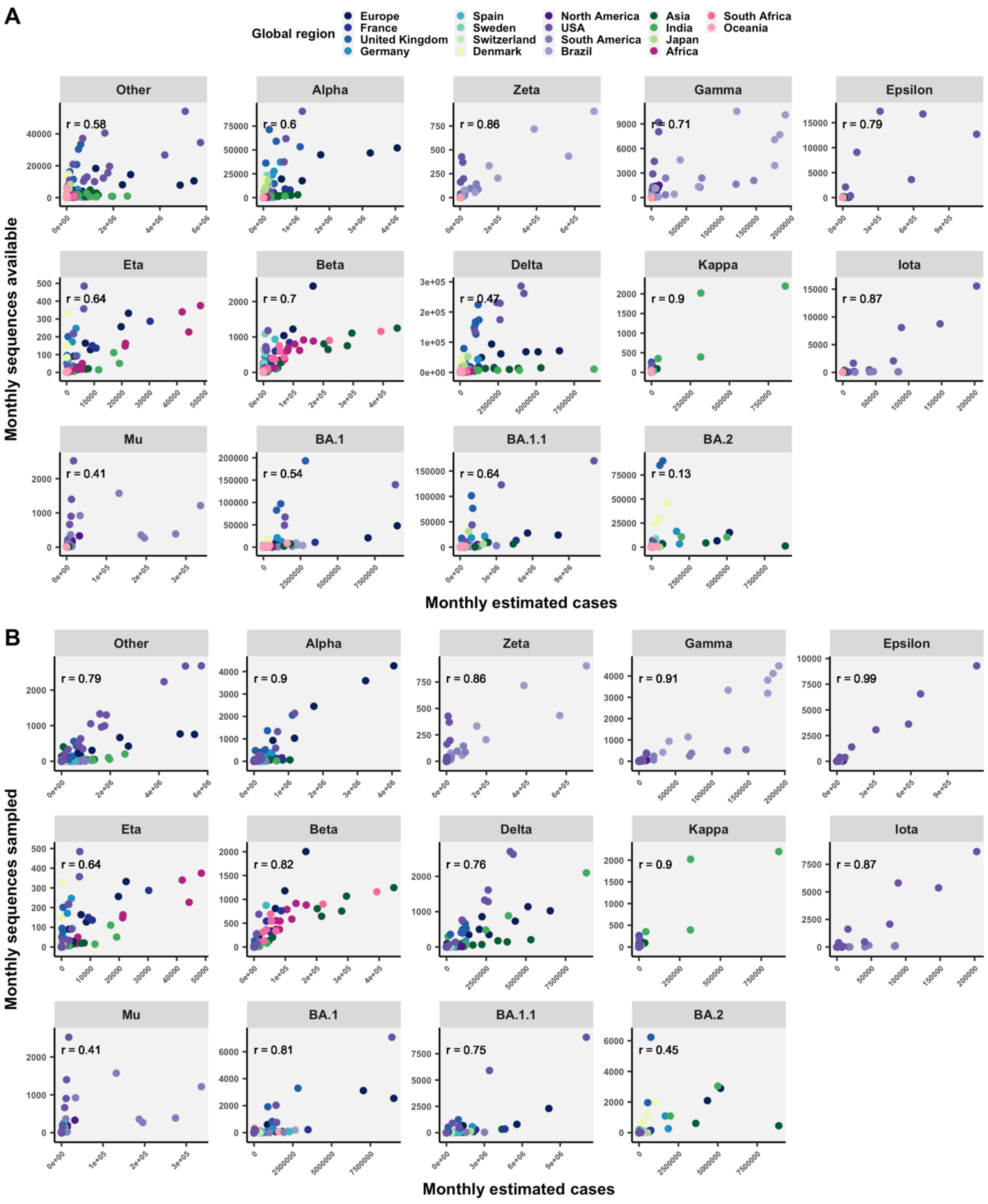


**Fig. S13**. **Global sequence representation.** The correlation between global monthly estimated variant cases and **(A)** monthly sequences available (pre-subsample) for global regions, or **(B)** monthly sequences sampled. Correlations summarized by Pearson's correlation coefficients.

Modelling travellers averted using historical travel data

Statistics Canada data on international travel arrivals was used to estimate likely travel volume in the absence of travel restrictions for all VOCs (*5*). For the Delta variant, for example, we analyzed travel volume from India to Canada from 2018 to 2023**.** Average daily travellers by month revealed seasonality (**Fig. S14C**), with higher-than-average travel volume from India to Canada in April and May, and to a lesser extent June to August, in 2018 and 2019 (pre-COVID-19), and 2022 (low to negligible COVID-19 restrictions). Seasonality was quantified as the deviation of monthly compared to annual average daily travellers - either as the difference in the monthly and annual means (diff.means) or the ratio of monthly to annual means (ratio.means) (**Fig. S15**). Across 2018, 2019 and 2022, we summarized the mean monthly deviation from the annual trend (**Fig. S15C**). Expected travel in the absence of the travel ban considering seasonality was estimated as the annual mean travel volume, adjusted by monthly diff.means or ratio.means (**Fig. S16**).

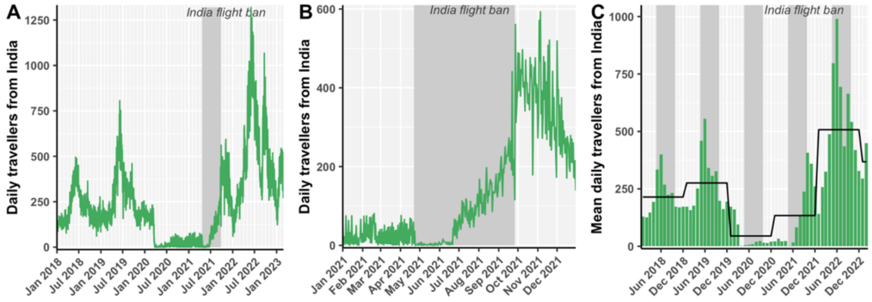


**Fig. S14**. **Observed daily travellers from India to Canada (A) from January 2018 to 2023, (B) in 2021. (C)** Bar chart of monthly mean daily travellers from India, where the black line is the mean daily travelers in each calendar year. Highlighted the time of year of the flight ban (April 22 - Sept 26) in adjacent years.

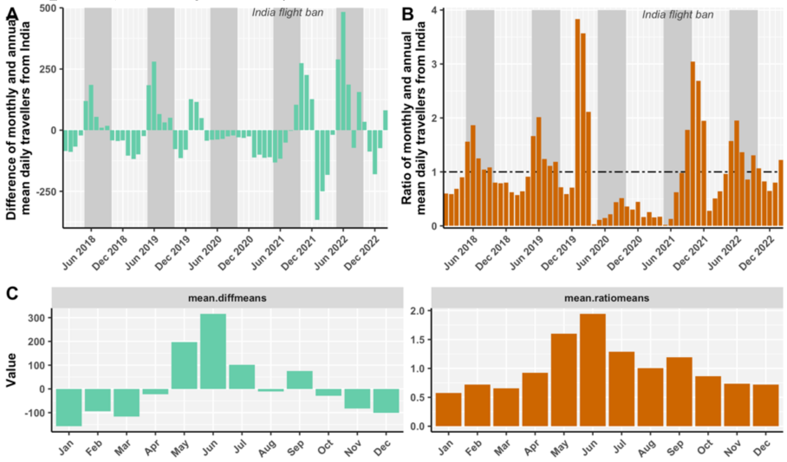


**Fig. S15**. **Seasonal variation in travel volume from India summarized by deviation of monthly compared to annual average daily travellers**. **(A)** Difference between monthly and annual mean daily travelers ('diffmeans'). **(B)** Ratio between monthly and annual mean daily travelers ('ratiomeans'). **(C)** The mean monthly deviation from the annual trends in 2018, 2019, and 2022.

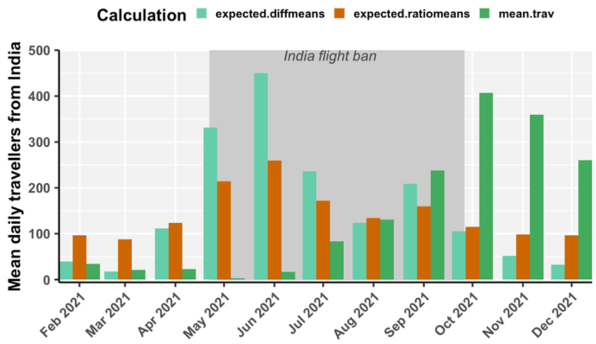


**Fig. S16. Observed (mean.trav) and expected mean daily travellers from India in the absence of the COVID-19-related flight ban based on seasonality trends**. Expected.diffmeans was calculated as the average daily travel in 2021 plus the mean diff.mean. Expected.ratiomeans was calculated as the average daily travel in 2021 times the mean ratio.mean.

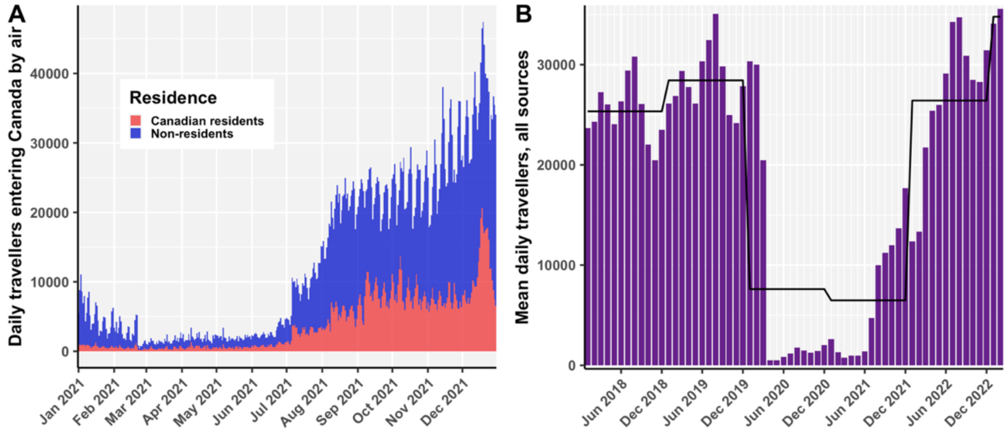


**Fig. S17. Observed daily air travel volume into Canada from all sources**. **(A)** Daily travellers entering Canada by air, colored by Canadian residents vs. non-residents in 2021. **(B)** Monthly mean daily travellers from 2018 to 2022 from all sources, with black line for annual means.

With seasonal fluctuations accounted for, we next considered the epochal changes in travel volume from all sources into Canada to incorporate broad changes in travel patterns due to other COVID-19 restrictions, which may have confounded our estimates (**Fig. S17**). Non-resident visitors and Canadian-resident visitors daily travel volume was steadily low in 2021 until July 5, when the requirement for fully vaccinated air travelers eligible to enter Canada to quarantine for 3 days in a hotel and provide a test after 8 days ended (*6*). In September 2021, fully vaccinated foreign nationals were allowed to enter Canada for non-essential reasons, corresponding to a further increase in travel volume. How did observed air travel in 2021 deviate from what we expected based on seasonality?

Towards this, we applied a similar strategy as conducted above for a singular source (India), but for all sources. We calculated the difference and ratio between expected daily travellers (resident and non-resident, all sources) based on seasonal trends in 2018, 2019, and 2022, and the observed daily travellers in 2021 from all sources (**Fig. S18**). The monthly ratio of means is an adjustment factor that takes into consideration broad travel patterns affected by other policies and willingness to travel into Canada in 2021. Monthly adjustment factors were used to correct our expectations of monthly travel in the absence of flight bans in India (**Fig. S19**). This adjusted expectation considers seasonality in travel from India to Canada based on historical data, average travel from India to Canada in 2021, and broad trends in air travel from all sources in 2021. Finally, we converted this to a daily expectation from monthly average by fitting a spline to the monthly averages (**Fig. S20**). The number of travellers averted was calculated by comparing expected (no ban) to observed (with ban).

For Delta and the India flight ban, 5,147 travellers from India were averted via the restrictions (17,958 expected during the ban, and 12,811 observed). This process was repeated for UK (Alpha), Brazil (Gamma), South Africa (Beta), and Africa (Omicron) (**Fig. S21**). We estimated 724 travellers from the UK were averted during the Alpha intervention; only 3 travellers from South Africa were averted during the Beta intervention; 383 travellers were averted from Brazil over both Gamma-related restriction periods; 1681 travellers from Africa were averted during the Omicron-related travel restriction. Then, we tried to incorporate expected travel volume as a confounding factor in the model between sublineage importation rate and variant cases in focal source.

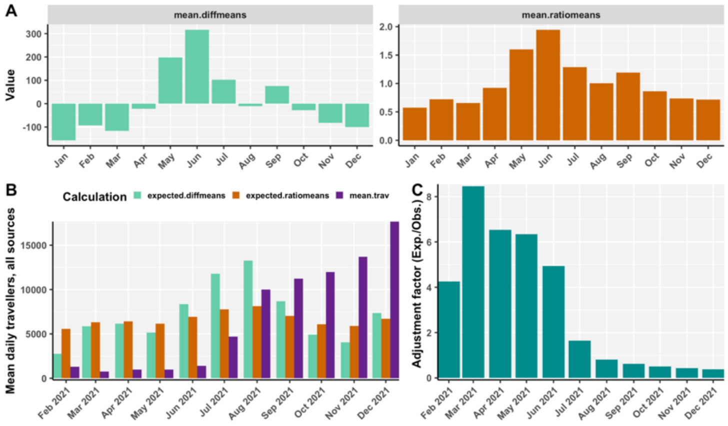


**Fig. S18**. **Seasonal variation in travel volume from all sources**. **(A)** Mean monthly deviation from the annual trend in 2018, 2019, and 2022, summarized as the mean (across years) of the difference in means or ratio of means for each month. **(B)** Expected daily travellers from all sources versus observed (mean.trav), using difference in means or ratio of means. **(C)** Adjustment factor reflecting epochal changes in overall travel was calculated for each month in 2021 as the ratio of expected (ratio means) over the observed mean daily travellers.

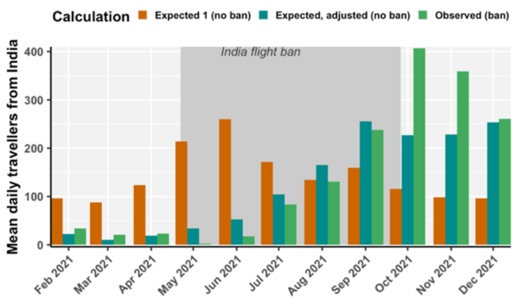


**Fig. S19. Adjusted expectations of monthly travel volume from India in the absence of flight bans.** 'Expected 1 (no ban)' is based on seasonality in India travel and 'Expected, adjusted (no ban)' is based on seasonality and epochal travel changes from all sources.

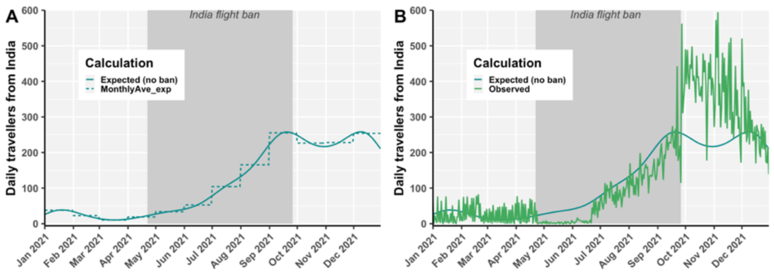


**Fig. S20. Splines convert monthly average expected travellers to daily average travellers.**

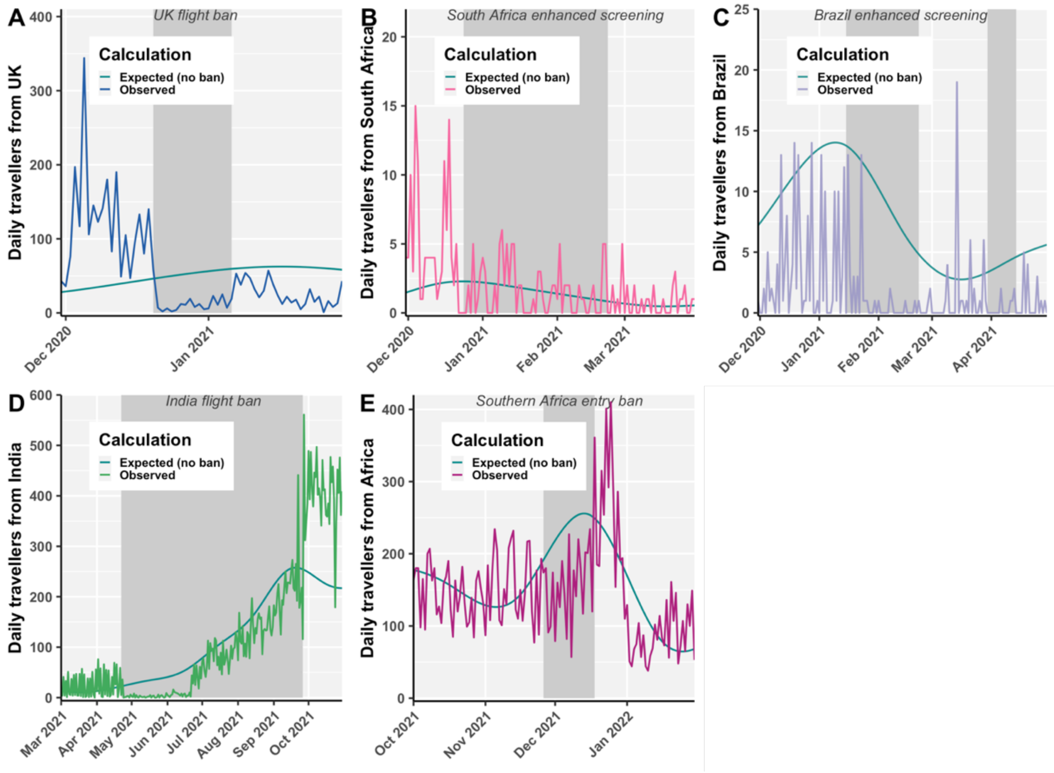


**Fig. S21. Expected (in the absence of restrictions) versus observed daily travellers from focal countries affected by COVID-19 travel restrictions imposed in Canada. (A)** Alpha from the UK, **(B)** Beta from South Africa**, (C)** Gamma from Brazil, **(D)** Delta from India, and **(E)** Omicron from southern Africa (presented for all of Africa).

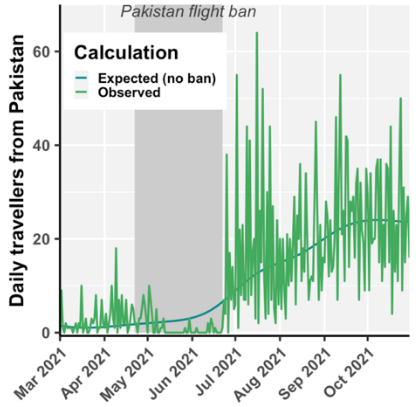


**Fig. S22. Expected versus observed daily travellers from Pakistan to Canada in the context of the two-month flight ban countering the Delta variant.**

<u>Modelling importations and cases averted</u>

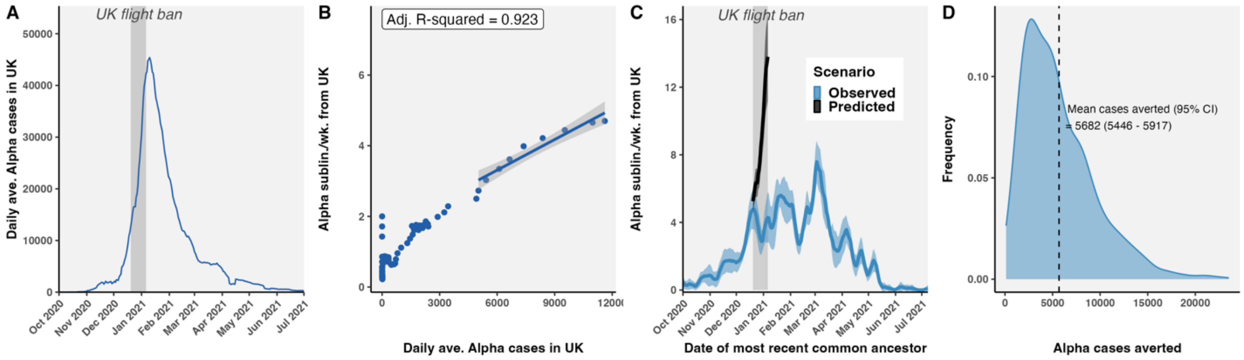


**Fig. S23**. **Alpha sublineages and descendant cases averted. (A) Estimated average daily Alpha cases in the UK. (B)** Linear model fit to Alpha sublineages per week from UK versus average daily Alpha cases in UK prior to restrictions, subset to greater than 5000 cases. **(C)** Observed and predicted Alpha sublineages per week from the UK. **(D)** Total descendant cases averted across all averted sublineages

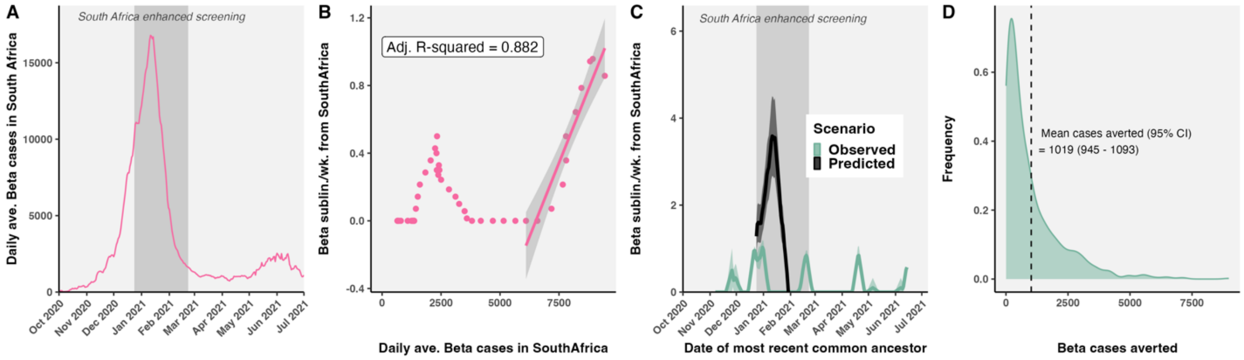


**Fig. S24**. **Beta sublineages and descendant cases averted. (A) Estimated average daily Beta cases in South Africa. (B)** Linear model fit to Beta sublineages per week from South Africa versus average daily Beta cases in South Africa prior to restrictions, subset to greater than 6000 cases. **(C)** Observed and predicted Beta sublineages per week from South Africa. **(D)** Total descendant cases across all averted sublineages.

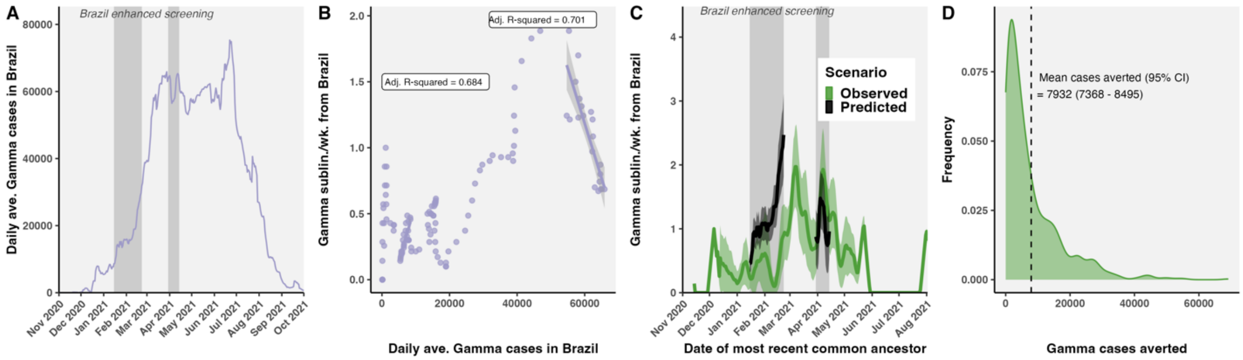


**Fig. S25. Gamma sublineages and descendant cases averted for two interventions. (A) Estimated average daily Gamma cases in Brazil. (B)** Linear model fit to Gamma sublineages per week from Brazil versus average daily Gamma cases in Brazil prior to restrictions, subset to greater than 5000 cases for the first intervention period**. (C)** Observed and predicted Gamma sublineages per week from Brazil. **(D)** Total descendant cases across all averted sublineages.

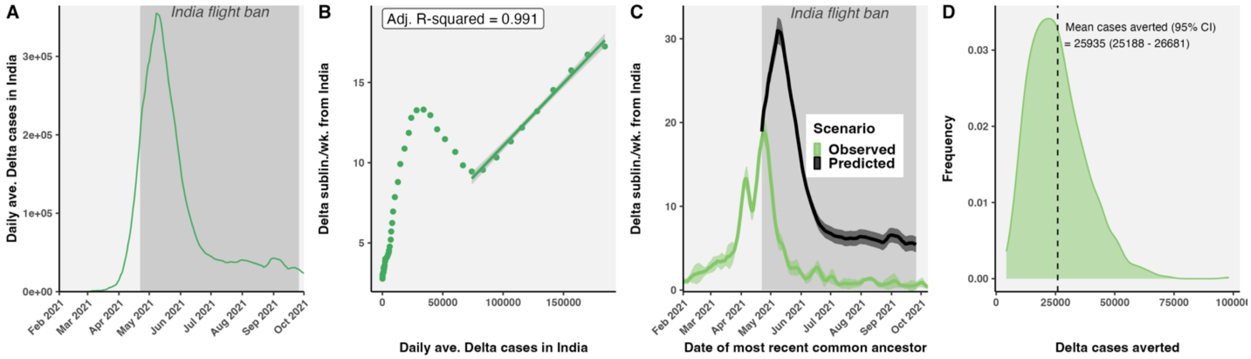


**Fig. S26. Delta sublineages and descendant cases averted. (A) Estimated average daily Delta cases in India. (B)** Linear model fit to Delta sublineages per week from India versus average daily Delta cases in India prior to restrictions, subset to greater than 70,000 cases. **(C)** Observed and predicted Delta sublineages per week from India. **(D)** Total descendant cases averted across all averted sublineages.

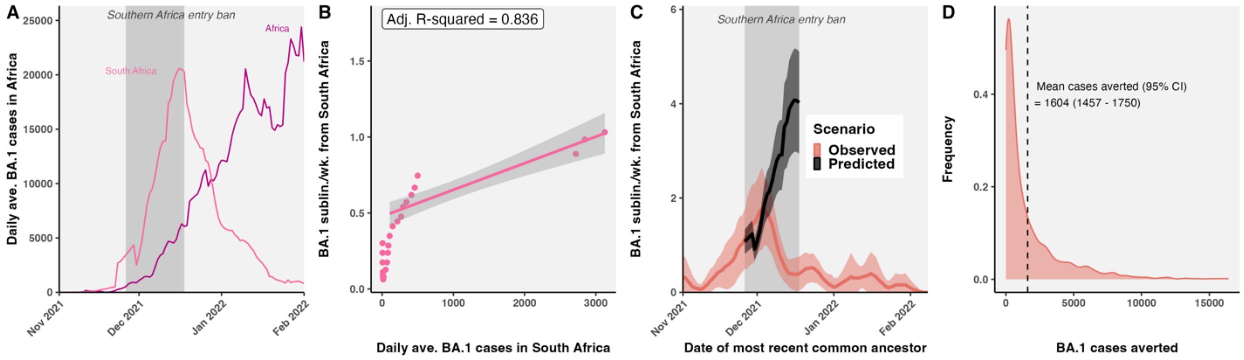


**Fig. S27. Omicron BA.1 sublineages and descendant cases averted. (A) Estimated average daily Omicron cases in South Africa and other African nations. (B)** Linear model fit to Omicron sublineages per week from South Africa versus average daily Omicron cases in South Africa prior to restrictions, subset to greater than 25 cases**. (C)** Observed and predicted Omicron sublineages per week from South Africa. **(D)** Total descendant cases averted across all averted sublineages.

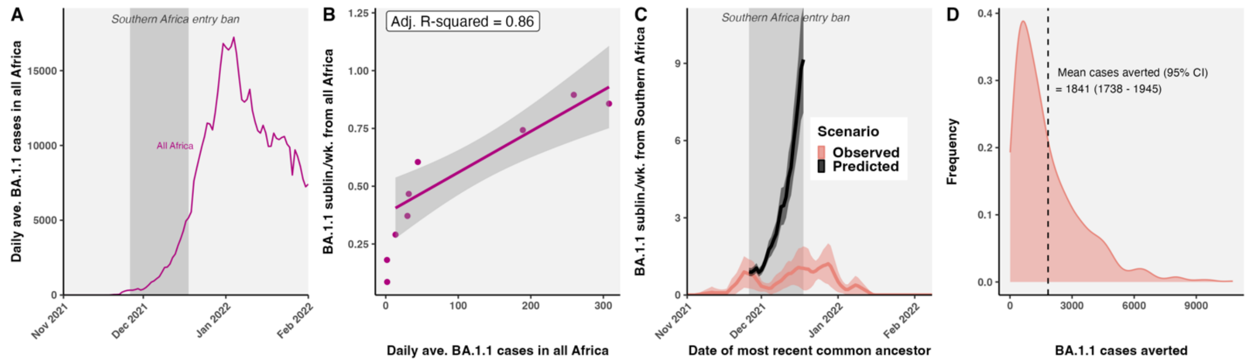


**Fig. S28. Omicron BA.1.1 sublineages and descendant cases averted. (A) Estimated average daily Omicron cases in all of Africa**. **(B)** Linear model fit to Omicron sublineages per week from Africa versus average daily Omicron cases in Africa prior to restrictions, subset to greater than 10 cases**. (C)** Observed and predicted Omicron sublineages per week, adjusted by travel, from South Africa over time. Model goodness-of-fit was improved by adjusting for travel volume. **(D)** Total descendant cases averted across all averted sublineages.

### Supplementary Text

Timeline of Canadian COVID-19 variant era restrictions

This timeline summarizes COVID-19 travel restrictions in Canada in the VOC era from November 2020 up to March 2022, including national entry requirements, epidemiological events, and variant-specific travel restrictions (**Fig. S29**). Data was collated from a combination of resources, including the Canadian Institute for Health Information (CIHI) (*7*), news releases from the Government of Canada (cited throughout), and a timeline of Canada Border Services Agency Border Measures (*8*). CIHI maintains a comprehensive list of federal, provincial and territorial government interventions, announcements and other measures to reduce the spread of and improve the health outcomes related to COVID-19. In the main text, we focus only upon variant-specific travel restrictions for Alpha, Beta, Gamma, Delta, and Omicron (**Fig. 29C**).

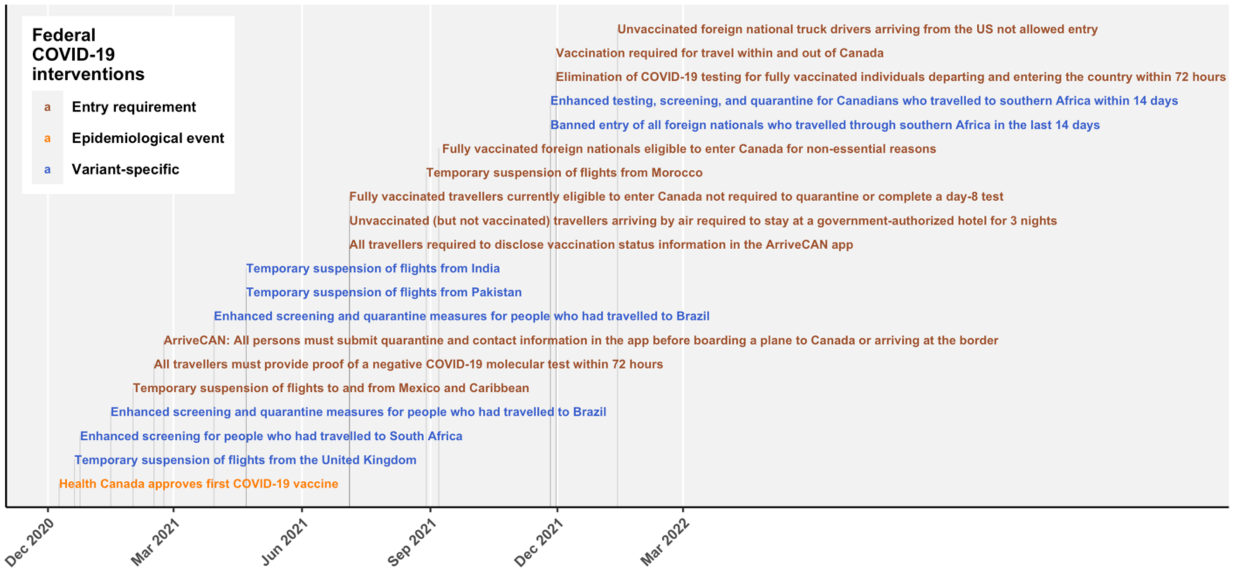


**Fig. S29.** Timeline of Canadian federal COVID-19 travel restrictions, including entry requirements and variant-specific enhanced screening or entry bans, between November 2020 and March 2022.

A synopsis of the timing of entry requirements, epidemiological events, and variant-specific measures in Canada towards reducing the importation and burden of VOCs.

- December 9, 2020 – Health Canada approves first COVID-19 vaccine.
- December 20, 2020 – January 6, 2021: Flight ban for flights arriving from the United Kingdom in response to the Alpha variant.
- December 24, 2020 – February 22, 2021, enhanced screening for arrivals from South Africa in response to the Beta variant.
- As of February 15, 2021, foreign nationals arriving from the US must provide proof of a valid COVID-19 molecular test from less than 72 hours before or prior positive COVID-19 molecular test taken between 14-90 days before entry into Canada (*9*).
- As of February 22, 2021, all persons arriving by air or land must submit quarantine and contact information electronically on the ArriveCAN app before boarding a plane to Canada or arriving at the border with limited exceptions (*9*).
- On June 21, 2021 it was announced that beginning July 5, 2021, only unvaccinated travellers arriving by air would be required to stay at a government-authorized hotel for three nights, but not vaccinated travellers (*6*).
- Effective July 5, 2021, fully vaccinated travellers eligible to enter Canada will not be required to quarantine while they await their on-arrival test result or complete a day-8 test. At any time after entry to Canada, if a fully vaccinated traveller tests positive or are exposed, local public health requirements, including quarantine or isolation, apply.
- On November 19, 2021, Transport Canada announced, effective of November 30, 2021, the elimination of COVID-19 testing for fully vaccinated individuals departing and

entering the country within 72 hours; expanded the list of accepted vaccines for the purpose of travel; valid COVID-19 tests no longer accepted as an alternative to vaccination for travel within Canada; new requirements for all exempt essential travellers, including the mandate to identify their vaccination status in ArriveCAN. Vaccination still required for travel within and out of Canada (*10*).

- January 13, 2022, effective January 15, PHAC announced Canadian truck drivers who were not fully vaccinated would not be denied entry into Canada, but unvaccinated or partially vaccinated foreign national truck drivers coming to Canada from the US by land would not be allowed entry.
- February 14, 2022, federal government invoked the Emergency Act to disperse 2022 Freedom Convoy - a protest of truckers and other anti-COVID-19 mandate supporters who occupied the capital for three weeks and blocked at least two major international borders.
- February 15, 2022, as of February 28, there would be easing of on-arrival testing for fully-vaccinated travellers with random testing and no quarantine required while awaiting test results (*11*).
- Study period end; data download date: March 22, 2022.
- April 1, 2022, fully vaccinated travellers were no longer required to provide a pre-entry COVID-19 test result to enter Canada by air, land or water; unvaccinated must continue to provide a COVID-19 test result (*12*).
- April 25, 2022, border measures eased: unvaccinated/partially vaccinated children accompanied by a fully vaccinated guardian no longer required to complete a pre-entry COVID-19 test; fully vaccinated travellers no longer need to provide a quarantine plan; fully vaccinated travellers no longer have to wear a mask in public spaces, monitor or report symptoms, quarantine if a traveller in their group shows symptoms, or maintain a list of close contacts and locations visited. Passengers still required to use ArriveCAN (*13*).
- October 1, 2022, the arriveCAN program ended and all entry requirements were dissolved. Vaccination, negative test, arriveCAN, or masking no longer required for all travellers entering Canada by air, land or marine mode.

### SARS-CoV-2 VOC and VOI sublineage and singleton dynamics in Canada

#### *Alpha (B.1.1.7)*

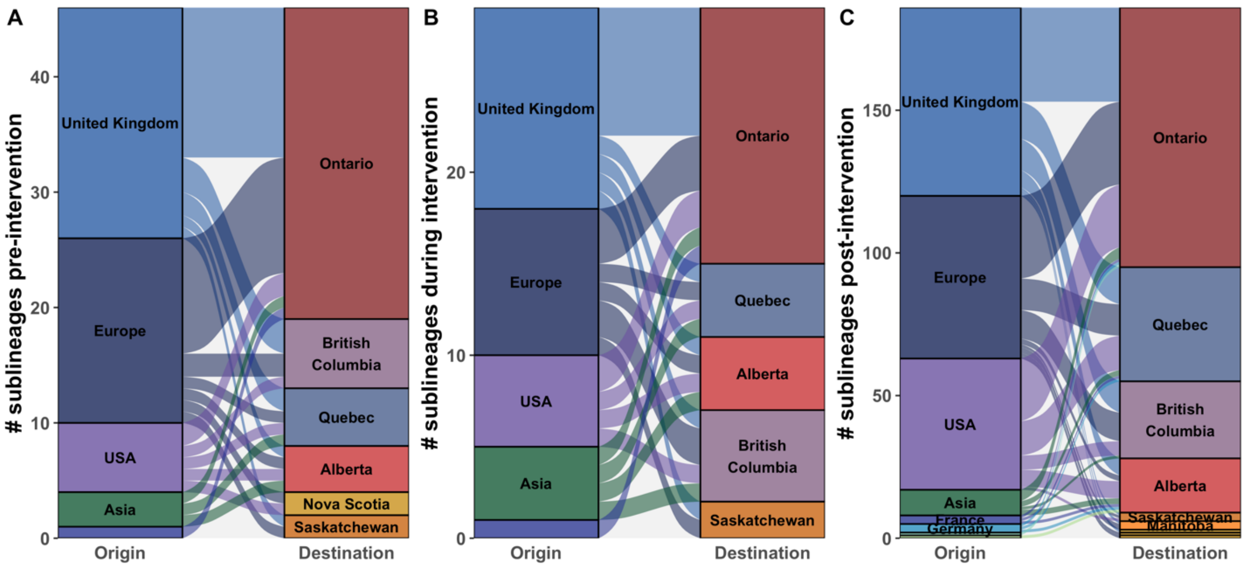


**Fig. S30**. Relative contributions of global origins and Canadian destinations to Alpha variant. sublineage importations into Canada **(A)** before, **(B)** during, and **(C)** after the UK flight ban.

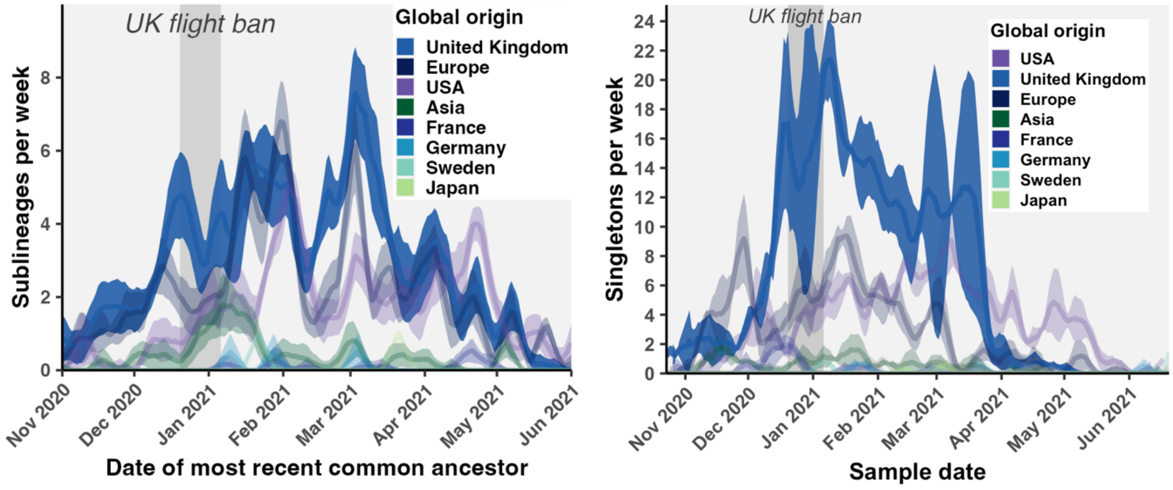


**Fig. S31. Alpha sublineages and singletons imported per week colored by global origins, in the context of the UK flight ban.** Ribbons denote 95% confidence intervals across 10 bootstraps.

#### *Zeta (P.2)*

The second variant to be identified in Canada on November 24, 2020 was Zeta (P.2, alias: B.1.1.28.2), which was closely related to the Gamma variant and initially identified in Brazil as early as April 2020. The majority of Zeta sublineages and singletons introduced to Canada were imported from Brazil, with smaller contributions from the USA and Europe (**Fig. S32**).

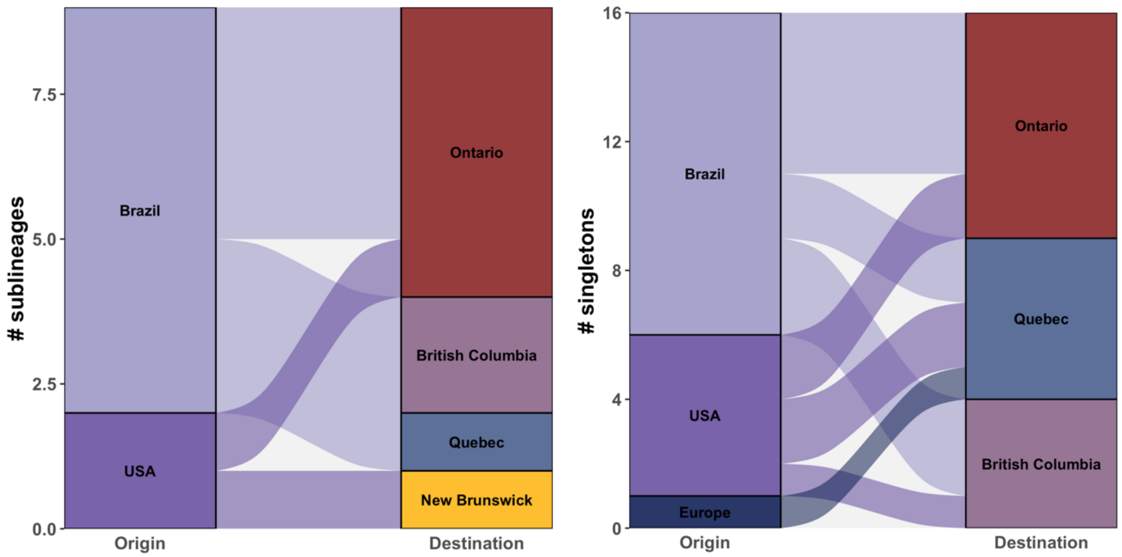


**Fig. S32. Zeta (P.2) sublineage and singleton flows.**

***Gamma (P.1)***

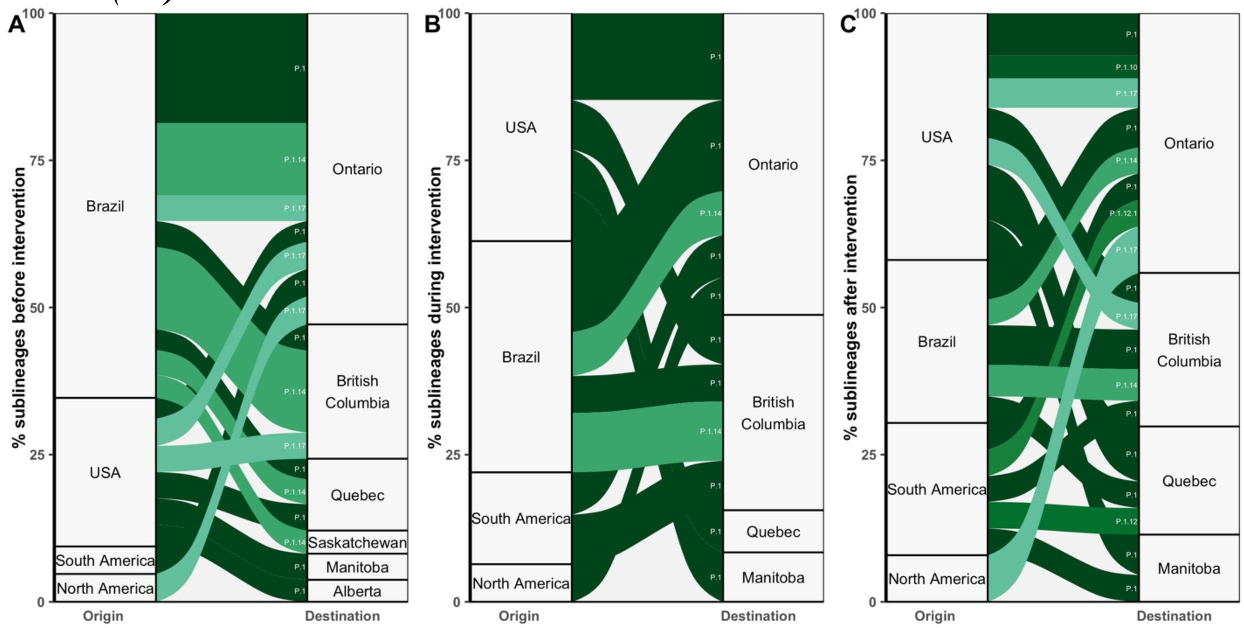


**Fig. S33. Relative flows of Gamma sublineages from global origins to Canadian provinces before, during, and after the Brazil enhanced screening restriction**. Flows are grouped by Pango lineage.

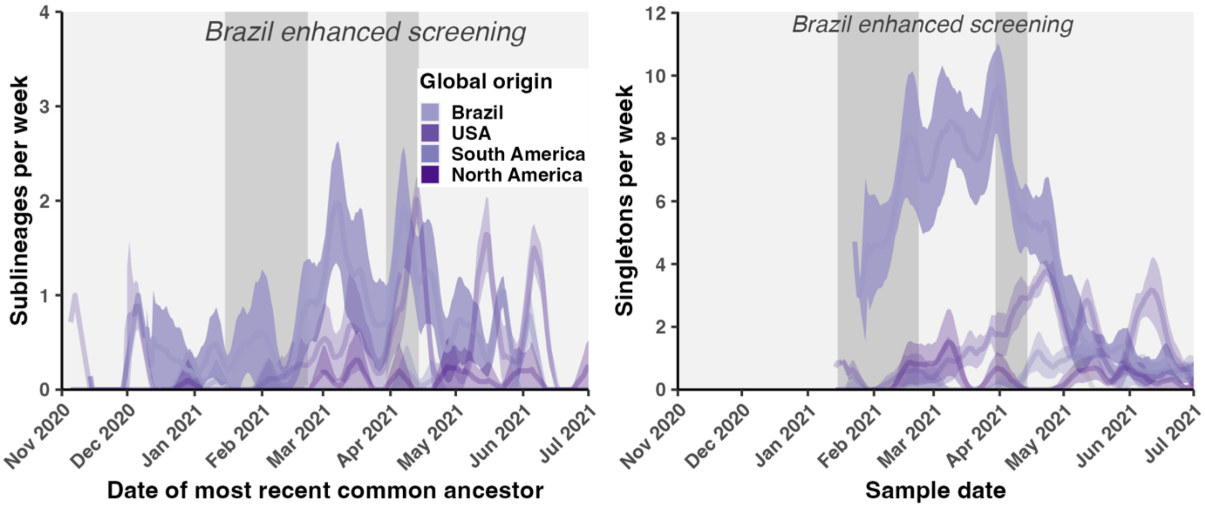


**Fig. S34. Gamma sublineages and singletons introduced per week over time.**

***Epsilon (B.1.427, B.1.429)***

Epsilon consists of Pango lineages, B.1.429 and B.1.427, both of which were first detected in California, USA, with earliest sample date for B.1.427 in April 2020 and B.1.429 in January 2020 according to cov-lineages.org (**Table S1**). The first detection in Canada was on December 5 2020. Our analysis suggests that Canada had 21 (20-22) B.1.429 and 6 (5-6) B.1.427 sublineages introduced between October 9, 2020 and April 12, 2021 (**Figs. S35-36**). All sublineages were inferred to have USA origins across all bootstraps.

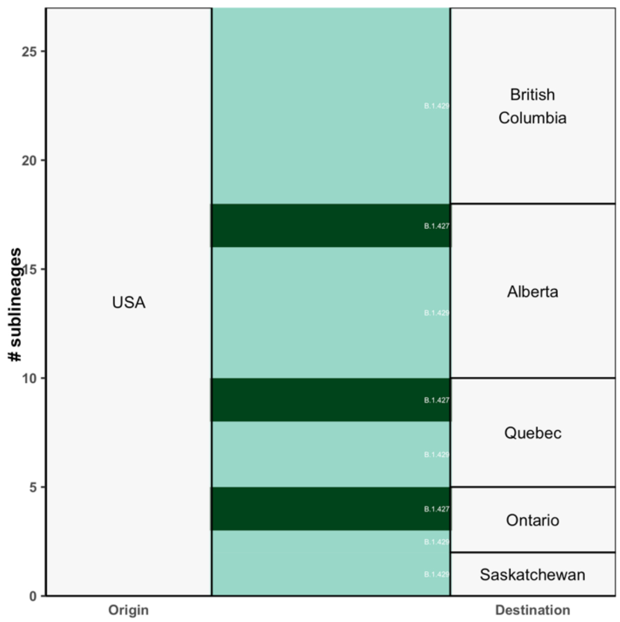


**Fig. S35**. **Epsilon sublineage flows grouped by Pango lineage.**

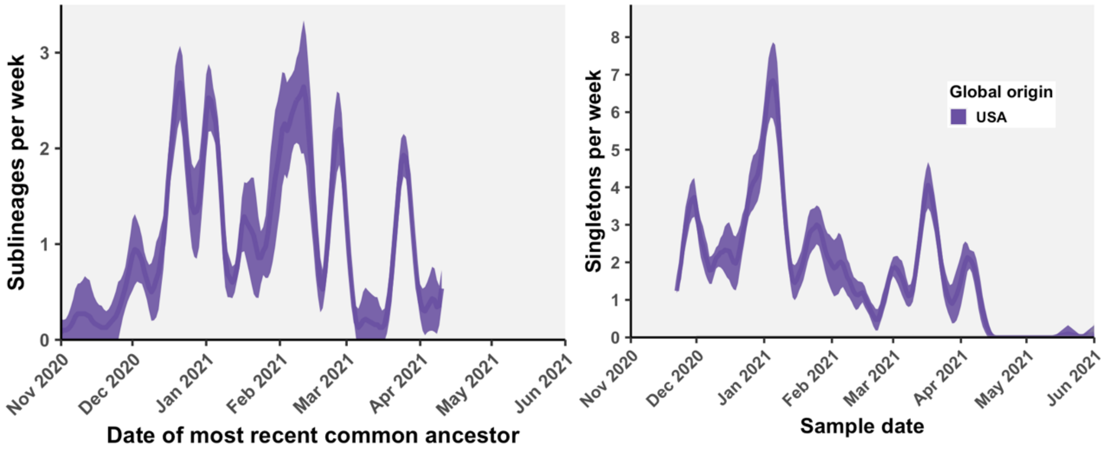


**Fig. S36. Epsilon sublineages and singletons per week by global region of origin.**

#### ***Eta (B.1.525)***

The Eta variant was first detected in the UK and Nigeria (**Table S1**). We estimated there were 24 (23 - 24) Eta sublineages imported into Canada between October 2020 and July 2021, resulting in 1762 (1759 - 1764) descendants sampled in Canada. There were additionally 38 (36 - 41) Eta singletons introduced into Canada, accounting for 62% (60-63) of all Eta introductions.

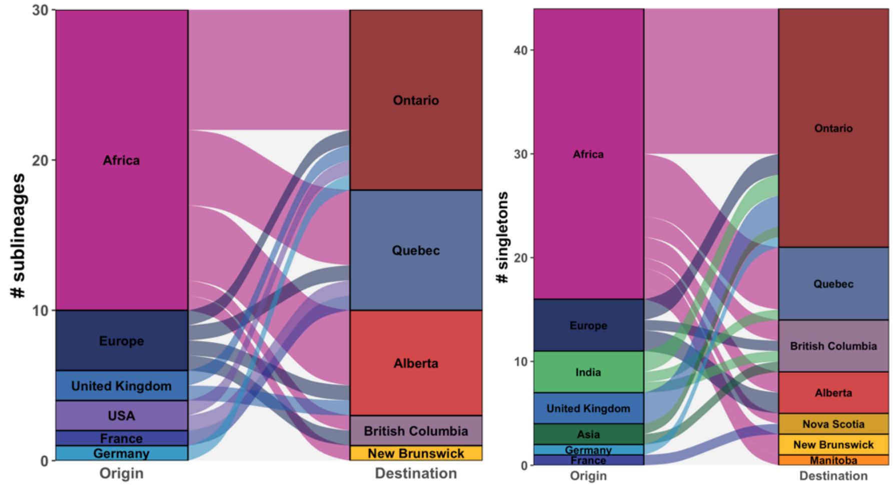


**Fig. S37**. **Eta sublineage and singleton flows.**

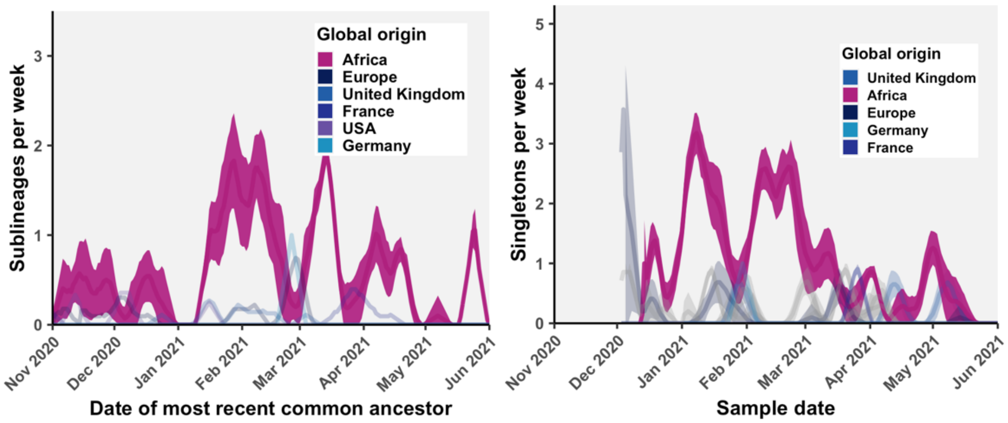


**Fig. S38. Eta sublineages and singletons over time.**

***Beta (B.1.351)***

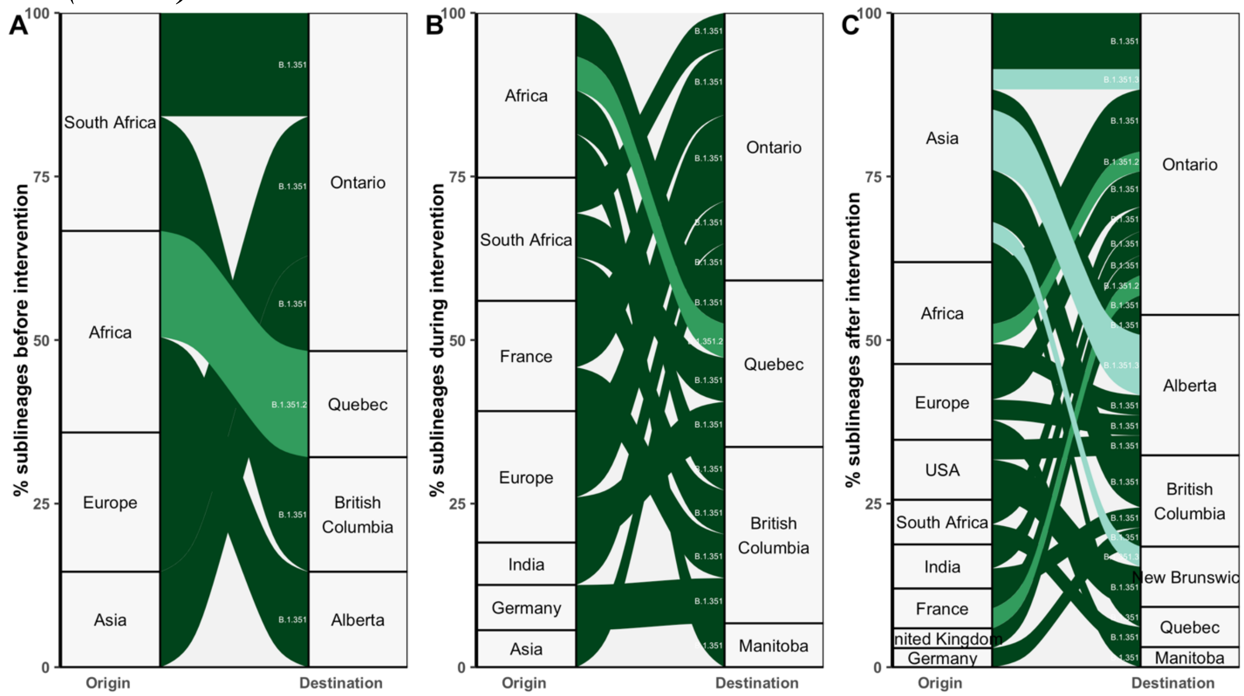


**Fig. S39. Beta sublineage flows, grouped by Pango lineage, before, during, and after the enhanced screening period for South Africa.**

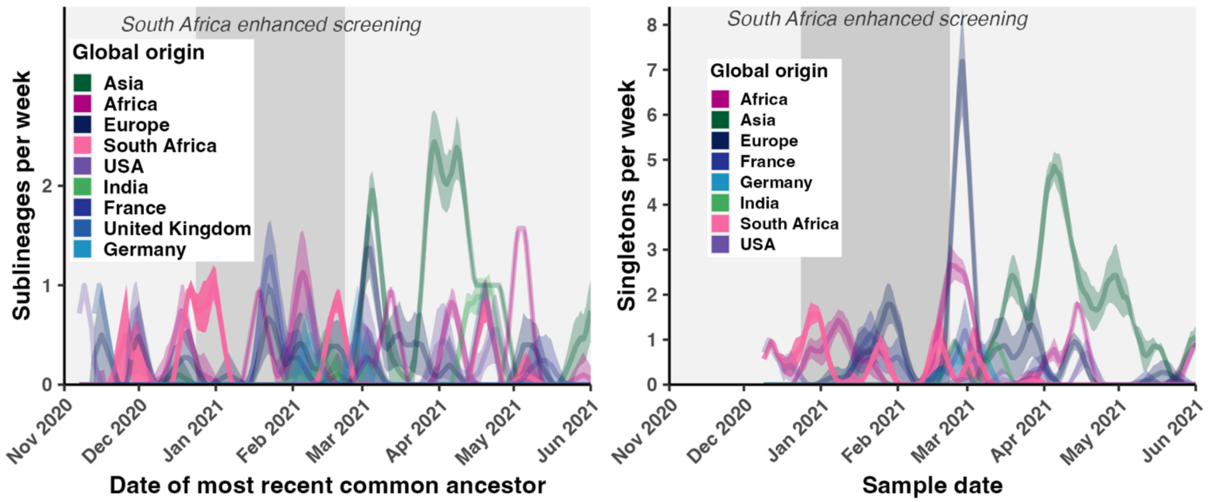


**Fig. S40. Beta sublineages and singletons imported per week into Canada.**

#### *Kappa (B.1.617.1)*

Kappa was a sister lineage to Delta (B.1.617.2) identified in India around the same time. We estimated there were 21 (20 - 21) Kappa sublineages and 81 (81 - 82) singletons.

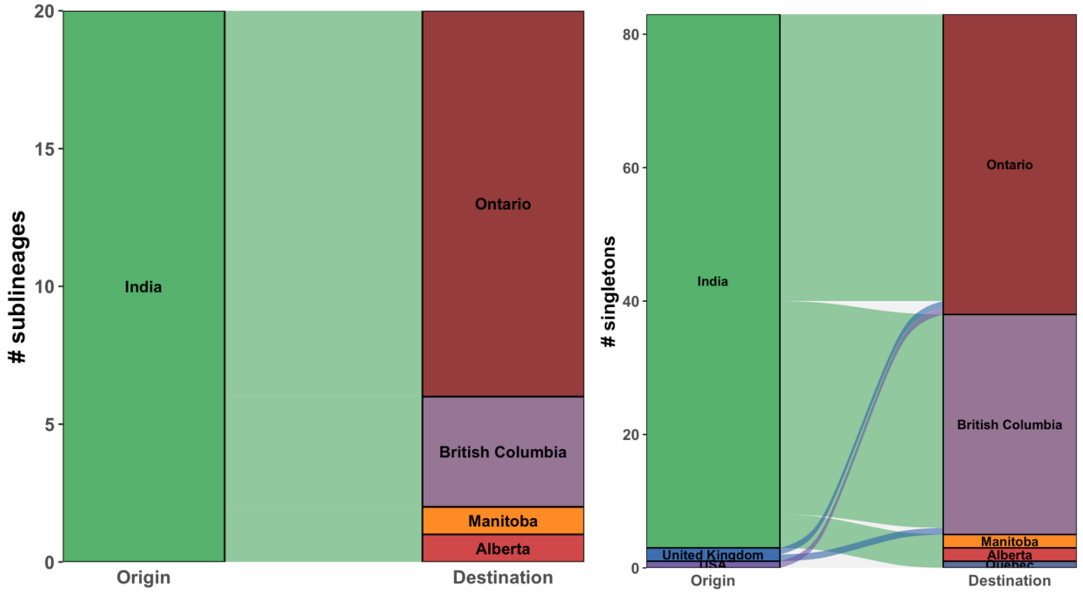


**Fig. S41**. **Kappa sublineage and singleton flows into Canada.**

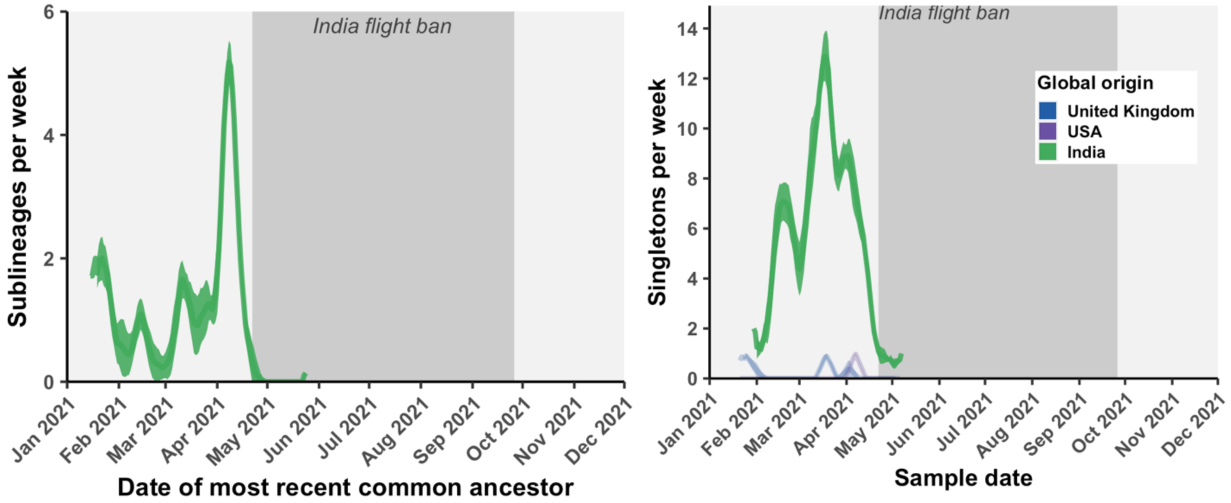


**Fig. S42. Kappa sublineages and singletons imported per week into Canada in the context of the Delta-associated India flight ban.**

***Delta***

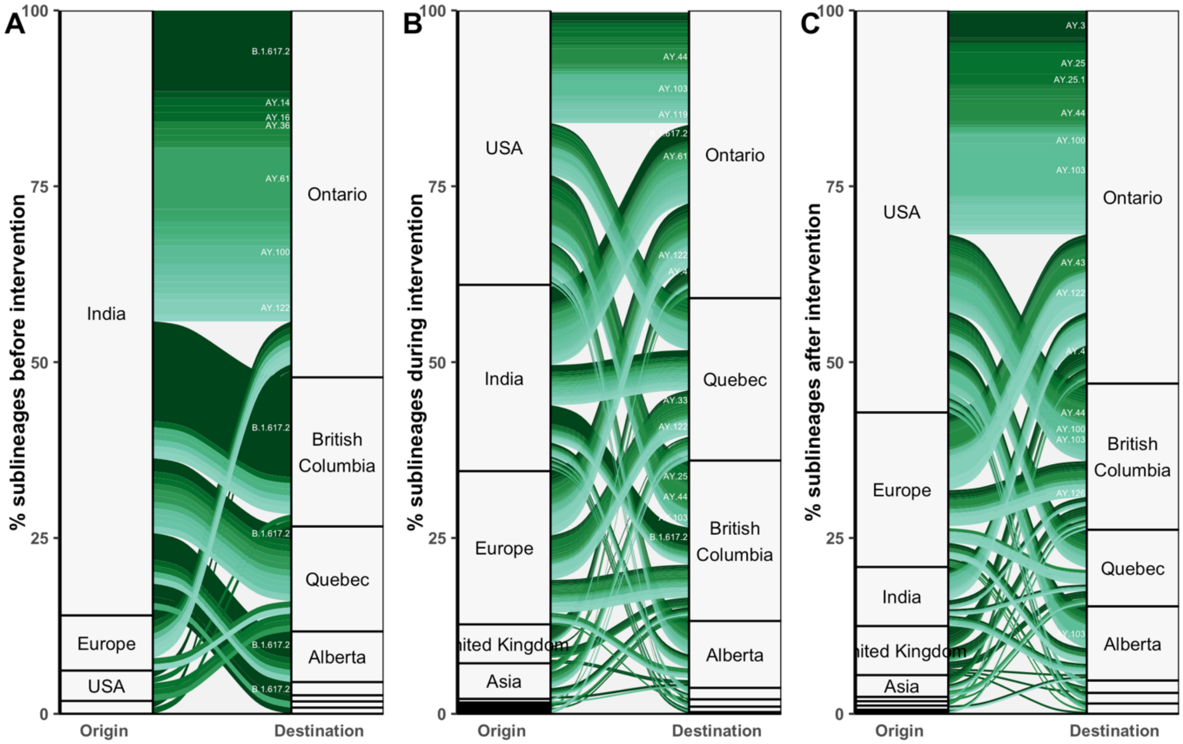


**Fig. S43. Relative contributions of global regions to Delta sublineages introduced to Canada (A) before, (B) during, and (C) after the flight ban intervention.** Sublineage flows are grouped and colored by Pango lineage.

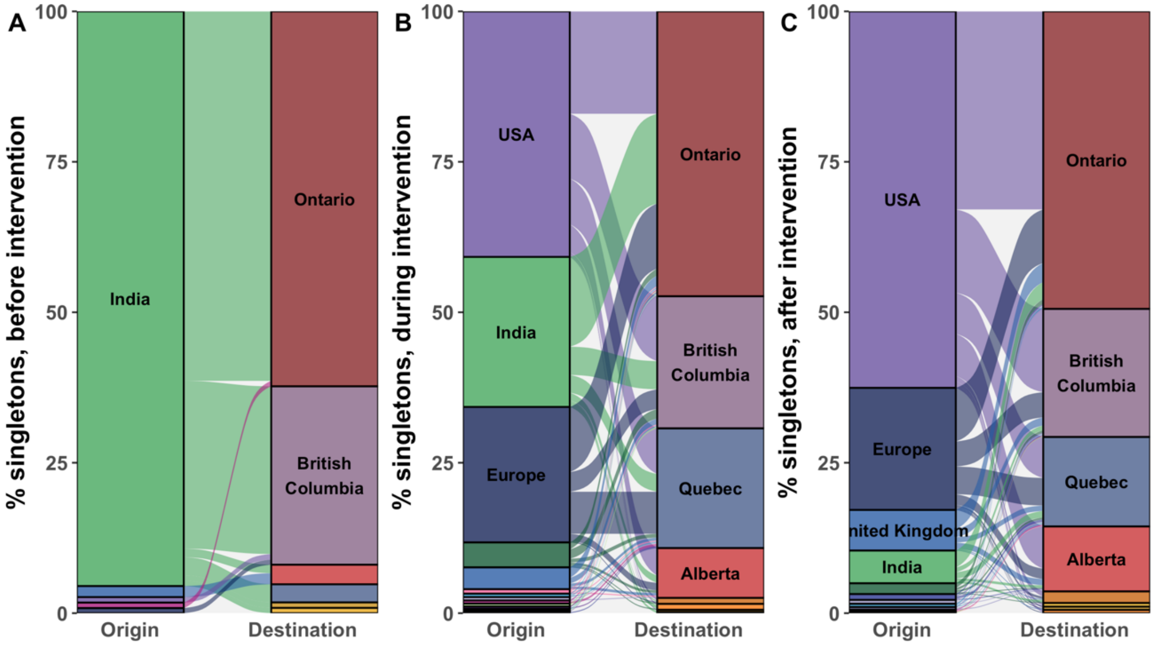


**Fig. S44. Relative contributions of global regions to Delta singletons introduced to Canada (A) before, (B) during, and (C) after the flight ban intervention.** Sublineage flows are colored by global origin.

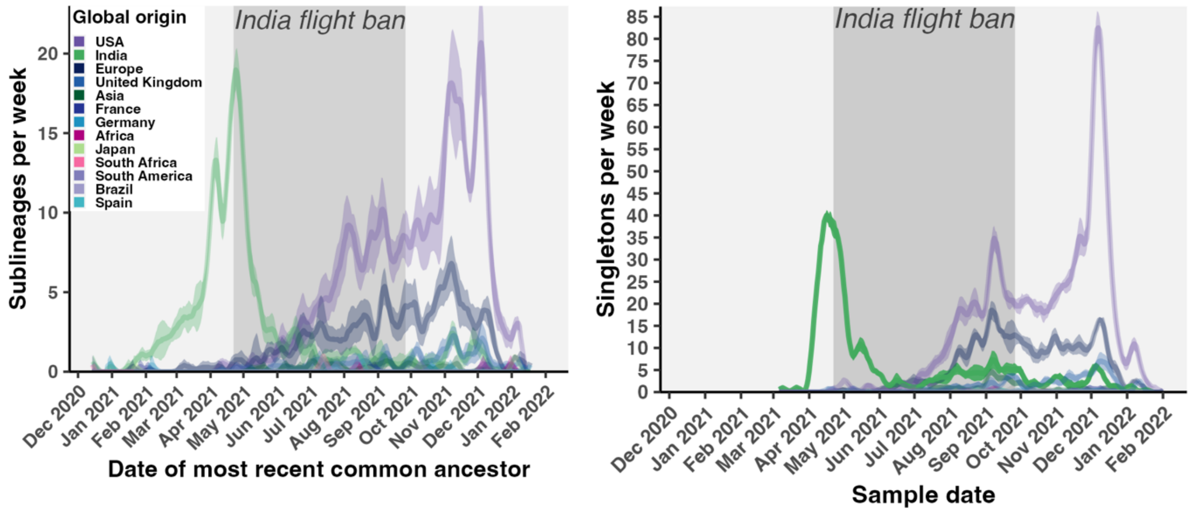


**Fig. S45**. **Delta sublineages and singletons introduced per week by global origin.**

***Iota (B.1.526)***

Iota was first identified in New York in early 2020. We inferred 14 (13 - 14) unique Iota sublineages and 37 (36 - 38) singletons introduced to Canada, almost entirely from the USA and primarily into Ontario.

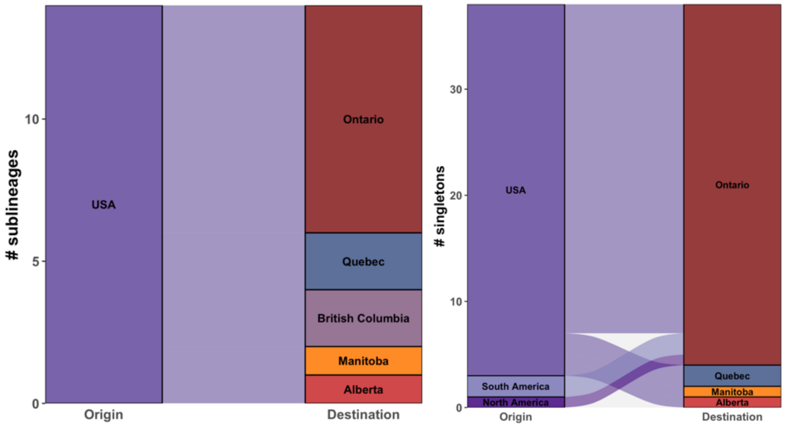


**Fig. S46. Iota sublineage and singleton flows.**

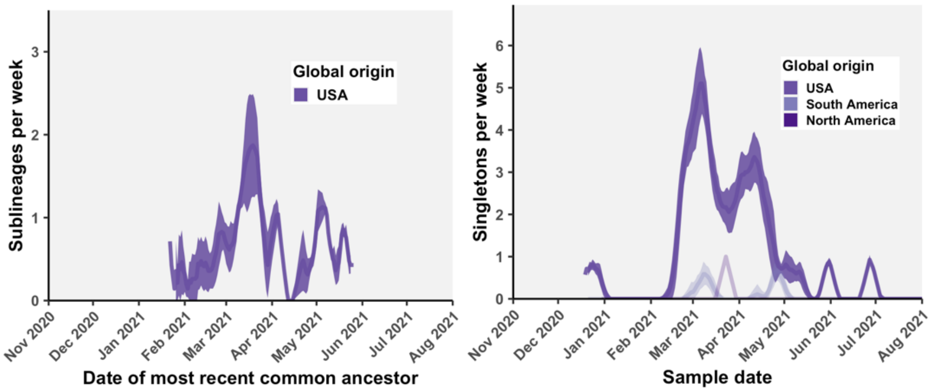


**Fig. S47. Iota sublineages and singletons introduced per week.**

***Mu (B.1.621)***

Mu was first identified in Colombia in December 2020 and first sampled in Canada in June 2021 (**Table S1**). There were at least 7 (7-7) Mu sublineages introduced to Canada and 55 (54 - 55) singletons. Despite its first discovery in Colombia, 72% of Mu sublineages and 79% of singletons were inferred to have come from the USA, with 28% of sublineages and 11% of singletons from South America. Sequences from the USA comprised 45.1% of global sequences sampled, while 39.8% were from South America.

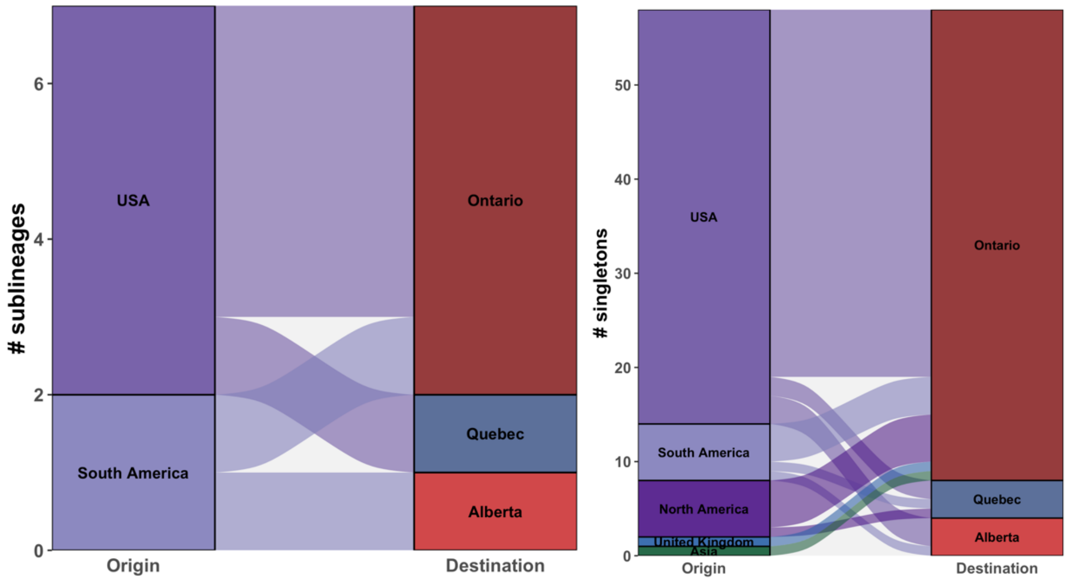


**Fig S48. Mu sublineage and singleton flows into Canada.**

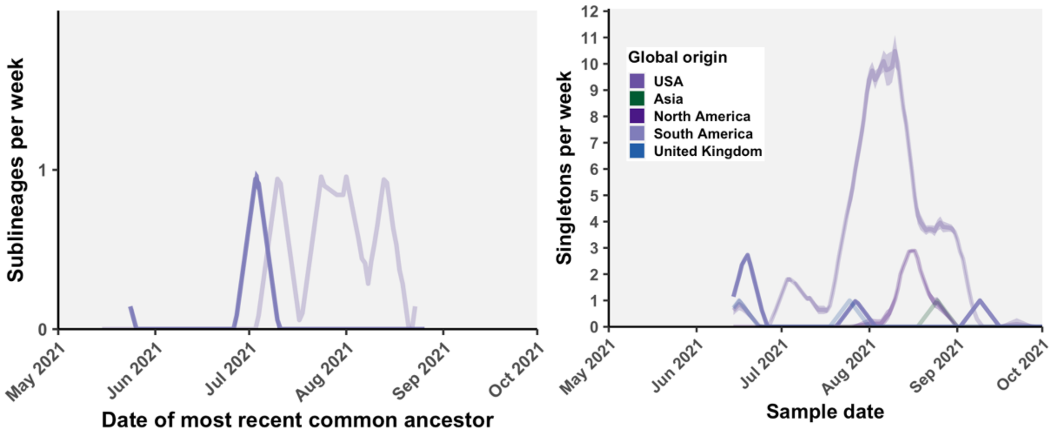


**Fig. S49. Mu sublineages and singletons imported per week into Canada.**

***Omicron (B.1.1.529; BA.*)***

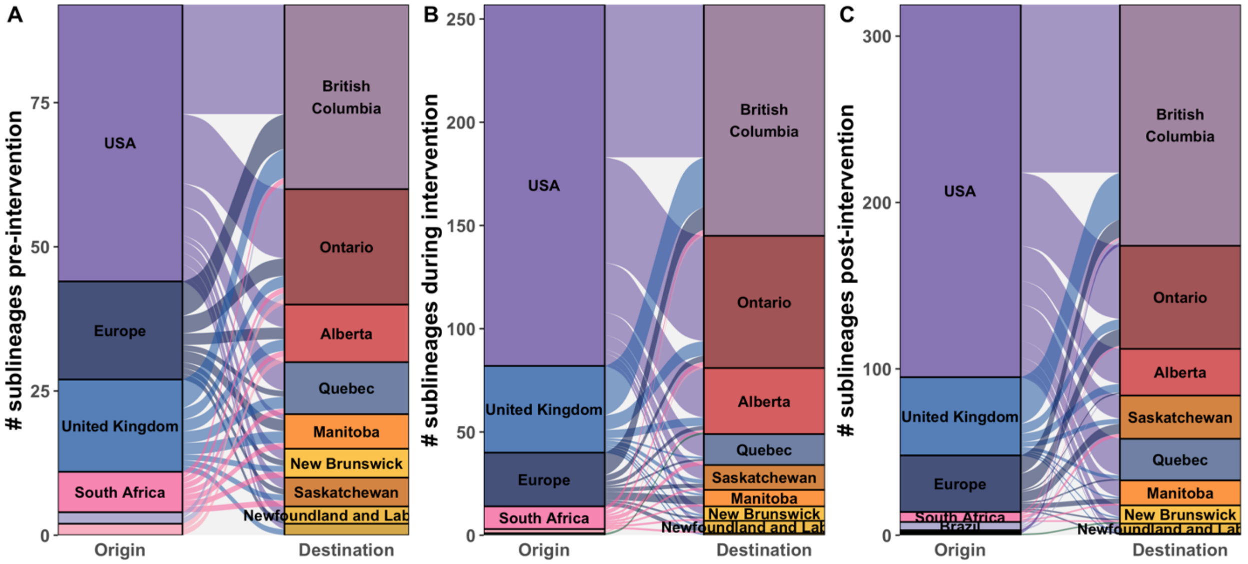


Fig. S50. Relative contributions of global regions to BA.1 sublineages (A) before, (B) during, (C) and after the travel restrictions.

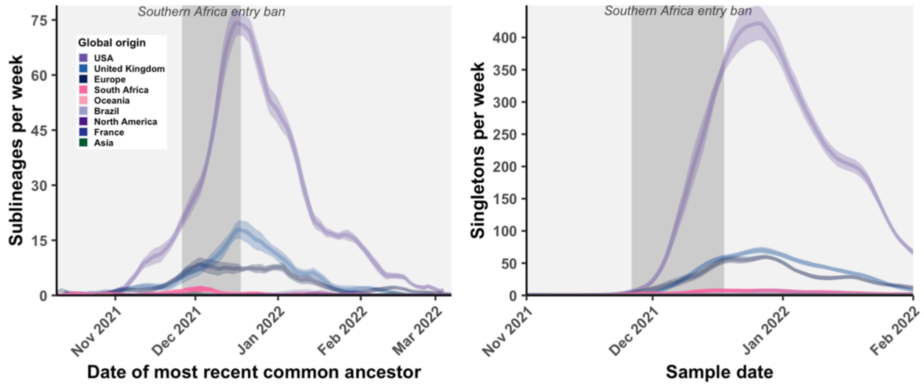


**Fig. S51. BA.1 sublineages and singletons introduced per week, in the context of the southern Africa entry ban and enhanced screening intervention.**

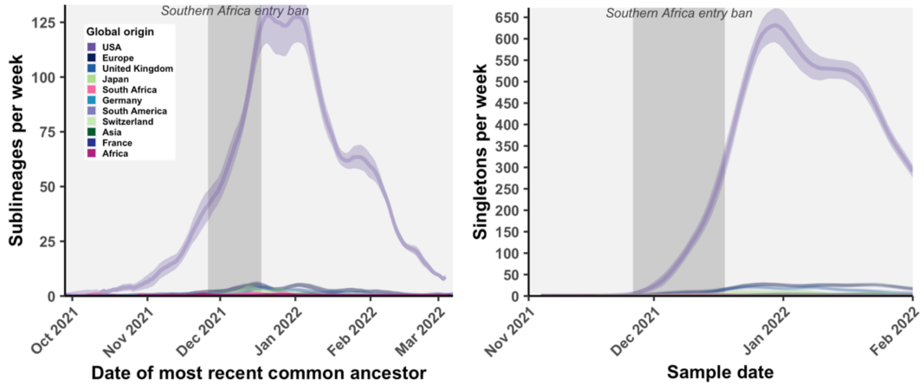


**Fig. S52. BA.1.1 sublineages and singletons introduced per week, in the context of the southern Africa entry ban and enhanced screening intervention.**

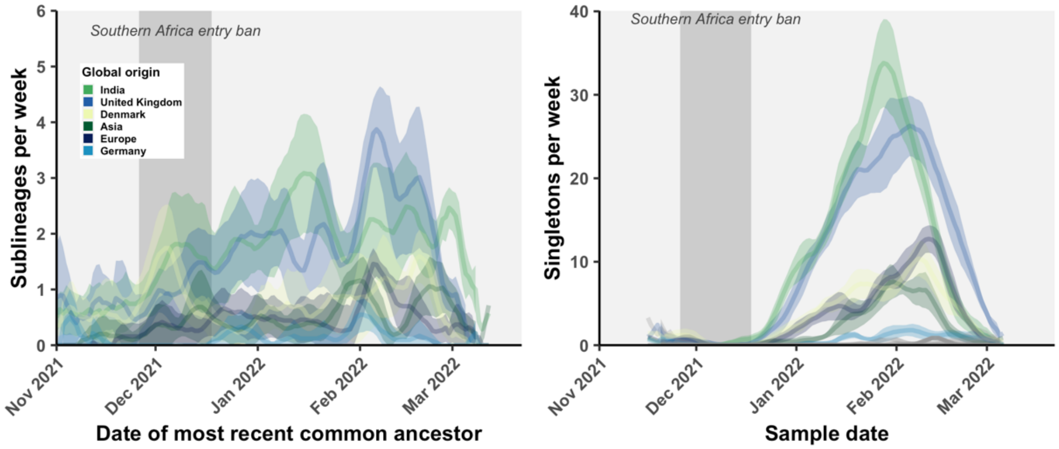


**Fig. S53. BA.2 sublineages and singletons introduced per week, in the context of the southern Africa entry ban and enhanced screening intervention.**
